# Exploring genetic risk factors for pneumonia using biobank resources - Insights across subpopulations

**DOI:** 10.1101/2024.12.09.24318706

**Authors:** Anni Heikkilä, Eeva Sliz, Sara Väyrynen, Kadri Reis, Abdelrahman G. Elnahas, Anu Reigo, Tõnu Esko, Estonian Biobank Research Team, FinnGen, Johannes Kettunen, Timo Hautala

## Abstract

**Background:** Pneumonia risk is influenced by demographics, chronic disease burden, lifestyle, and environmental factors. Despite genome-wide association studies (GWAS), the role of host genetics in pneumonia is still not fully understood, especially in certain subgroups.

**Methods:** We conducted a GWAS for pneumonia across FinnGen and Estonian Biobank (EstBB) (91,062 cases and 520,119 controls) populations. Cases with at least one pneumonia episode and subgroups based on the age at first pneumonia diagnosis, recurrent pneumonia, and asthma status were considered. We further estimated genetic correlations and causal relationships between pneumonia and other traits using linkage disequilibrium score regression and Mendelian randomization.

**Results:** We identified 13 loci including 5 replicated (*PTGER4, TNFRSF1A, CHRNA5, MUC5A, HLA*) and 8 novel associations (*PTPN22, CRP, APOE, FHIT, MAPKAPK2, TNFSF15, HNF1A, RIN3*) mainly harboring genes regulating immunity or lung health associated with pneumonia across subgroups. Interestingly, novel associations included the *APOE* locus with the ε4 allele having a protective effect on pneumonia. Additionally, we report genetic correlations between pneumonia and 204 other traits, and nine traits potentially causal to pneumonia risk.

**Conclusions:** Loci associated with pneumonia harbour genes mainly related to acute inflammation, T cell development, antigen presentation and lung health. The stratified analyses demonstrate differences in genetic risk factors contributing to development of pneumonia in different patient subgroups. In summary, the findings of our study highlight the importance of immunological mechanisms at early life and in adults as well as significance of lifestyle and lung health among the elderly.

## Introduction

Pneumonia is the leading infectious cause of hospitalisation and death. ^1^ Young children, immunocompromised patients and the elderly are at increased risk. ^2^ Low socioeconomic status, smoking and chronic diseases are also risk factors for pneumonia. ^3,4^ Although pneumonia is caused by an infectious agent, host responses contribute to its pathogenesis. ^5^ ^6^ Genetic and immunological mechanisms of pneumonia susceptibility are understood in selected inborn errors in immunity conditions such as common variable immunodeficiency (CVID). ^7^ Genome-wide association studies (GWAS) have also identified population level pneumonia risk associations. ^8^ ^9^ ^10^ ^11^ ^12^

Previous GWASs have focused on general pneumonia, childhood pneumonia or pneumonia caused by COVID-19. Our current study utilises well-defined data from two large populations, FinnGen (data freeze 10) and the Estonian Biobank (EstBB). Importantly, large population size in our study allowed genetic analyses of pneumonia subgroups in age strata, among asthmatic pneumonia patients and those suffering from recurrent pneumonia. Our study design also allowed estimation of genetic correlations and causal relationships of pneumonia using linkage-disequilibrium score (LDSC) regression and Mendelian randomization (MR), respectively.

## Methods

### Study populations

FinnGen and EstBB study populations, case definitions through the International Classification of Diseases version 10 (ICD-10) codes and the rationale of the analyses are described in detail in **Supplementary Methods**. We evaluated data from up to 91,062 patients with at least one pneumonia episode and 520,119 controls in FinnGen and EstBB across six distinct GWAS analyses: general pneumonia (91,062 cases), pneumonia in age categories according to the age at first diagnosis (“children”, 0 to 16 years of age (6,641 cases), “adults”, 16 to 60 years of age (40,436 cases) and “elderly”, those over 60 years of age (44,053 cases)), recurrent pneumonia (at least three pneumonia episodes; 8,199 cases) and pneumonia among asthmatic patients (19,520 cases, 53,212 controls). We also tested the genetic associations in hospitalised cases (44,906 cases) as a sensitivity analysis. **Supplementary Table 2** provides more detailed case definitions.

Patients and controls in FinnGen provided written informed consent for biobank research, based on the Finnish Biobank Act. Alternatively, separate research cohorts, collected prior the Finnish Biobank Act came into effect (in September 2013) and start of FinnGen (August 2017), were collected based on study-specific consents and later transferred to the Finnish biobanks after approval by Fimea (Finnish Medicines Agency), the National Supervisory Authority for Welfare and Health. Recruitment protocols followed the biobank protocols approved by Fimea. The Coordinating Ethics Committee of the Hospital District of Helsinki and Uusimaa (HUS) statement number for the FinnGen study is Nr HUS/990/2017. The activities of the Estonian Biobank are regulated by the Human Genes Research Act, which was adopted in 2000 specifically for the operations of the Estonian Biobank.

## Results

### Meta-analysis of genetic predisposition of pneumonia

We identified altogether 13 loci showing genome-wide significant association (P<5×10^-8^) with pneumonia across subgroups. 8 of these loci are previously unreported in pneumonia GWASs. Lead variants are summarised in **Table 1**, and the Manhattan plots of results are shown in **Figure 1**. **Figure 2** shows an effect estimate comparison of the lead variants between subpopulations. Regional association plots of significant associations can be found from **Supplementary Figures 1-28**. We estimated SNP-based heritability and genomic inflation factor for each of the subgroups utilising the LDSC command line tool. ^13^ Due to the difficulty in establishing population prevalence for pneumonia, we used our sample prevalence as a population prevalence for the calculation, as in earlier pneumonia GWAS studies. ^10^ ^11^ We report following SNP-based heritability estimates: general pneumonia 0.026 (standard error 0.0024), pneumonia in children 0.019 (0.0099), pneumonia in adults 0.027 (0.0033), pneumonia in elderly 0.018 (0.0035), recurrent pneumonia 0.038 (0.0092) and pneumonia among asthmatics 0.001 (0.0015). The estimate for pneumonia in the general population is in accordance with estimates from earlier studies, 0.03 ^10^ and 0.0269.^11^ Genomic inflation factor estimates, however, suggest minor inflation in test statistics (general pneumonia 1.15, pneumonia in children 1.04, pneumonia in adults 1.12, pneumonia in elderly 1.12, recurrent pneumonia 1.07 and pneumonia among asthmatics 1.04). However, minor inflation is likely due to polygenic signals, given the intercepts of 1.04, 1.02, 1.04, 1.06, 1.03 and 1.04, respectively.

**Figure 1.**
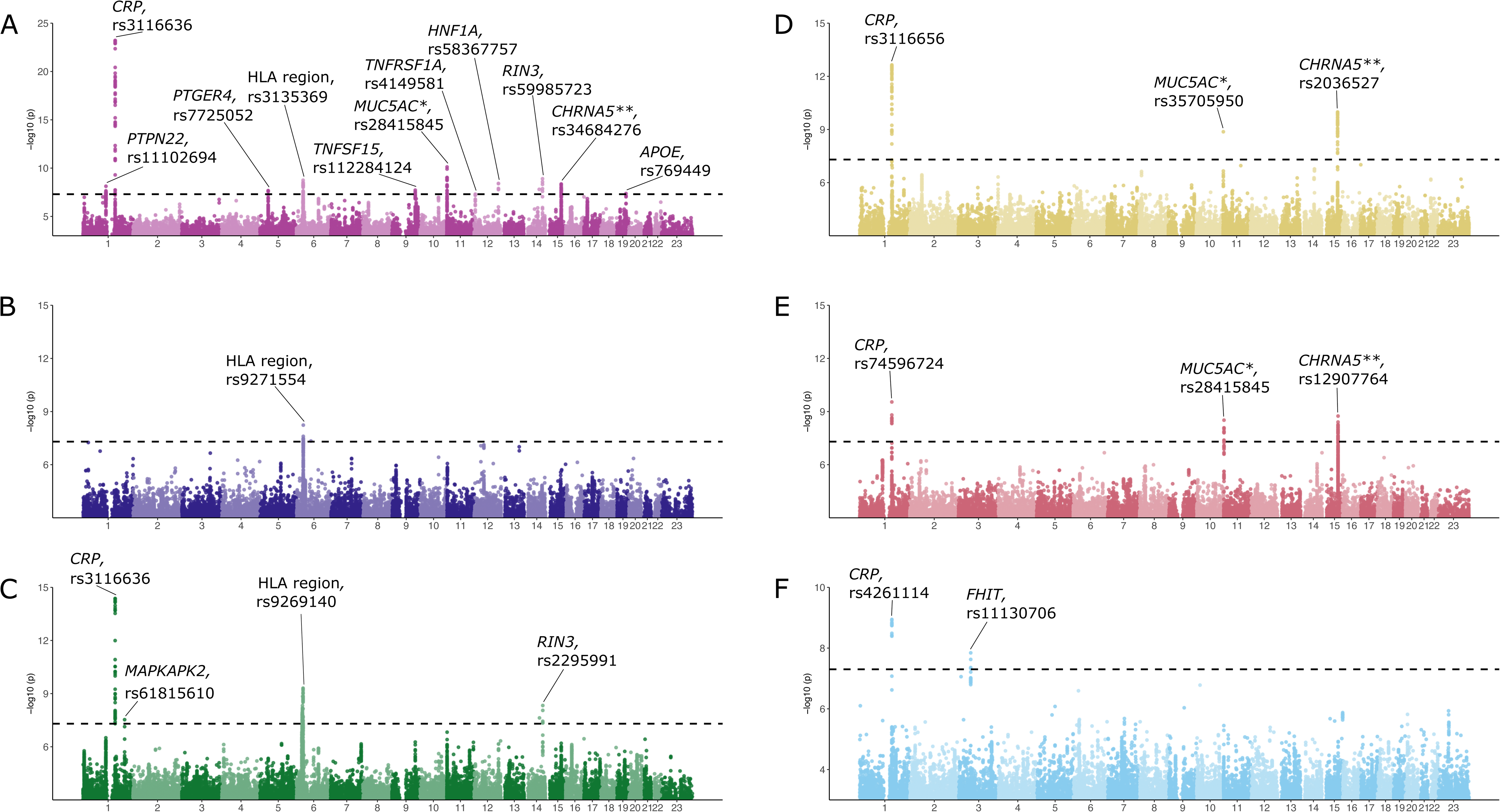
Manhattan plots of genome-wide meta-analysis (FinnGen and Estonian Biobank) of pneumonia (A), pneumonia in under 16-year-olds (B), pneumonia in 16-60 year-olds (C), pneumonia in over 60 year olds (D), recurrent pneumonia (E), pneumonia among asthma patients (F). Candidate genes and rs-numbers for novel lead variants are marked in each plot. Y-axis represents the -log10 p-value. The genome-wide significance threshold is marked with black dashed horizontal line. * This locus has multiple candidate genes, but for clarity, only one is included in the figure. All candidate genes of the area are *MUC6*, *MUC2*, *MUC5AC* and *MUC5B*. ** This locus has multiple candidate genes, but for clarity, only one is included in the figure. All candidate genes of the area are *IREB2, HYKK, CHRNA5, CHRNA3 and CHRNB4*.

**Figure 2.**
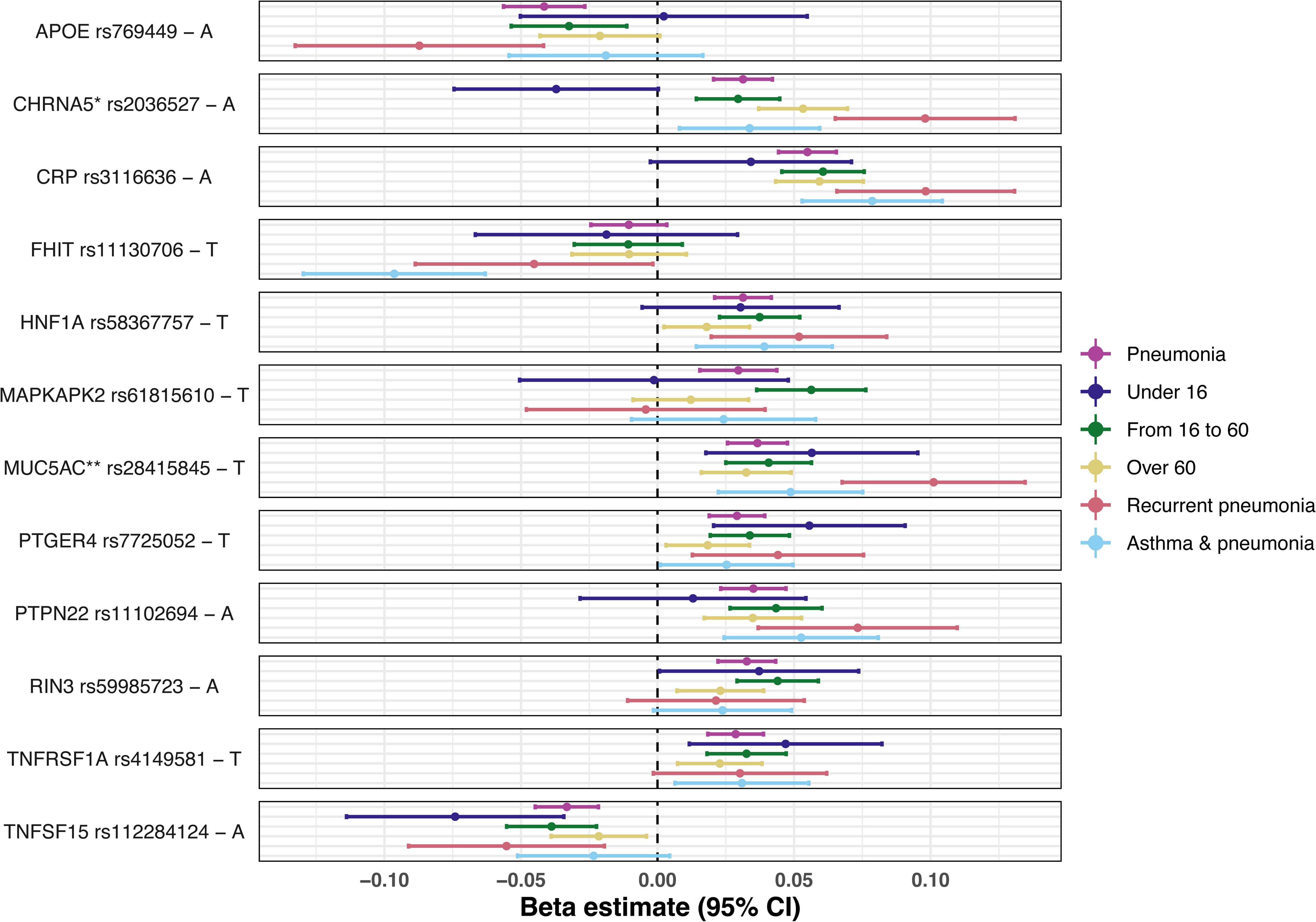
Effect estimates of lead variants across all analytical subgroups in FinnGen and Estonian Biobank meta-analysis. In case of multiple different lead variants in the area, the most significant in any subgroup was picked for the plot. The effect sizes in log OR scale (x-axis) and corresponding 95% confidence intervals are given for each lead variant (y-axis). The dashed vertical line indicates the beta value 0. Color coding of the analysis groups is shown in the legend on the right side of the plot; for each variant panel, the order of the groups always goes as follows from top to down: pneumonia, pneumonia in under 16 year olds, pneumonia in from 16 to 60 year olds, pneumonia in over 60 years olds, recurrent pneumonia, pneumonia among asthmatics. * This locus has multiple candidate genes, but for clarity, only one is included in the figure. All candidate genes of the area are *IREB2, HYKK, CHRNA5, CHRNA3 and CHRNB4*. ** This locus has multiple candidate genes, but for clarity, only one is included in the figure. All candidate genes of the area are *MUC6*, *MUC2*, *MUC5AC* and *MUC5B*.

**Table 1.**
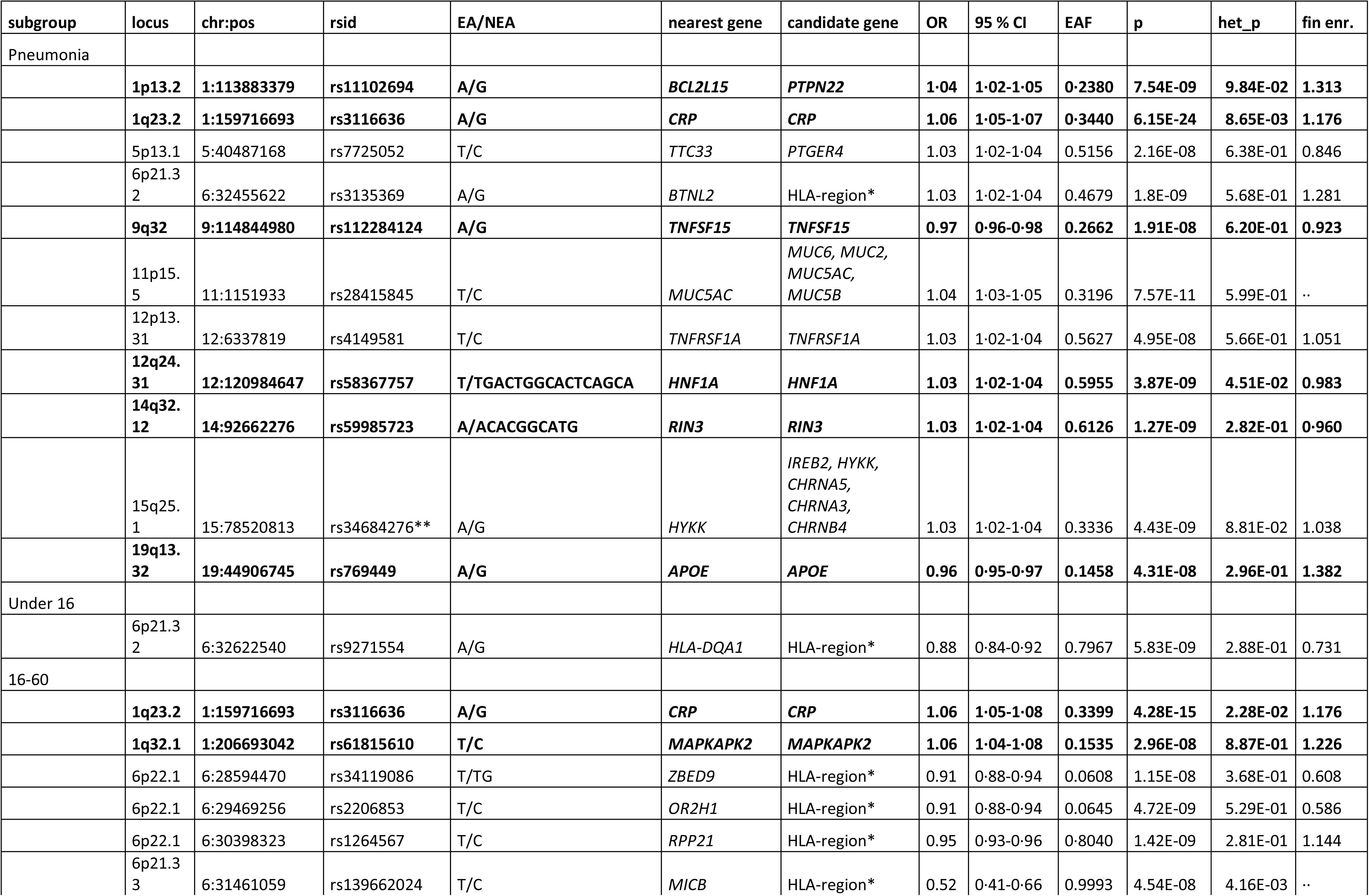
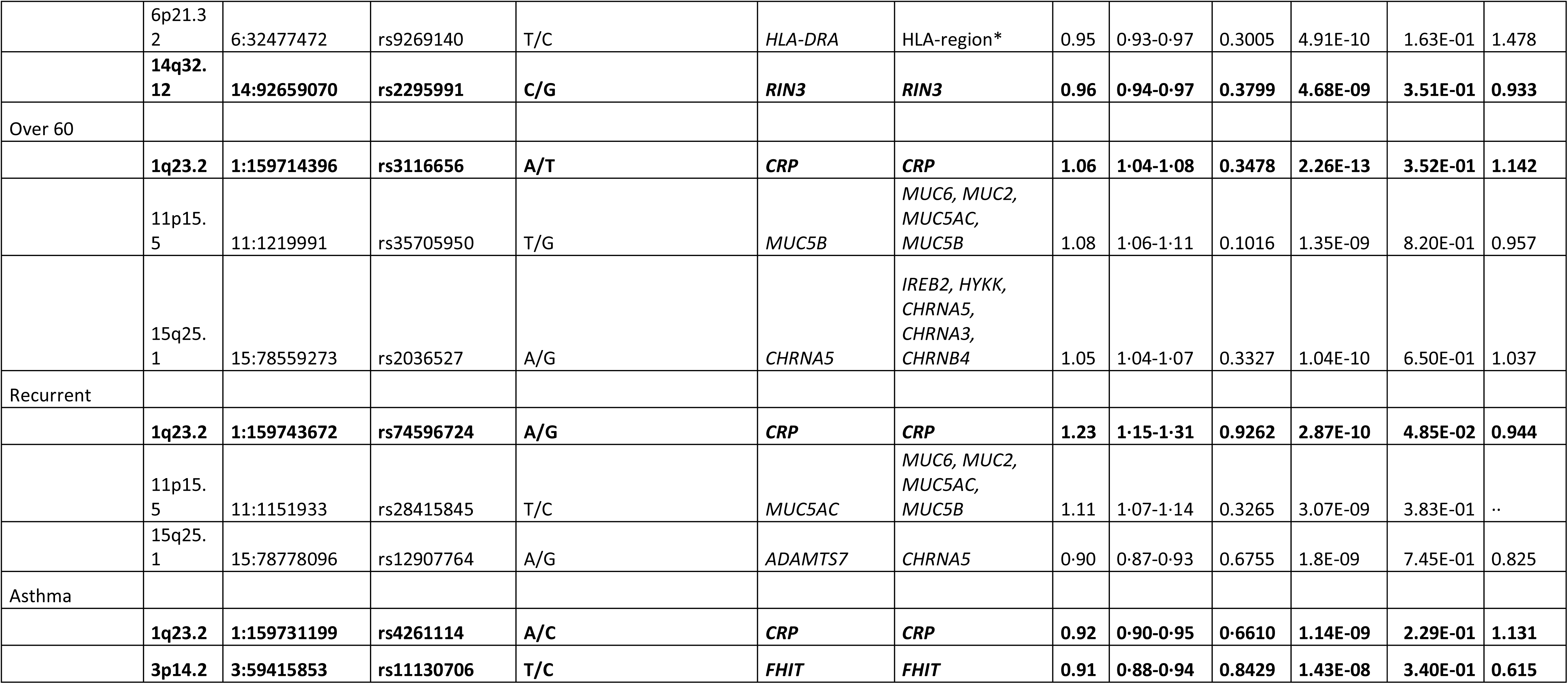
Pneumonia-associated loci in the general population and among patient subgroups. The table lists lead variants at loci associated with pneumonia, identified at genome-wide significance (p<5x10-8) in the genome-wide meta-analyses combining data from FinnGen and Estonian Biobank. Cases and controls in subgroups: Pneumonia, 91,062 cases, 520,119 controls; Under 16, 6,641 cases, 520,440 controls; 16-60 40,436 cases, 520,440 controls; Over 60, 44,053 cases, 520,440 controls; Recurrent, 8,199 cases, 524,171 controls; Asthma, 19,520 cases, 53,212 controls.Variants highlighted in bold have not been reported in previous pneumonia GWASes. EA/NEA, effect allele/non-effect allele; candidate gene, gene most likely to cause the association according to our literature-based annotation; OR, odds ratio (for effect allele); 95 % CI, 95% confidence interval for odds ratio; EAF, effect allele frequency; p, p-value of the association; het_p, meta-analysis heterogeneity p-value; fin enr., Finnish enrichment of the variant (calculated by comparing allele frequency in FinnGen individuals to allele frequency in gnomAD non-Finnish, Swedish or Estonian European individuals). *Candidate genes are not specified due to high LD in the HLA-region. ** Previously also known as rs113858005.

Our main analysis consisted of cases with at least one pneumonia episode (91,062 cases and 520,119 controls; **Figure 1A**, **Table 1**). Our novel findings include pneumonia association near protein tyrosine phosphatase non-receptor type 22 (*PTPN22*), a gene that encodes a regulator of T-cell receptor signalling pathways associated with autoimmunity and infections. ^14^ We also identified novel pneumonia associations near the genes encoding C-reactive protein (*CRP*), a product of which can be used as an indicator of acute inflammation, apolipoprotein E (*APOE*) encoding a protein known for its role in lipid metabolism Ras interaction protein 3 (*RIN3*), vascular endothelial cell growth inhibitor *TNFSF15* and hepatic nuclear factor 1 alpha (*HNF1A*). ^15^ We replicated associations near *MUC5AC*, *TNFRSF1A*, *CHRNA5* and *PTGER4* genes and the HLA region.

### Age-stratified pneumonia

In pneumonia among ‘children’, (6,641 cases, 520,440 controls), an association was identified in the HLA region (**Figure 1B**, **Table 1**). In the ’adult’ cohort, (40,436 cases, 520,440 controls), associations near *CRP*, MAPK activated protein kinase 2 (*MAPKAPK2*), HLA region and *RIN3* were identified. (**Figure 1C**, **Table 1**). ^16^ Among the ‘elderly’ population (44,053 cases, 520,440 controls), associations near *CRP*, *MUC5B* and *CHRNA5* (**Figure 1D**, **Table 1**) were found. ^17^

### Recurrent pneumonia

Among those suffering from recurrent pneumonia episodes (8,199 cases, 524,171 controls), we identified associations near *CRP*, *MUC5AC*, and *CHRNA5* (**Figure 1E**, **Table 1**).

### Pneumonia among asthmatic patients

Analysis among asthmatics (19,520 cases with both asthma and pneumonia, 53,212 controls with asthma but no pneumonia) revealed associations near *CRP* and fragile histidine triad diadenosine triphosphatase (*FHIT*) (**Figure 1F**, **Table 1**). ^18^

### Pneumonia treated at hospital among FinnGen population

In the FinnGen, we analysed the genetic associations of hospitalised pneumonia cases (44,906 cases) (**Supplementary Figure 29**, **Supplementary Table 3**). Since almost 70 % of all cases had been hospitalised, the effect estimates were similar when compared to the general pneumonia population.

### Comparison of effect estimates across pneumonia subgroups

Effect estimates of the lead variants across subgroups were compared (**Figure 2**). In the case of multiple lead variants in the locus, the variant with the smallest p-value was evaluated. The effect estimate of locus containing nicotine dependency-associated genes, *CHRNA5* rs2036527-A, among “children” is nonsignificant consistent with a limited role for smoking in childhood pneumonia, whereas a positive association in other subgroups is observed. The same allele shows a significantly larger risk-increasing effect in the “recurrent pneumonia” subgroup compared to others (**Supplementary Table 4**). Likewise, the effect of the *MUC5AC* rs28415845-T is significantly larger in the “recurrent pneumonia” subgroup compared to most other subgroups (**Supplementary Table 4**). Further, *FHIT* rs11130706- T, a locus associated with pneumonia in the asthmatic subgroup, has a significantly larger effect estimate among asthmatics when compared to others.

### Effect estimates of HLA alleles

Role of HLA alleles associated with pneumonia were further assessed in the FinnGen population. The HLA alleles significantly (false discovery rate (FDR)-corrected p-value < 0.05) associated with at least one pneumonia subgroup are displayed in **Figure 3**. Full list of results can be found in **Supplementary Table 5**. Several HLA alleles, such as *DQB1*03:02* or *DQA1*03:01*. *DQB1*03:02*, *DQA1*03:01* and *DRB4*01:03* alleles are strongly associated with recurrent pneumonia. Interestingly, multiple alleles (*DRB3*01:01*, *DRB1*03:01*, *DQB*02:01*, *DQA1*05:01*, *A:01:01*) suggest protective effects for pneumonia, especially among the adult subgroup.

**Figure 3.**
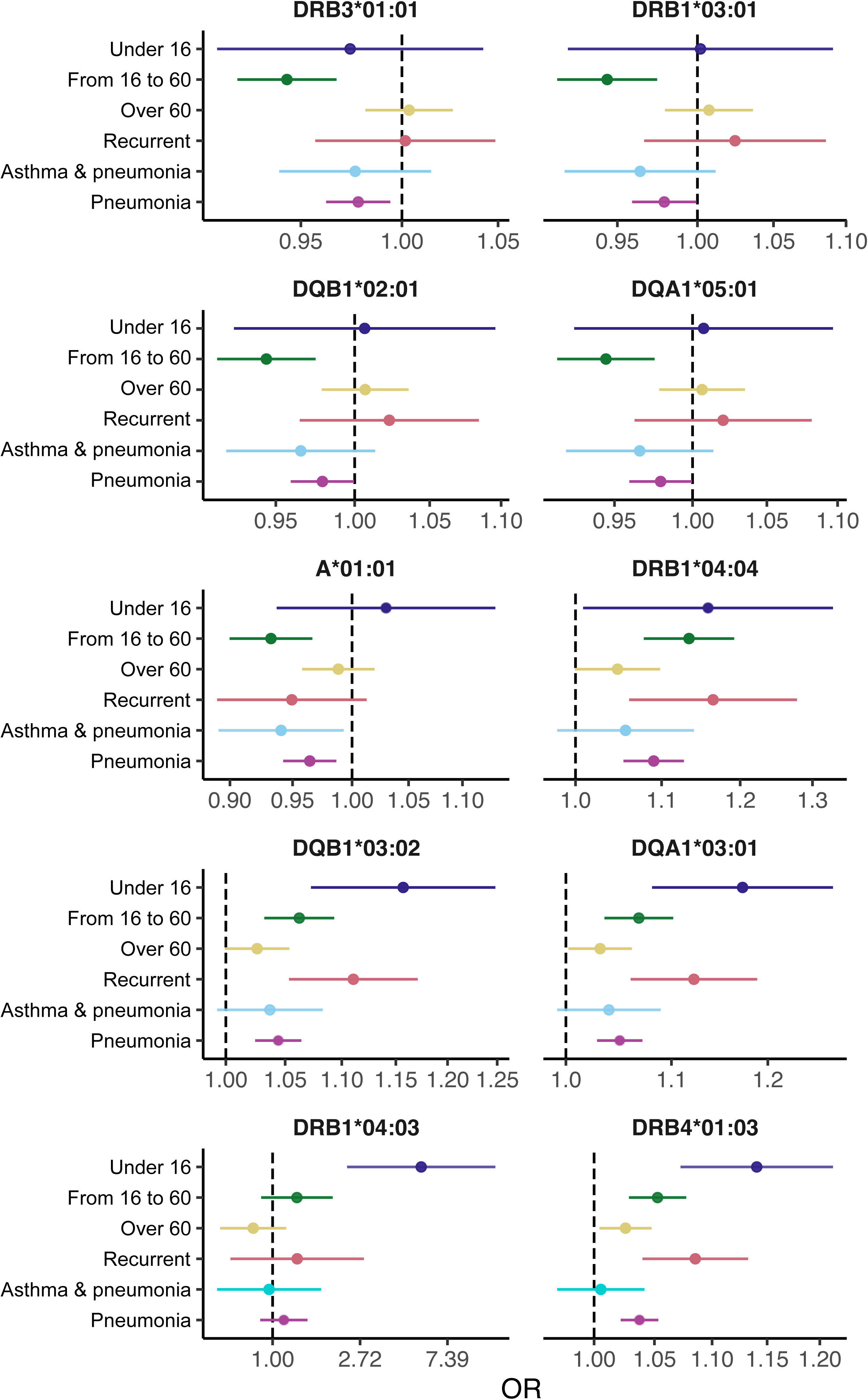
Effect estimates of HLA alleles showing significant association with at least one pneumonia subgroup. Name of the allele is shown above each panel. Each panel shows the name of the pneumonia subgroup in the y-axis and odds ratio on the x-axis, whiskers show 95 % confidence interval. Vertical dashed line show location of OR = 1.0.

### Pneumonia risk and APOE alleles

*APOE* gene, a known risk factor for multiple diseases, is associated with general pneumonia risk. We found that *APOE* ε4 is associated with a lower risk of pneumonia especially among adults, asthmatics and those with recurrent pneumonia (per allele effects shown in **Figure 4**; full genotypes in **Supplementary Figure 30**). The protective effect size was largest in the recurrent pneumonia and asthmatic subgroups, but the most significant p-values were found among “adults” and those with at least one pneumonia episode (**Supplementary Tables 6-7**).

**Figure 4.**
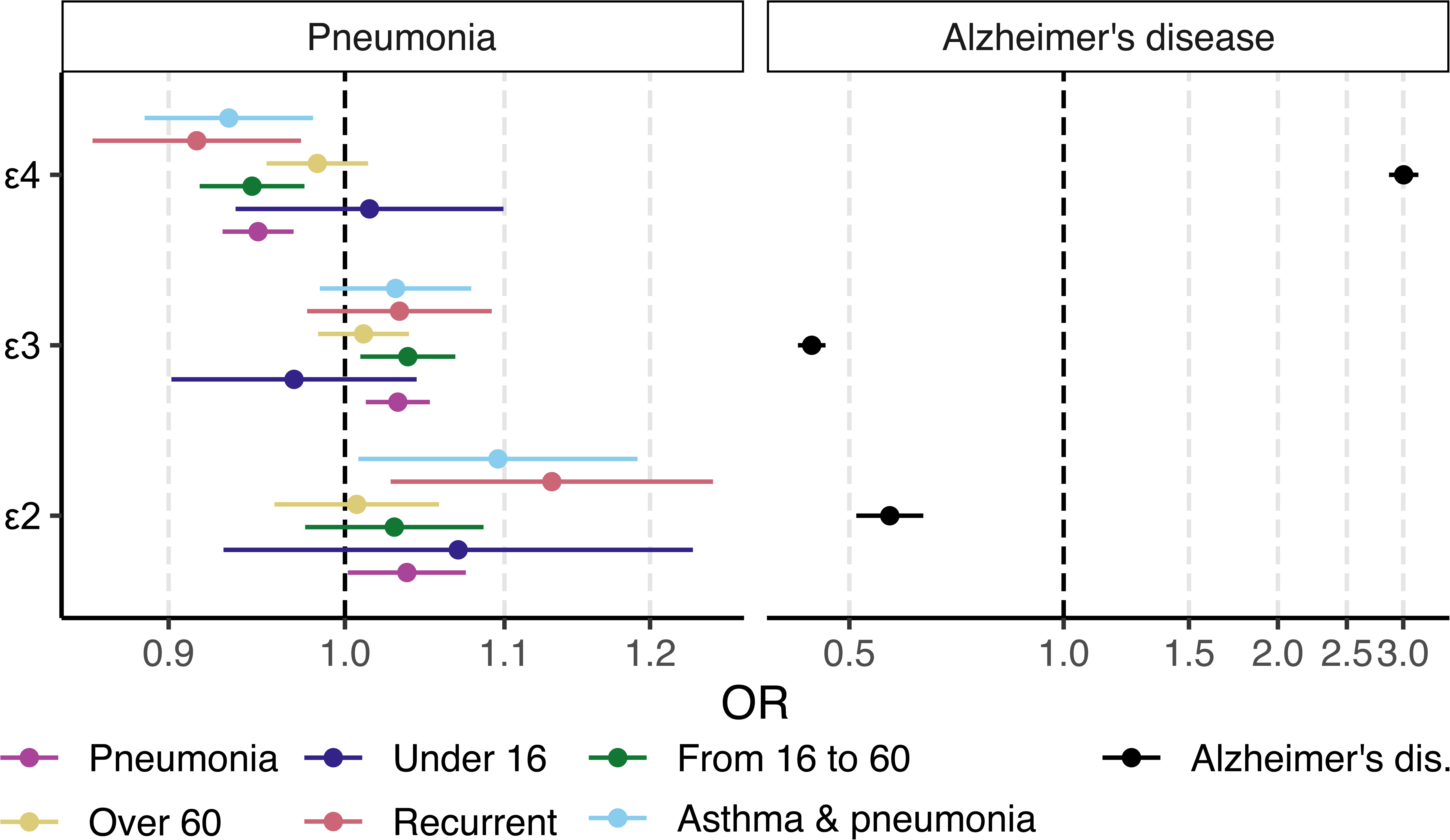
Effect estimates of *APOE* alleles in pneumonia subgroups in FinnGen. Left panel shows effect estimates for pneumonia, and the right shows effect estimates for Alzheimer’s disease. Effects are given per copy of each allele, derived from a logistic regression assuming additive effects of the alleles. Y-axis shows *APOE* alleles and x-axis shows odds ratio for the estimates. Vertical dashed line shows the place of value 1. Legend below the figure shows color coding for pneumonia subgroups.

### Gene expression colocalizations

We found 7 significant colocalizations between pneumonia signals and gene expression in tissues relevant to pneumonia (**Supplementary Table 8**, **Supplementary Figures 31-41**). Out of manually curated candidate genes, *CHRNA3* expression in lungs colocalized with pneumonia association signal in chromosome 15. We found two colocalizations with long non-coding RNAs: pneumonia association signals colocalized with *AP4B1-AS1* expression in lung near our candidate gene *PTPN22*, and with *RP11-754B17.1* expression in EBV-transformed lymphocytes near mucin gene cluster in chromosome 11.

### Genetic correlations

204 traits were genetically correlated with pneumonia (**Supplementary Table 9**). Top correlated traits were related to smoking (e.g., “Current tobacco smoking” [genetic correlation (rg) = 0.460, P=6.984×10^-30^], “Smoking status: Never” [rg = -0.359, P =2.280×10^-21^]), higher body weight (e.g., “Body mass index (BMI)” [rg = 0.317, P=2.955×10^-26^], “Whole body fat mass” [rg = 0.298, P=6.114×10^-26^]), other health problems (e.g., “Wheeze or whistling in the chest in the last year” [rg = 0.496, P=8.221×10^-39^], “Diagnoses -main ICD10: R07 Pain in throat and chest” [rg = 0.523, P=4.257×10^-16^]) or lower education level (“Years of schooling” [rg = -0.310, P=2.570×10^-25^], “Qualifications: College or university degree” [rg = -0.316, P=6.142×10^-22^].

### Mendelian randomization

In bidirectional two-sample MR ^19^, we found 9 traits possibly exposing to pneumonia (**Figure 5**) :“Whole body fat mass” (b = 0.166, P=1.41×10^-17^), “Overall health rating” (b = 0.480, P=4.99×10^-15^), “Whole body fat-free mass” (b = 0.137, P=7.45×10^-9^), “Years of schooling” (b = -0.181, P=1.37×10^-8^), “Pack years of smoking” (b = 0.318, P=1.97×10^-7^), “Frequency of tiredness/lethargy in the last 2 weeks” (b = 0.401, P=2.62×10^-5^), “Usual walking pace” (b = -0.373, P=4.23×10^-5^) and “Wheeze or whistling in the chest in the last year” (b = 0.903, P=0.0003). Full description of the results is available in **Supplementary Tables 10** and **11**.

**Figure 5.**
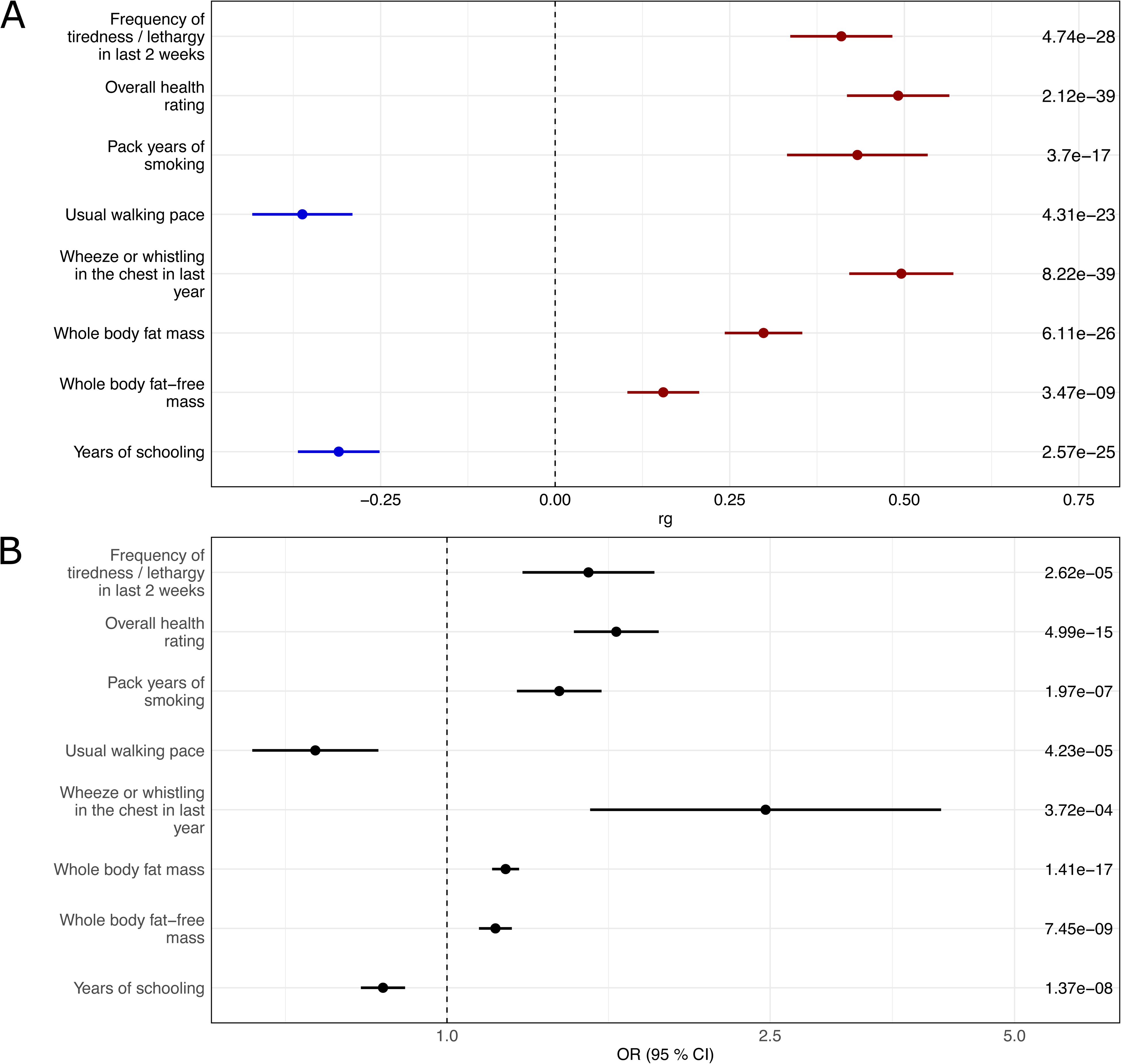
Forest plot of genetic correlations (GC) (A), and significant MR associations (B). The plot displays the results for traits that reached statistical significance in the MR analyses. A) The y-axis includes names of the traits (on the left side of the plot) and p-values of the correlations (on the right side of the plot). The x-axis shows the genetic correlation value, the vertical dashed line shows the place of value 0. The rg value of each association is indicated by a dot, the whiskers around the dot show the 95 % confidence interval of the value. The results for all 204 significantly correlated traits can be found in Supplementary Table 7. C) The y-axis includes names of the traits tested as exposure for pneumonia (on the left side of the plot) and p-values of the causal effects (on the right side of the plot). The x-axis shows the odds ratio for the causal effects; the vertical dashed line shows the place of odds ratio value 1. Dots show exact odds ratio value, whiskers show the 95 % confidence interval for the odds ratio. The results for all tested traits can be found in Supplementary Tables 8 and 9.

## Discussion

Our findings derived from up to 91,062 pneumonia cases and 520,119 healthy controls replicated previous findings (*PTGER4, TNFRSF1A, CHRNA5, MUC5A, HLA*) and identified novel genetic associations (*PTPN22, CRP, APOE, FHIT, MAPKAPK2, TNFSF15, HNF1A, RIN3*) of pneumonia. While the associations among 6,641 childhood pneumonia cases are limited to HLA locus, pneumonia risk among the 40,436 adults is associated with several immunologically active genes and genes involved in development of chronic lung diseases. Among the elderly (n=44,053) and those with recurrent pneumonia (n=8,199), associations near genes involved in mucus production (*MUC5AC*) and nicotine dependency (*CHRNA5*) are most obviously implicated. We further found a unique genetic association near *FHIT* among 19,520 asthmatics with pneumonia. In conclusion, the differences in genetic associations between subgroups of age, asthma or recurrence highlight the view that several genetic and biological mechanisms are involved in pneumonia susceptibility.

Hyperinflammatory pneumonia patients commonly suffer from complications and mortality. ^6^ For example, *MAPKAPK2* and *PTPN22* are well recognised regulators of proinflammatory interleukin-1 (IL-1) expression while *PTPN22* is also implicated in primary antibody deficiency and autoimmunity. ^20^ ^21^ *TNFRSF1A* is responsible for autoinflammatory tumor necrosis factor receptor-associated periodic syndrome ^7^ ^11^ while *TNFSF15* induces proinflammatory cytokines, costimulates T cells and innate lymphoid cells and promotes differentiation of IL-9-producing T cells. ^22^ Hepatocyte nuclear factor-1 alpha (HNF-1α) regulates the expression of acute phase proteins, such as fibrinogen, CRP, and IL-1 receptor. In summary, genetic mechanisms involved in regulation of inflammation and T cell biology may mediate the risk of pneumonia especially among the adult population.

HLA molecules present intracellular (major histocompatibility complex (MHC) class I) and extracellular (MHC class II) infectious antigens to T cells. While previous pneumonia studies have found associations with both MHC class I and II alleles, ^23^ ^24^ our study finds the strongest HLA associations in class II alleles especially among the youngest subgroup. Our MHC class II findings (DQB1*03:02 and DQA1*03:01), for example, are comparable to those observed among CVID patients ^25^; these results are suggestive of mechanisms involving B cell maturation. Further, our MHC class II (DRB4*01:03 and DRB1*04:03) findings among the youngest subgroup may also indicate that biology of autoimmunity can contribute to pneumonia risk. ^26^ Finally, we conclude that the MHC class II allele associations in our study suggest a potential role for extracellular pathogen antigen presentation to modify pneumonia risk. ^27^

Apolipoprotein E (APOE) alleles ε4 and ε2 are associated with a lower and higher pneumonia risk in our study population, respectively. APOE ε4 is associated with harmful health consequences such as Alzheimer’s disease (AD), hypercholesterolemia and cardiovascular disease. ^28^ While ε4 allele predisposes to human immunodeficiency virus, herpes simplex virus-1 and Covid-19 infections, the ε4 may also protect against several diseases. ApoE protein can function in binding and internalisation of infectious agents and regulation of inflammatory responses; ^29^ it has been suggested that the ε4 can be associated with lower inflammation when compared to ε3 genotype. ^30^ The ε4 is associated with higher fecundity possibly explaining the maintenance of the ε4 in the human population. ^31^ It is also possible that the ε4 protective role on pneumonia may contribute to maintaining the ε4 allele in the population.

Life-style as well as general and lung health may be implicated in airway infection risk. ^3^ Recurrent pneumonia in our study is associated with *CRP* and mucin genes, highlighting the role of inflammation and mucus production. *RIN3,* a gene that contributes to AD, chronic obstructive pulmonary disease and obesity, is also associated with pneumonia. ^15^ We further found that pneumonia risk among asthmatic patients is associated with *FHIT*; this tumor suppressor protein is implicated in lung cancer, regulation of naive T cells and MHC class I cell surface expression. ^18^ ^32^ *CHRNA5*, a gene involved in nicotine dependency ^17^, modifies the pneumonia risk. Smoking triggers proinflammatory IL-1β production, which in turn regulates *MUC5B*-mediated mucin production. ^33^ Importantly, smoking can also disturb the B cells and serum immunoglobulin G concentration. ^34^ These examples highlight the view that combinations of complex genetic mechanisms influenced by life-style, immunity and general health may contribute to pneumonia risk.

Results of our genetic correlation analysis are consistent with pneumonia risk factors identified in earlier observational and genetic correlation studies ^35^ ^10^ Correlated traits, such as smoking and obesity, are well-known risk factors for pneumonia. Lung health status indicated by wheezing or chronic bronchitis, as well as low education, are also risk factors for pneumonia. ^3^ We further found that obesity, low educational level, slow walking and smoking may act as causal exposures to pneumonia. These findings suggest a role for socio-economic status in pneumonia risk while underscoring the significance of addressing smoking and obesity in pneumonia prevention.

The strength of our study lies in its large and accurately defined study populations. Previously, pneumonia GWASs have been conducted in samples with limited size or in cohorts partly based on self-reported data. ^8^ ^10^ ^9^ ^11^ In our study, the pneumonia diagnosis is confirmed at health care, and most patients have been hospitalised. Limitations of our study include the findings related to the *CRP* locus; the individuals carrying high-response *CRP* allele may be assigned pneumonia diagnosis more readily. ^36^ Importantly, our study population was limited to Northern Europeans; similar studies should be completed in additional ethnic and geographical populations.

In summary, we have conducted a pneumonia GWAS in the general population and, for the first time, among specific patient subgroups categorised by age at diagnosis, recurrence of pneumonia, and the presence of asthma. Many of our findings involve the regulation of immunity, including acute inflammation, antigen presentation, and T cell biology. The genetic correlations and causal evidence support the earlier findings and suggest causal roles for obesity, smoking, and lower education level in the development of pneumonia. We conclude that the genetic influence and mechanisms on pneumonia risk differ between patient subgroups, highlighting immunological mechanisms in early life in contrast to lifestyle and lung function mechanisms among the elderly.

## Data Availability

Summary statistics will be made available through the NHGRI-EBI GWAS Catalog upon publication. The Finnish biobank data can be accessed through the Fingenious services (https://site.fingenious.fi/en/) managed by FINBB.

## Acknowledgements

The authors wish to acknowledge CSC – IT Center for Science, Finland, for computational resources. We want to acknowledge the participants and investigators of the FinnGen study. Following biobanks are acknowledged for delivering biobank samples to FinnGen: Auria Biobank (www.auria.fi/biopankki), THL Biobank (www.thl.fi/biobank), Helsinki Biobank (www.helsinginbiopankki.fi), Biobank Borealis of Northern Finland (https://www.ppshp.fi/Tutkimus-ja-opetus/Biopankki/Pages/Biobank-Borealis-briefly-in-English.aspx), Finnish Clinical Biobank Tampere (www.tays.fi/en-US/Research_and_development/Finnish_Clinical_Biobank_Tampere), Biobank of Eastern Finland (www.ita-suomenbiopankki.fi/en), Central Finland Biobank (www.ksshp.fi/fi-FI/Potilaalle/Biopankki), Finnish Red Cross Blood Service Biobank (www.veripalvelu.fi/verenluovutus/biopankkitoiminta), Terveystalo Biobank (www.terveystalo.com/fi/Yritystietoa/Terveystalo-Biopankki/Biopankki/) and Arctic Biobank (https://www.oulu.fi/en/university/faculties-and-units/faculty-medicine/northern-finland-birth-cohorts-and-arctic-biobank). All Finnish Biobanks are members of BBMRI.fi infrastructure (https://www.bbmri-eric.eu/national-nodes/finland/). Finnish Biobank Cooperative -FINBB (https://finbb.fi/) is the coordinator of BBMRI-ERIC operations in Finland. The Finnish biobank data can be accessed through the Fingenious^®^ services (https://site.fingenious.fi/en/) managed by FINBB.

We want to acknowledge the participants of the Estonian Biobank for their contributions. The Estonian Genome Center analyses were partially carried out in the High Performance Computing Center, University of Tartu.

