## Supplementary Appendix for "Exploring genetic risk factors for pneumonia using biobank resources - Insights across subpopulations"

### Supplementary Methods

#### *Study populations*

Life-long data of 64,635 pneumonia hospitalisation episodes among 412,181 participants of FinnGen project was analysed. The FinnGen project combines genetic samples from biobanks and information from Finnish registry data, offering a resource for a wide range of genomic analyses. We used FinnGen data freeze 10 to conduct six different GWAS analyses: general pneumonia (64,635 episodes), pneumonia in age subgroups (under 16 years olds (3,193 pneumonia episodes), 16-60 years olds (24,032 pneumonia episodes) and over 60 years olds (37,410 pneumonia episodes), recurrent pneumonia (at least three separate pneumonia episodes during lifetime, 6,903 cases) and pneumonia among asthma patients (50,929 asthma cases, out of which 14,234 also had pneumonia diagnosis). Further information on the case definition of each analysis, as well as exact amounts of cases and controls can be found from **Supplementary Table 2**.

Patients and control subjects in FinnGen provided informed consent for biobank research, based on the Finnish Biobank Act. Alternatively, separate research cohorts, collected prior the Finnish Biobank Act came into effect (in September 2013) and start of FinnGen (August 2017), were collected based on study-specific consents and later transferred to the Finnish biobanks after approval by Fimea (Finnish Medicines Agency), the National Supervisory Authority for Welfare and Health. Recruitment protocols followed the biobank protocols approved by Fimea. The Coordinating Ethics Committee of the Hospital District of Helsinki and Uusimaa (HUS) statement number for the FinnGen study is Nr HUS/990/2017.

The FinnGen study is approved by Finnish Institute for Health and Welfare (permit numbers: THL/2031/6.02.00/2017, THL/1101/5.05.00/2017, THL/341/6.02.00/2018, THL/2222/6.02.00/2018, THL/283/6.02.00/2019, THL/1721/5.05.00/2019 and THL/1524/5.05.00/2020), Digital and population data service agency (permit numbers:

VRK43431/2017-3, VRK/6909/2018-3, VRK/4415/2019-3), the Social Insurance Institution (permit numbers: KELA 58/522/2017, KELA 131/522/2018, KELA 70/522/2019, KELA 98/522/2019, KELA 134/522/2019, KELA 138/522/2019, KELA 2/522/2020, KELA 16/522/2020), Findata permit numbers THL/2364/14.02/2020, THL/4055/14.06.00/2020, THL/3433/14.06.00/2020, THL/4432/14.06/2020, THL/5189/14.06/2020, THL/5894/14.06.00/2020, THL/6619/14.06.00/2020, THL/209/14.06.00/2021, THL/688/14.06.00/2021, THL/1284/14.06.00/2021, THL/1965/14.06.00/2021, THL/5546/14.02.00/2020, THL/2658/14.06.00/2021, THL/4235/14.06.00/2021, Statistics Finland (permit numbers: TK-53-1041-17 and TK/143/07.03.00/2020 (earlier TK-53-90-20) TK/1735/07.03.00/2021, TK/3112/07.03.00/2021) and Finnish Registry for Kidney Diseases permission/extract from the meeting minutes on 4th July 2019.

The Biobank Access Decisions for FinnGen samples and data utilized in FinnGen Data Freeze 10 include: THL Biobank BB2017_55, BB2017_111, BB2018_19, BB_2018_34, BB_2018_67, BB2018_71, BB2019_7, BB2019_8, BB2019_26, BB2020_1, BB2021_65, Finnish Red Cross Blood Service Biobank 7.12.2017, Helsinki Biobank HUS/359/2017, HUS/248/2020, HUS/150/2022 § 12, §13, §14, §15, §16, §17, §18, and §23, Auria Biobank AB17-5154 and amendment #1 (August 17 2020) and amendments BB_2021-0140, BB_2021-0156 (August 26 2021, Feb 2 2022), BB_2021-0169, BB_2021-0179, BB_2021-0161, AB20-5926 and amendment #1 (April 23 2020)and it´s modification (Sep 22 2021), Biobank Borealis of Northern Finland_2017_1013, 2021_5010, 2021_5018, 2021_5015, 2021_5023, 2021_5017, 2022_6001, Biobank of Eastern Finland 1186/2018 and amendment 22 § /2020, 53§/2021, 13§/2022, 14§/2022, 15§/2022, Finnish Clinical Biobank Tampere MH0004 and amendments (21.02.2020 & 06.10.2020), §8/2021, §9/2022, §10/2022, §12/2022, §20/2022, §21/2022, §22/2022, §23/2022, Central Finland Biobank 1-2017, and Terveystalo Biobank STB 2018001 and amendment 25th Aug 2020, Finnish

Hematological Registry and Clinical Biobank decision 18th June 2021, Arctic biobank P0844: ARC_2021_1001.

The activities of the EstBB are regulated by the Human Genes Research Act. Individual level data analysis in EstBB was carried out under ethical approval 1.1-12/624 from the Estonian Committee on Bioethics and Human Research (Estonian Ministry of Social Affairs), using data according to release application 6-7/GI/33501 from the Estonian Biobank.

***Estonian Biobank***

The Estonian Biobank (EstBB) is a population-based biobank with around 200k participants. All biobank participants have signed a broad informed consent form and information on ICD-10 codes is obtained via regular linking with the national Health Insurance Fund and other relevant databases, with majority of the electronic health records having been collected since 2004.[1]

**Pneumonia definition**

In “unselected pneumonia” group, cases were defined as individuals with pneumonia diagnosis at least once during lifetime. Controls were individuals without pneumonia diagnosis. There were no control exclusions.

In the age group of under 16 year olds, cases were individuals diagnosed with pneumonia for the first time during lifetime at under 16 years of age. We excluded individuals that were diagnosed with pneumonia for the first time during lifetime when over 16 years old from this analysis. Controls were individuals without pneumonia diagnosis.

In the age group of 16-60 year olds, cases were individuals diagnosed with pneumonia at 16-60 years of age for the first time during lifetime. We excluded individuals that were diagnosed with pneumonia for the first time during lifetime when either under 16 years old or over 60 years old. Controls were individuals without pneumonia diagnosis.

In the age group of over 60 year olds, cases were individuals diagnosed with pneumonia at over 60 years of age for the first time during lifetime. We excluded individuals that were diagnosed with pneumonia for the first time when under 60 years old. Controls were individuals without pneumonia diagnosis.

For “recurrent pneumonia” group, cases were defined as individuals who had had 3 or more separate pneumonia episodes (we considered episodes separate if there was at least 3 months (90 days) between diagnoses). Controls were individuals wihtout pneumonia diagnosis.

In the asthma group, we considered cases as individuals with both pneumonia and asthma diagnosis. Controls were individuals with asthma diagnosis, but without pneumonia diagnosis. Individuals with no asthma diagnosis were excluded from this analysis.

ICD-codes used for pneumonia diagnosis:

ICD-10: J10.0, J11.0, J12, J12.0, J12.1, J12.2, J12.3, J12.8, J12.9, J13, J14, J15.0, J15.1, J15.2, J15.3, J15.4, J15.5, J15.6, J15.7, J15.8, J15.9, J16.0, J16.8, J18.0, J18.1, J18.2, J18.8, J18.9

ICD-9: 480, 481, 482, 483, 485, 4822, 4810A, 4820A, 4821A, 4822A, 4823A, 4824A, 4828X, 4870A

ICD-8: 482, 486, 47101, 47109, 48099, 48199, 48201, 48210, 48220, 48230, 48298, 48399, 48502, 48509

ICD-codes used for asthma diagnosis:

ICD-10: J45.0, J45.1, J45.8, J45.9, J46

ICD-9: 493

ICD-8: 493

***Genotyping, imputation, and quality control***

FinnGen samples were genotyped using Illumina and Affymetrix arrays (Illumina Inc., San Diego and Thermo Fischer Scientific, Santa Clara, CA, USA). Sample quality control (QC) was conducted to exclude individuals with genotype missingness of over 5 %, ambiguous gender, excess heterozygosity (±4 SDs), and non-Finnish ancestry. In variant QC, variants with low Hardy-Weinberg equilibrium (p < 1 * 10-6), over 2 % missingness or minor allele count under 3 %, were excluded.. Eagle 2.3.5 [2] was used for pre-phasing. Imputation was conducted with Beagle 4.1. [3], using Finnish population specific SISu v4.2 reference panel. In post-imputation QC, imputation info threshold of < 0.6 was used.

All EstBB participants have been genotyped at the Core Genotyping Lab of the Institute of Genomics, University of Tartu, using Illumina Global Screening Array v1.0 and v2.0. Samples were genotyped and PLINK format files were created using Illumina

GenomeStudio v2.0.4. Individuals were excluded from the analysis if their call-rate was <95% or if sex defined based on heterozygosity of X chromosome did not match sex in phenotype data. Before imputation, variants were filtered by call-rate <95%, HWE p value < 1e-4 (autosomal variants only), and minor allele frequency <1%. Variant positions were updated to b37 and all variants were changed to be from TOP strand using GSAMD-24v1-0_20011747_A1-b37.strand.RefAlt.zip files from https://www.well.ox.ac.uk/~wrayner/strand/ webpage. Prephasing was done using Eagle v2.3 software[2] (number of conditioning haplotypes Eagle[4] uses when phasing each sample was set to:–Kpbwt=20000) and imputation was done using Beagle v.28Sep18.79339[4] with effective population size ne = 20,000. Population specific imputation reference of 2297 WGS samples was used.[5] (Genotype and imputation information from this article[6]).

***GWAS and meta-analysis***

Each pneumonia GWAS in FinnGen was performed using regenie software [7]. The association model was adjusted for age, sex, the first 10 genetic principal components, and genotyping batch. In EstBB, GWAS was performed using REGENIE 2.24 (https://rgcgithub.github.io/regenie/), using sex, age, age_sq and ten PCs as covariates.

Meta-analysis resulted in total 91,062 cases with general pneumonia, 6,641 cases with pneumonia as under 16 year old, 40,436 cases with pneumonia as 16-60 years old, 44,053 cases with pneumonia as over 60 years old, 8,199 cases with recurrent pneumonia and 19,520 cases of asthma patients with pneumonia. Further information on pneumonia and subgroup definitions, and medical codes used can be found in Supplementary Table 1.

For meta-analysis, EstBB summary statistics were lifted from hg19 to hg38 build. We used METAL [8] software for performing inverse-variance weighted meta-analysis. Cochran's Q-test was performed to estimate heterogeneity.

***Characterization of association signals***

Candidate genes were annotated with the help of previous literature and databases, such as GenBank [9] and UniProt [10]. We considered the association novel, if previous pneumonia associations in a ±1 MB window from the lead variant could not be found on GWAS Catalog [11] .

To estimate the SNP-based heritability and genomic inflation factor, we used the linkage disequilibrium score (LDSC) pipeline tool [12].

**Colocalizations**

We used the coloc R library [13,14] and its 'coloc.abf'-function to estimate colocalizations between Genotype-Tissue Expression Project (GTEx) [15] expression quantitative trait loci (eQTLs) data (V8) in different tissues and pneumonia-associated signals. Under single causal variant assumption, this method calculates the posterior probability that two tested traits share a single causal variant, and, therefore, are likely to share a causal mechanism. We utilized the following eQTL tissues relevant for pneumonia: EBV-transformed lymphocytes, Lung and Whole Blood. Colocalizations with the posterior probability (PP4) over 0.8 were considered significant.

***HLA imputation and associations***

In FinnGen, HLA imputation was carried out using HIBAG R library [16] , employing imputation models trained on Finnish individuals. [17] The imputation was based on a set of SNPs directly genotyped on the FinnGen array. The associations between HLA alleles and pneumonia endpoints were tested using regenie software [7], employing the same covariate structure as the main analysis.

***APOE associations***

*APOE* alleles were determined based on the combination of rs429358 and rs7412 variants as detailed in **Supplementary Table 10** below:

| ***APOE* alleles** | ***rs429358 genotype*** | ***rs7412 genotype*** |
|---|---|---|
| ε4ε4 | C/C | C/C |
| ε3ε4 | T/C | C/C |
| ε2ε4* | T/C | T/C |
| ε3ε3 | T/T | C/C |
| ε2ε3 | T/T | T/C |
| ε2ε2 | T/T | T/T |
| ε1ε3* | T/C | T/C |
| ε1ε4** | C/C | T/C |
| ε1ε2** | C/T | T/T |

*) Ambiguous alleles were coded as ε2ε4 due to the rarity of ε1 allele. **) Extremely rare, omitted from analyses.

We tested for the associations between *APOE* alleles and pneumonia endpoints, as well as Alzheimer's disease (FinnGen endpoint G6_ALZHEIMER; ICD-10: G30), using it as a positive control since *APOE* ε4 is a well-known risk factor for Alzheimer's disease. The associations were tested in R using logistic regression, adjusting for the same covariates as in the primary analysis. We conducted two sets of association tests: first, each pneumonia endpoint and Alzheimer's disease were tested against APOE alleles as shown in **Table S5**, with ε3ε3 as the reference; second, we assessed the effects of the number of copies of each allele (i.e., zero, one or two copies of each ε2, ε3, and ε4) to obtain per allele effect estimate. APOE allele association tests were conducted in a subset of unrelated individuals in FinnGen.

#### *Genetic correlations*

LDSC regression [12] was further used to estimate the genetic correlations between pneumonia and other traits. We tested the correlations of pneumonia with 772 phenotypes available in the in-house LDSC analysis pipeline.

#### *Mendelian randomization*

"TwoSampleMR" R library [18] was used to perform a bi-directional two-sample Mendelian randomization between pneumonia and a selection of traits found to be genetically correlated with pneumonia. Genetic instruments were extracted from the meta-analysis summary statistics. For other traits, we used mostly UKBB-based data provided by the Medical Research Council Integrative Epidemiology Unit (MRC IEU) database implemented in the TwoSampleMR. LD clumping was performed with "clump_data" function. We used its default values (r2=0.001, a clumping window of 10 kb, European population reference). The inverse-variance weighted method (IVW) was considered as the primary analysis. Egger intercepts were used to evaluate horizontal pleiotropy. Additionally, we tested heterogeneity with Cochrans Q test ("mr_heterogeneity") and performed leave-one-out analysis ("mr_leaveoneout"). Causal effects were considered significant if MR Egger estimates acquired in the sensitivity analyses were in a matching direction with the IVW-estimates, false-discovery rate (FDR)-corrected p-value was under 0.05, and if there was no significant pleiotropy.

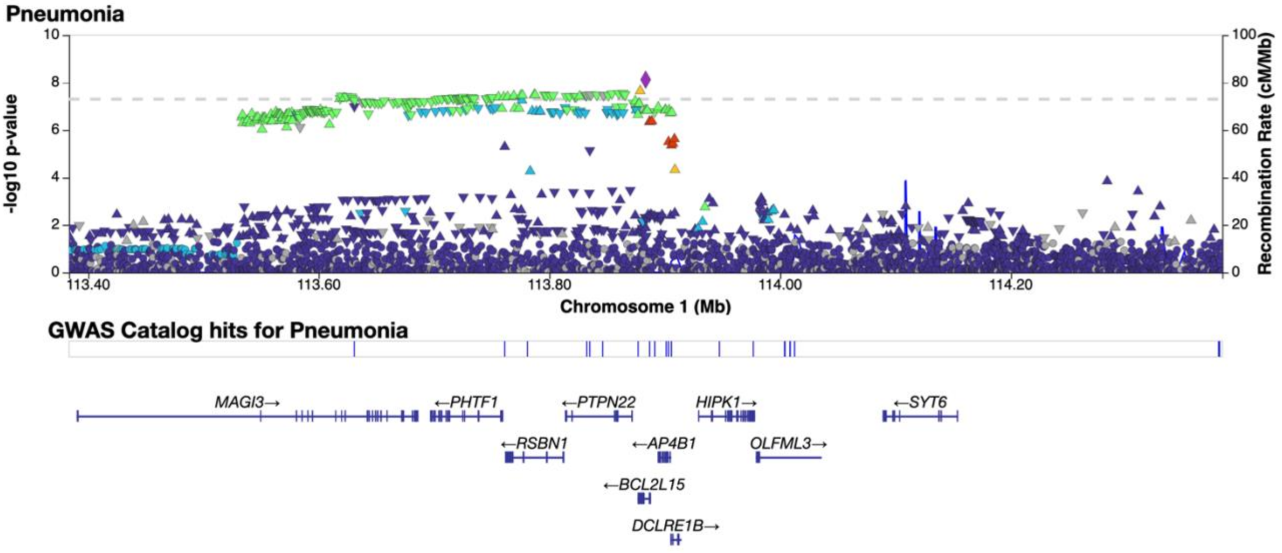

**Figure S1. Regional association plot of the unselected pneumonia risk loci at 1p13.2 (lead variant rs11102694).**

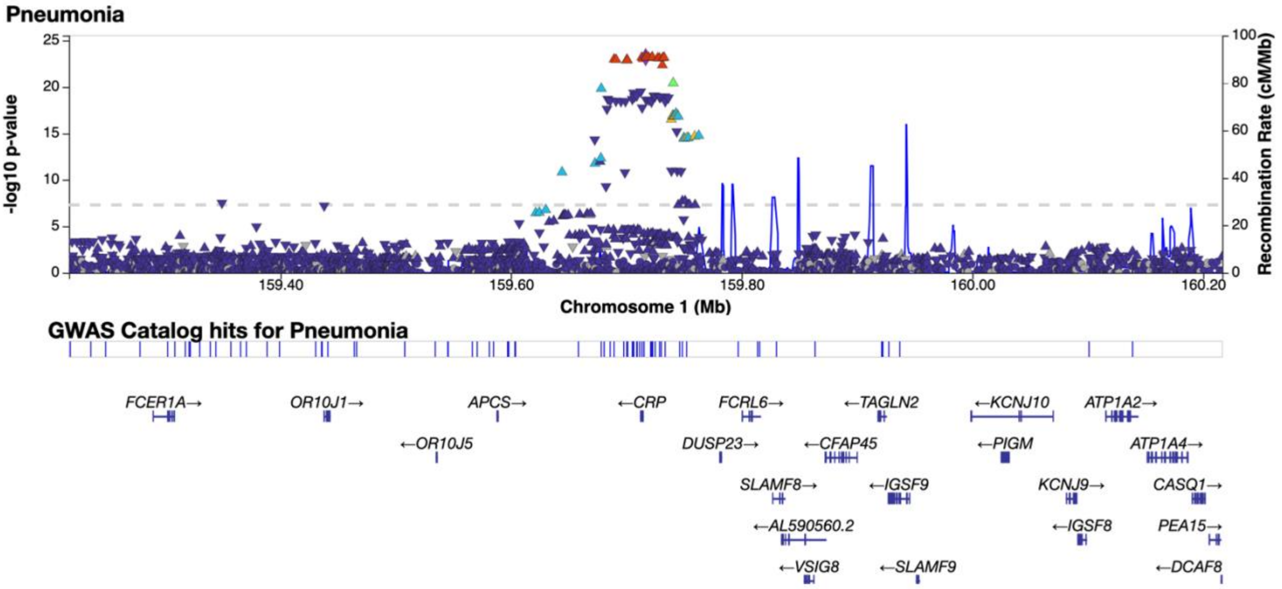

**Figure S2. Regional association plot of the unselected pneumonia risk loci at 1q23.2 (lead variant rs3116636).**

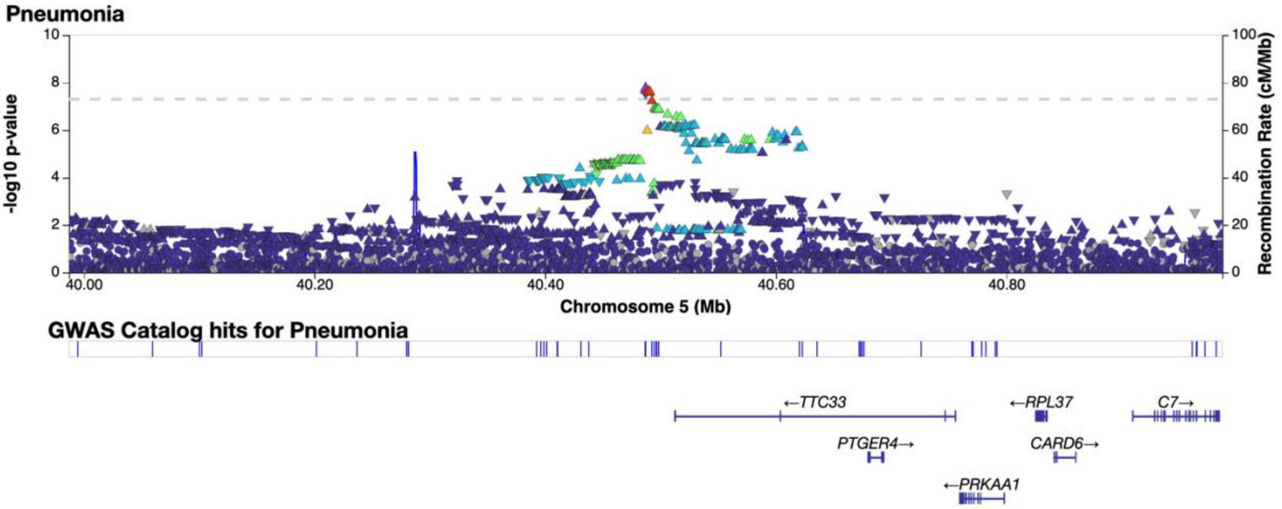

**Figure S3. Regional association plot of the unselected pneumonia risk loci at 5p13.1 (lead variant rs7725052).**

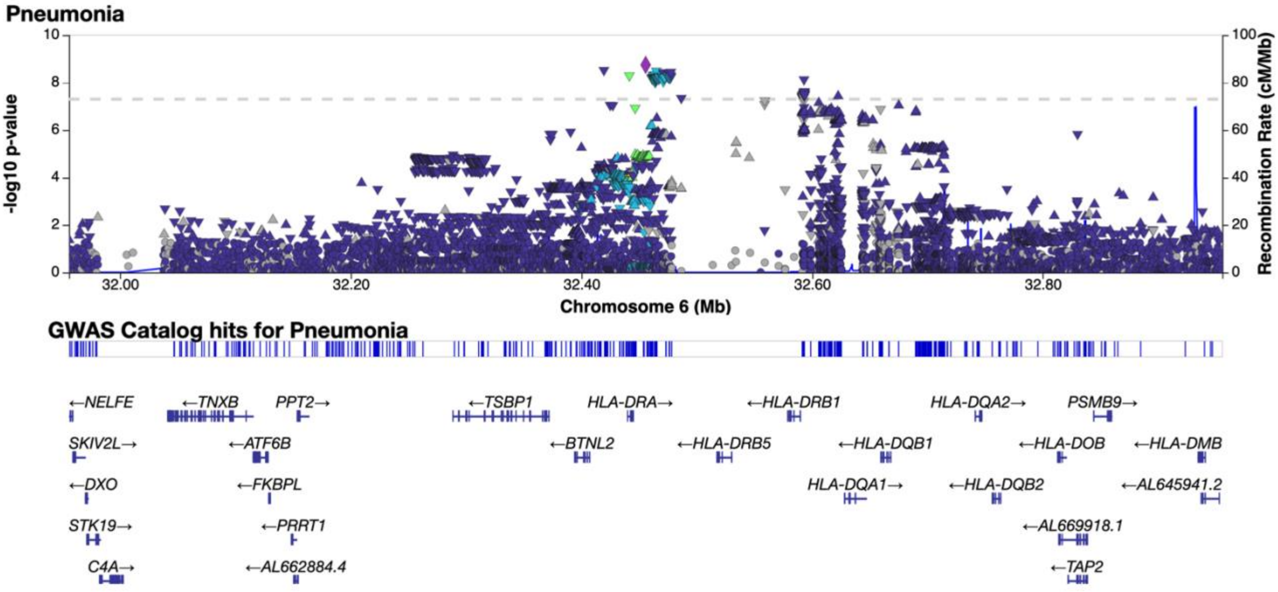

**Figure S4. Regional association plot of the unselected pneumonia risk loci at 6p21.32 (lead variant rs3135369).**

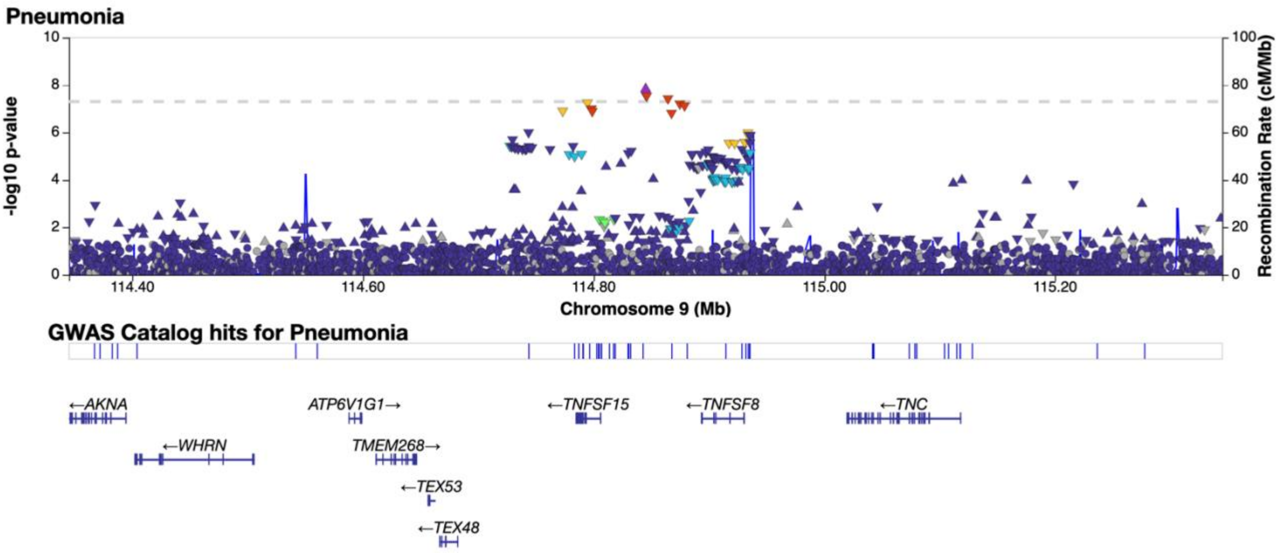

**Figure S5. Regional association plot of the unselected pneumonia risk loci at 9q32 (lead variant rs112284124).**

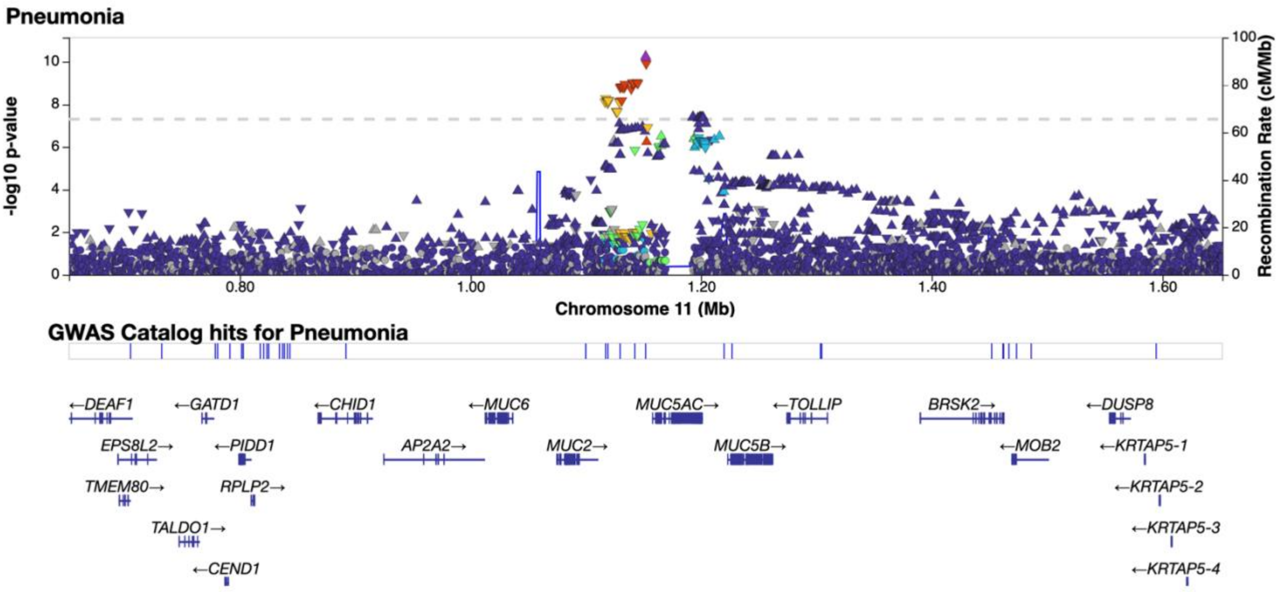

**Figure S6. Regional association plot of the unselected pneumonia risk loci at 11p15.5 (lead variant rs28415845).**

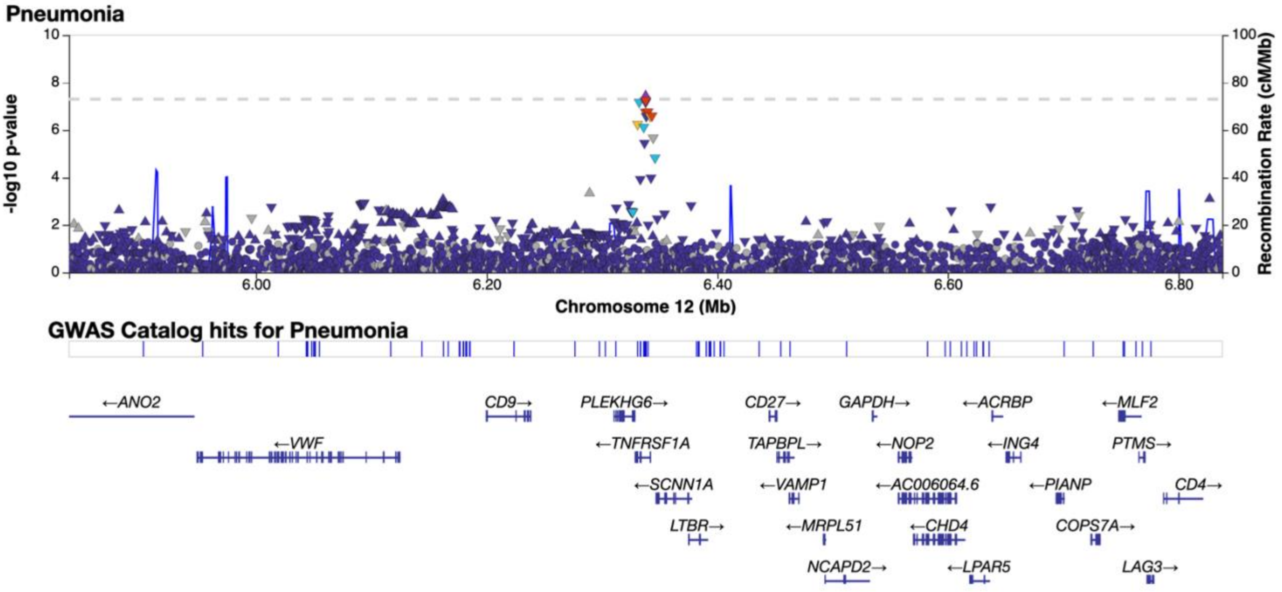

**Figure S7. Regional association plot of the unselected pneumonia risk loci at 12p13.31 (lead variant rs4149581).**

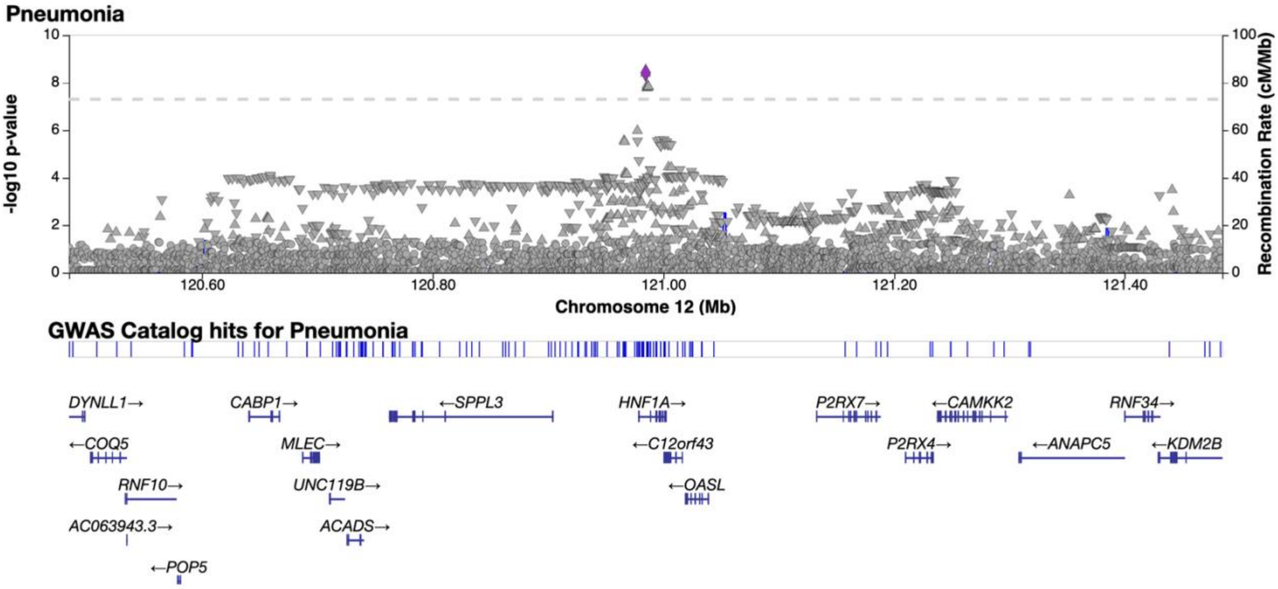

**Figure S8. Regional association plot of the unselected pneumonia risk loci at 12q24.31 (lead variant rs58367757).**

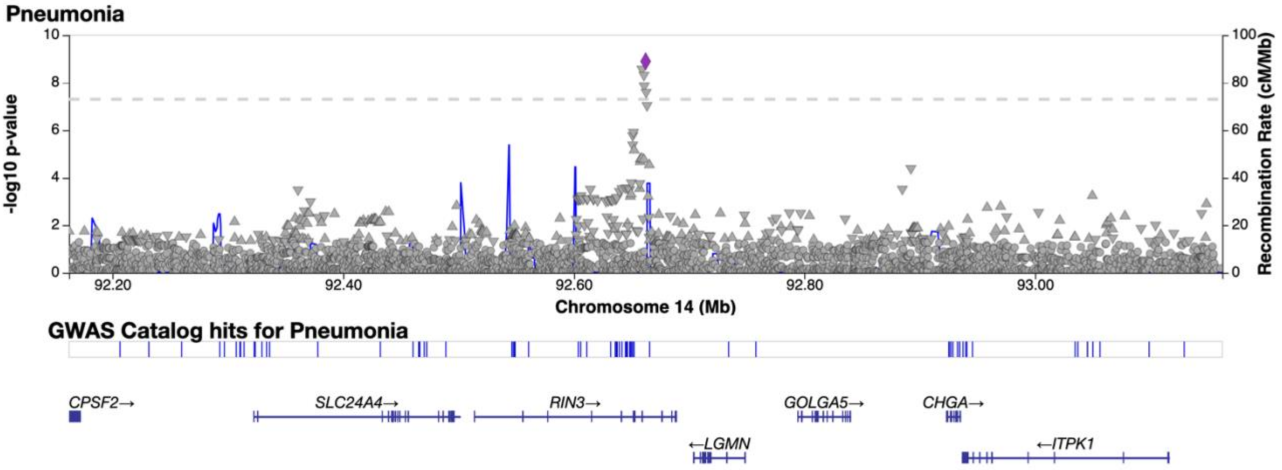

**Figure S9. Regional association plot of the unselected pneumonia risk loci at 14q32.12 (lead variant rs59985723).**

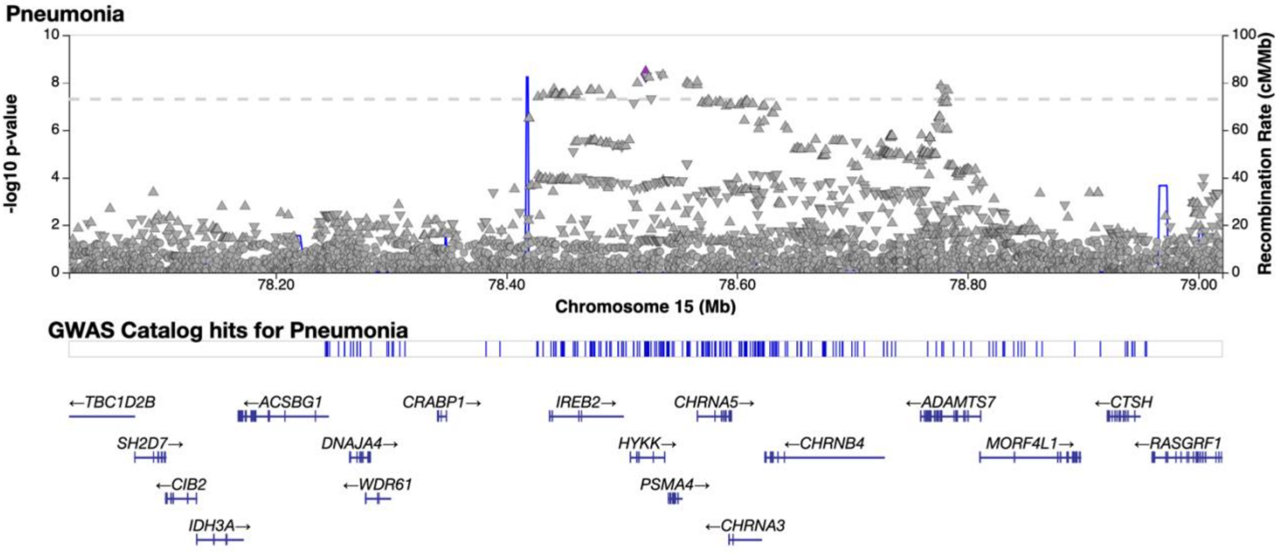

**Figure S10. Regional association plot of the unselected pneumonia risk loci at 15q25.1 (lead variant rs34684276 (previously known as rs113858005)).**

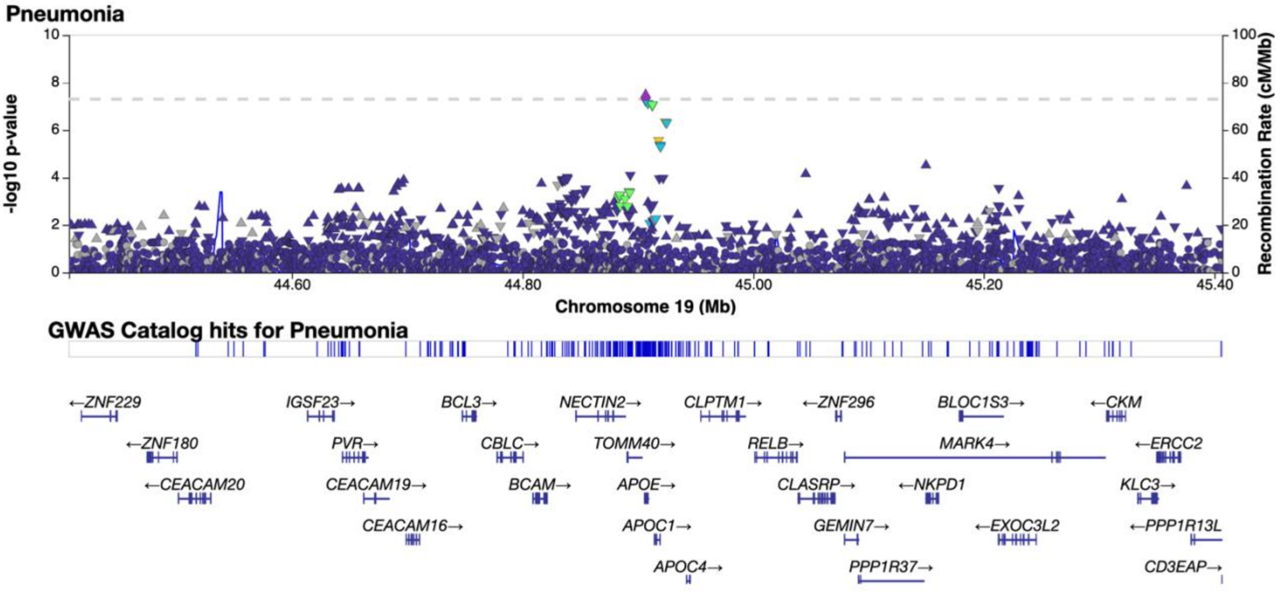

**Figure S11. Regional association plot of the unselected pneumonia risk loci at 19q13.32 (lead variant rs769449).**

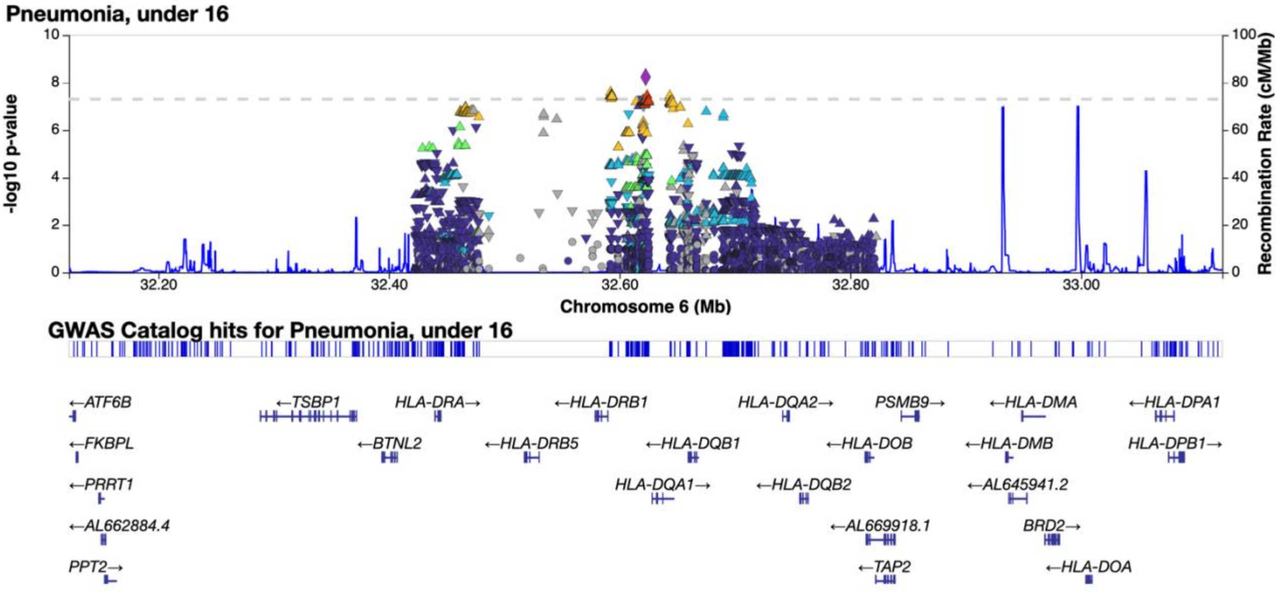

**Figure S12. Regional association plot of pneumonia in under 16 year olds risk loci at 6p21.32 (lead variant rs9271554).**

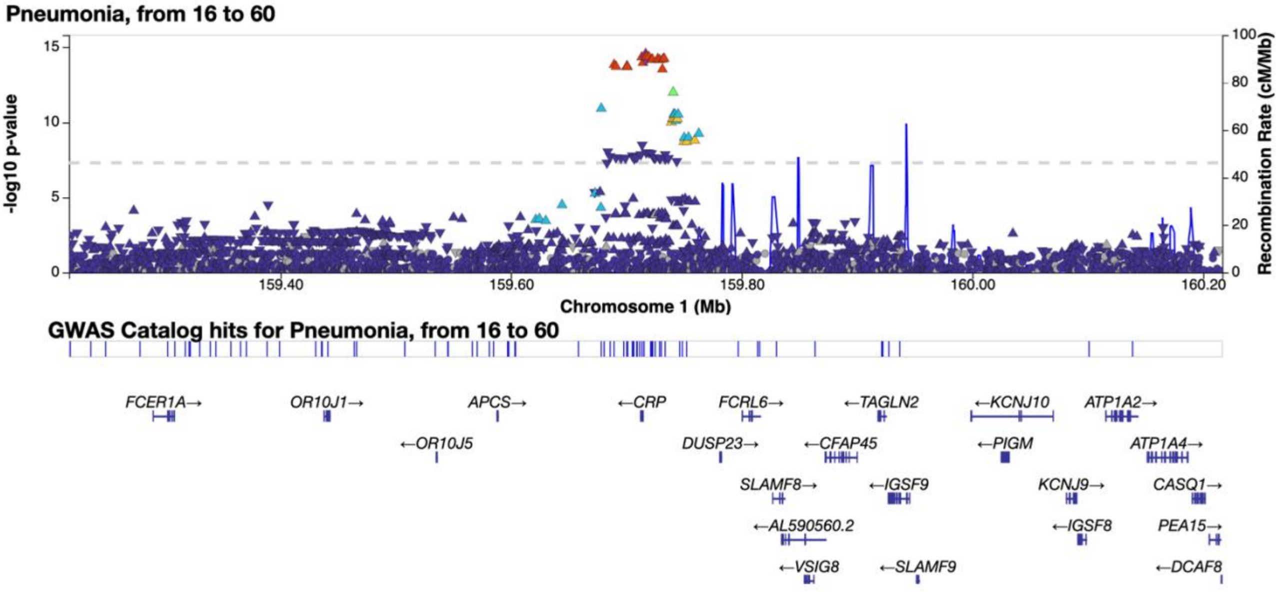

**Figure S13. Regional association plot of pneumonia in 16-60 year olds risk loci at 1q23.2 (lead variant rs3116636).**

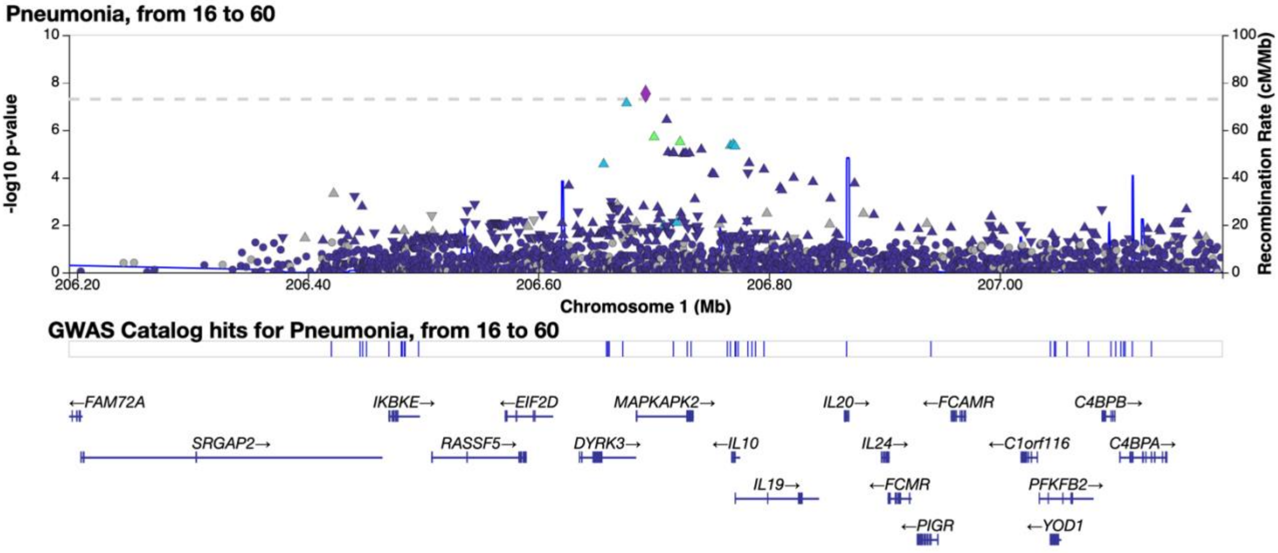

**Figure S14. Regional association plot of pneumonia in 16-60 year olds risk loci at 1q32.1 (lead variant rs61815610).**

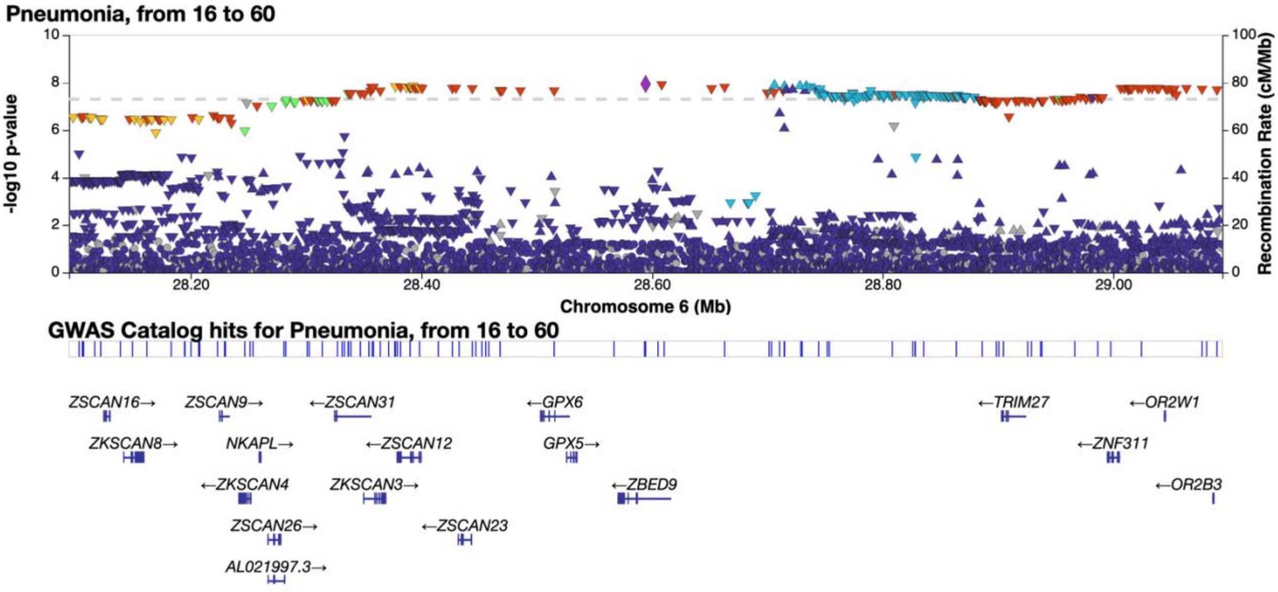

**Figure S15. Regional association plot of pneumonia in 16-60 year olds risk loci at 6p22.1 (lead variant rs34119086).**

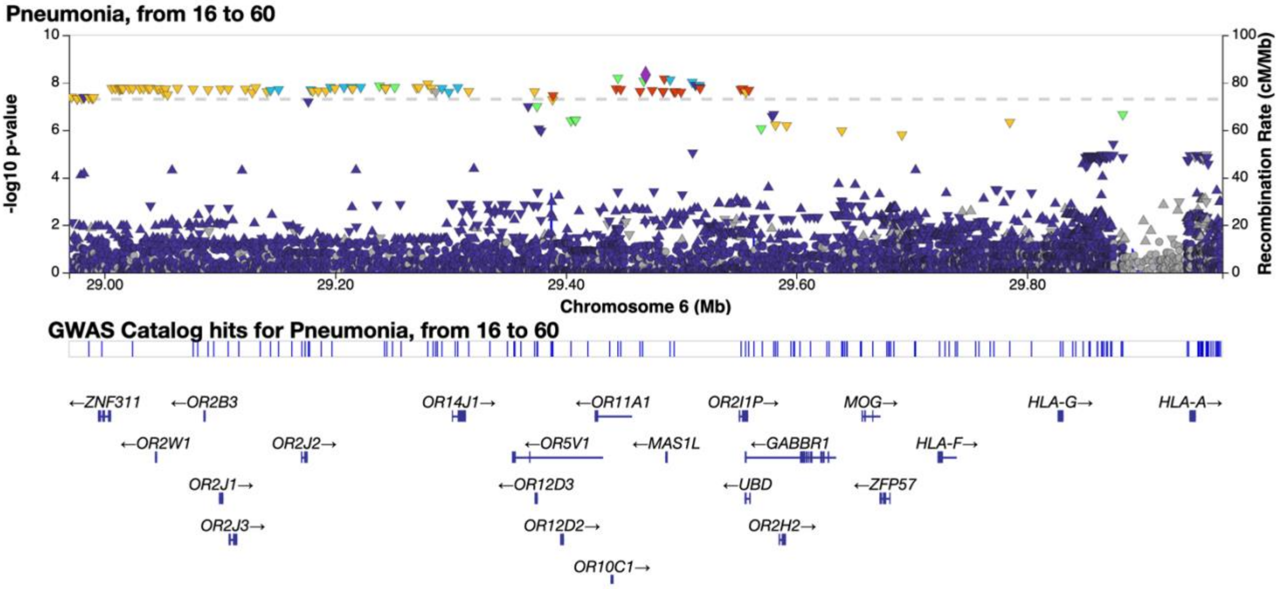

**Figure S16. Regional association plot of pneumonia in 16-60 year olds risk loci at 6p22.1 (lead variant rs2206853).**

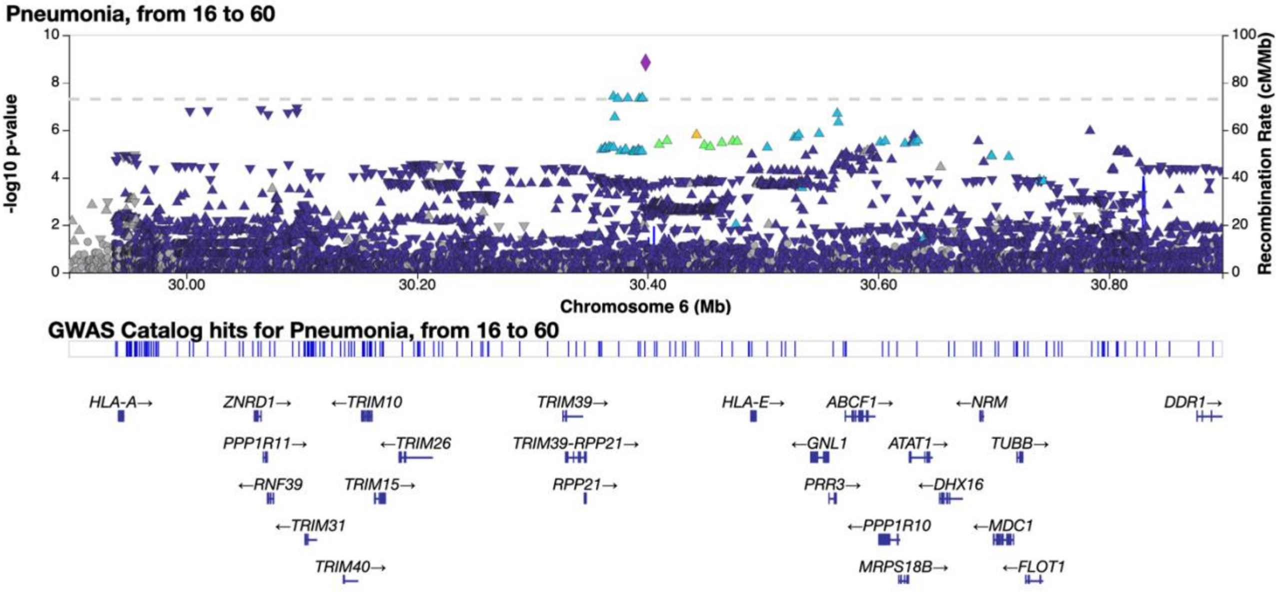

**Figure S17. Regional association plot of pneumonia in 16-60 year olds risk loci at 6p22.1 (lead variant rs1264567).**

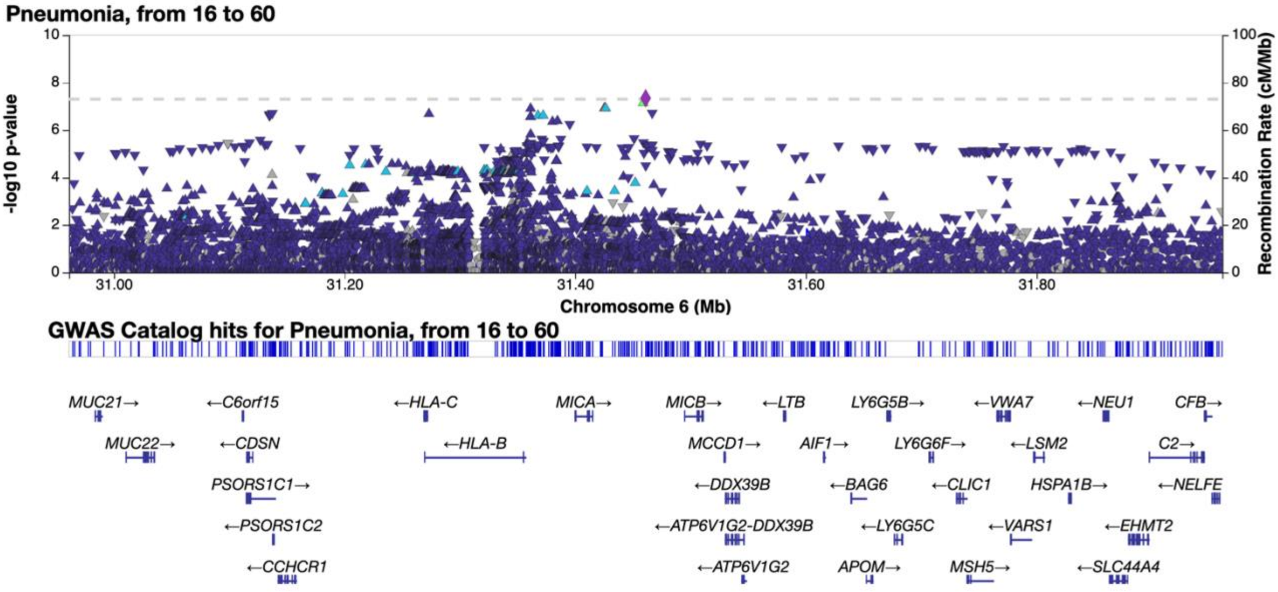

**Figure S18. Regional association plot of pneumonia in 16-60 year olds risk loci at 6p21.33 (lead variant rs139662024).**

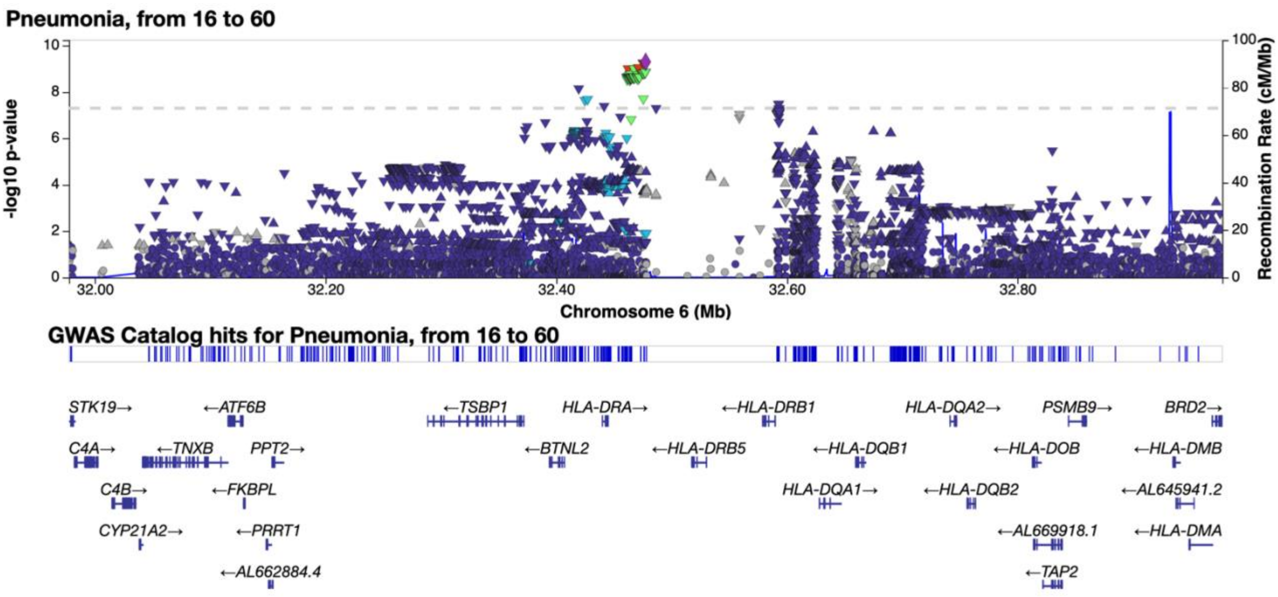

**Figure S19. Regional association plot of pneumonia in 16-60 year olds risk loci at 6p21.32 (lead variant rs9269140).**

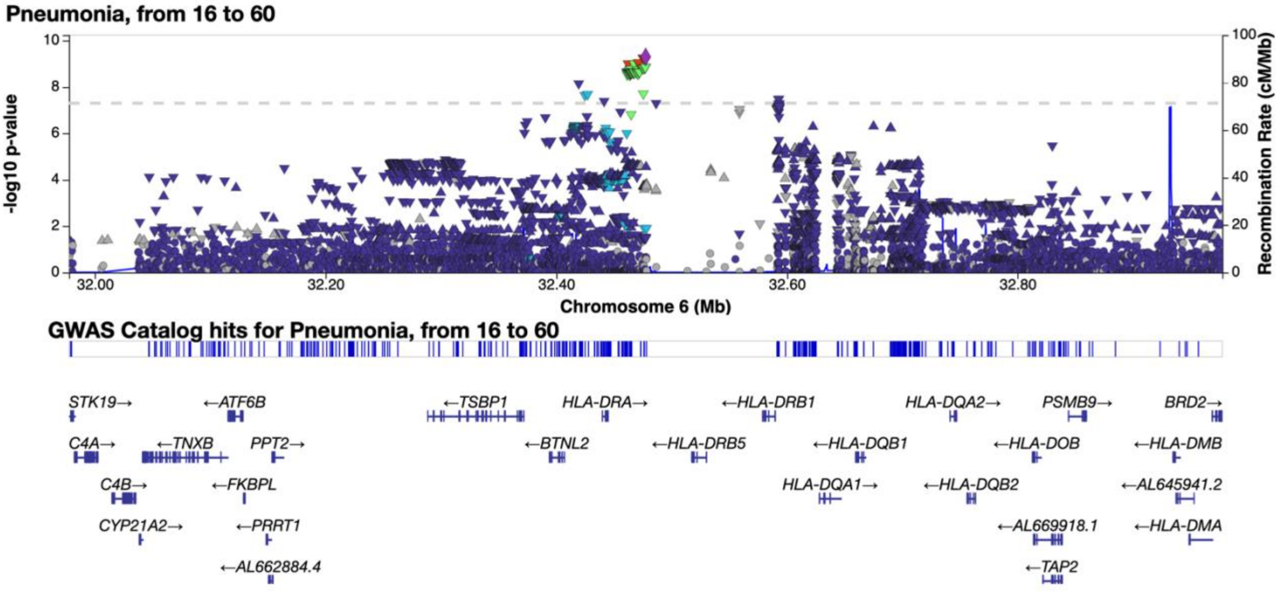

**Figure S20. Regional association plot of pneumonia in 16-60 year olds risk loci at 14q32.12 (lead variant rs2295991).**

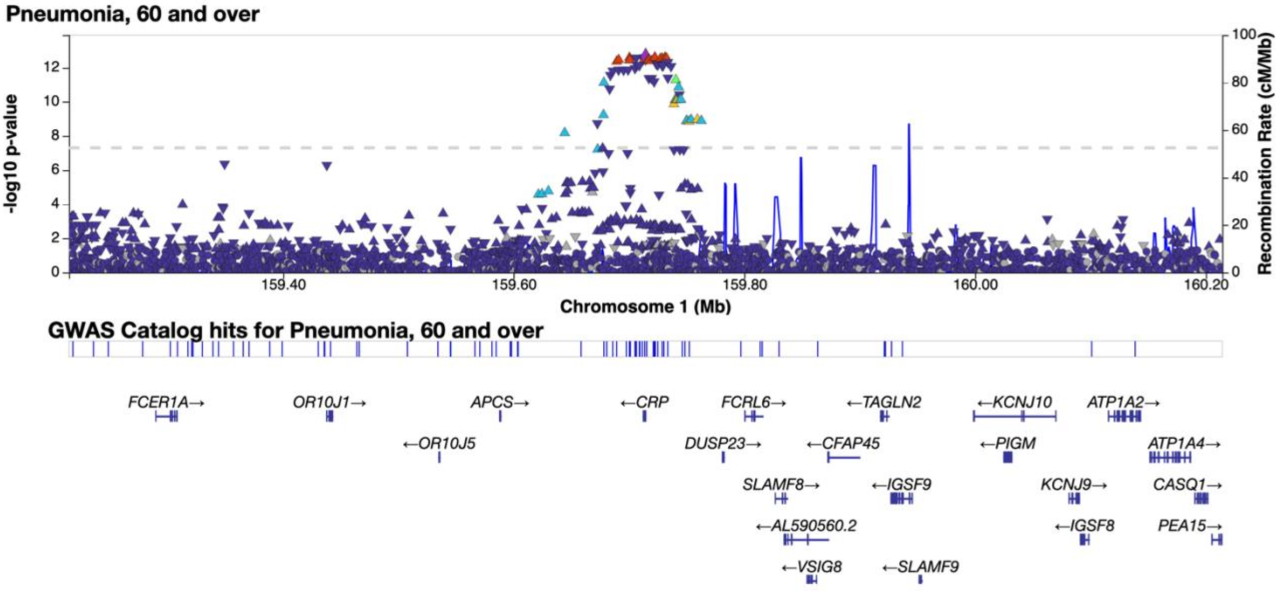

**Figure S21. Regional association plot of pneumonia in over 60 year olds risk loci at 1q23.2 (lead variant rs3116656).**

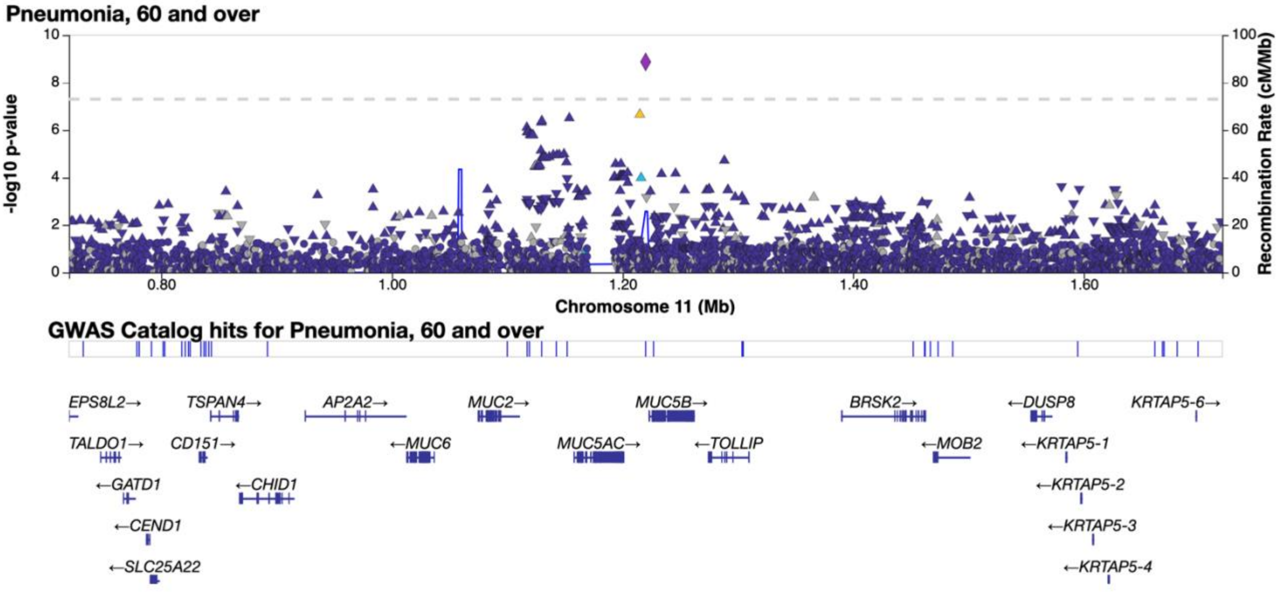

**Figure S22. Regional association plot of pneumonia in over 60 year olds risk loci at 11p15.5 (lead variant rs35705950).**

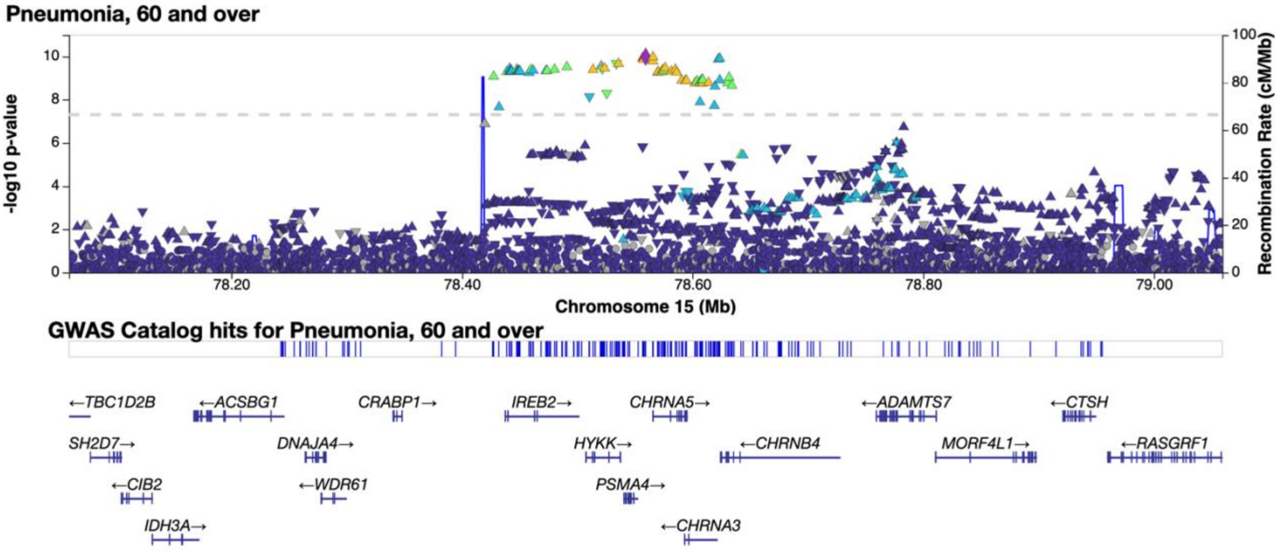

**Figure S23. Regional association plot of pneumonia in over 60 year olds risk loci at 15q25.1 (lead variant rs2036527).**

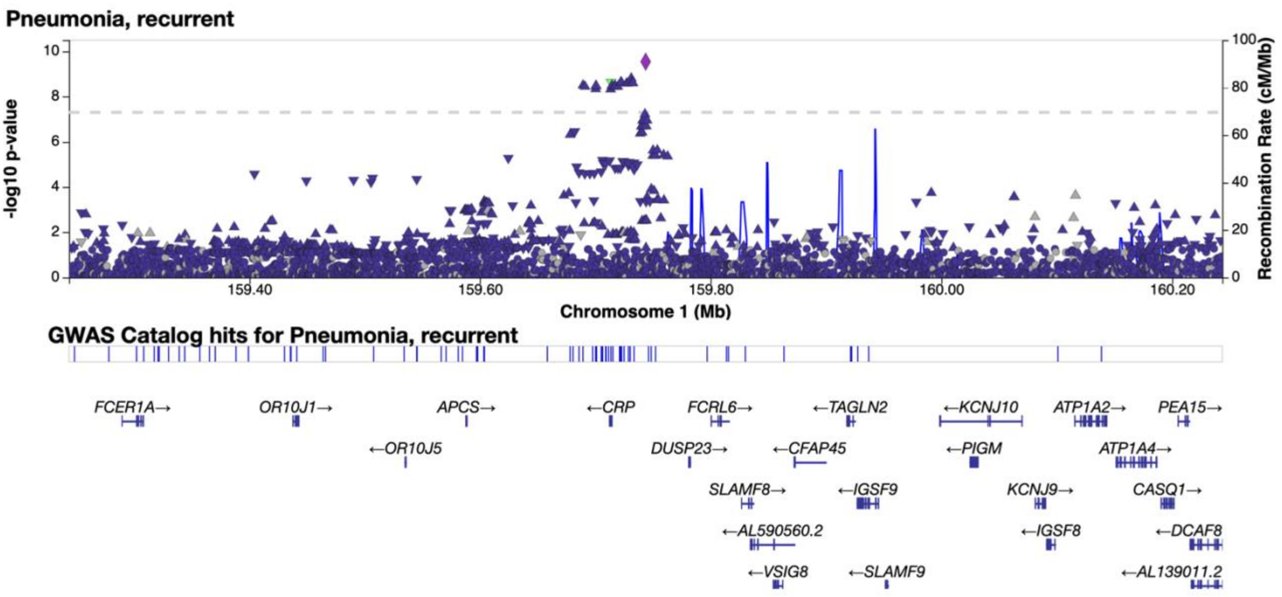

**Figure S24. Regional association plot of recurrent pneumonia risk loci at 1q23.2 (lead variant rs74596724).**

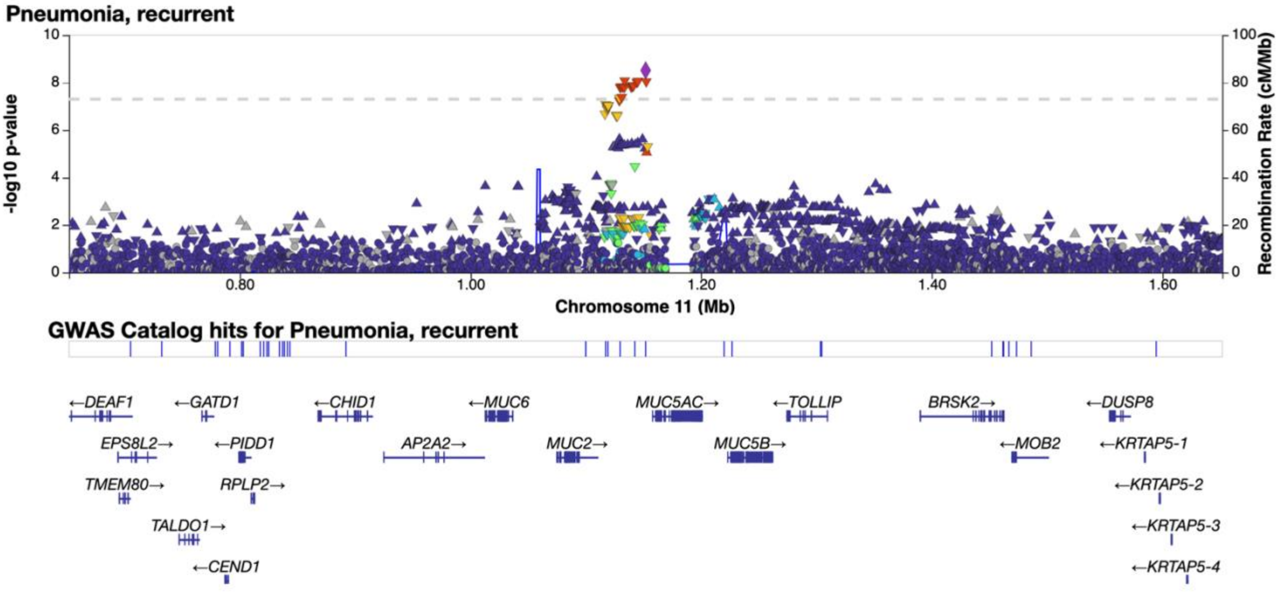

**Figure S25. Regional association plot of recurrent pneumonia risk loci at 11p15.5 (lead variant 11p15.5).**

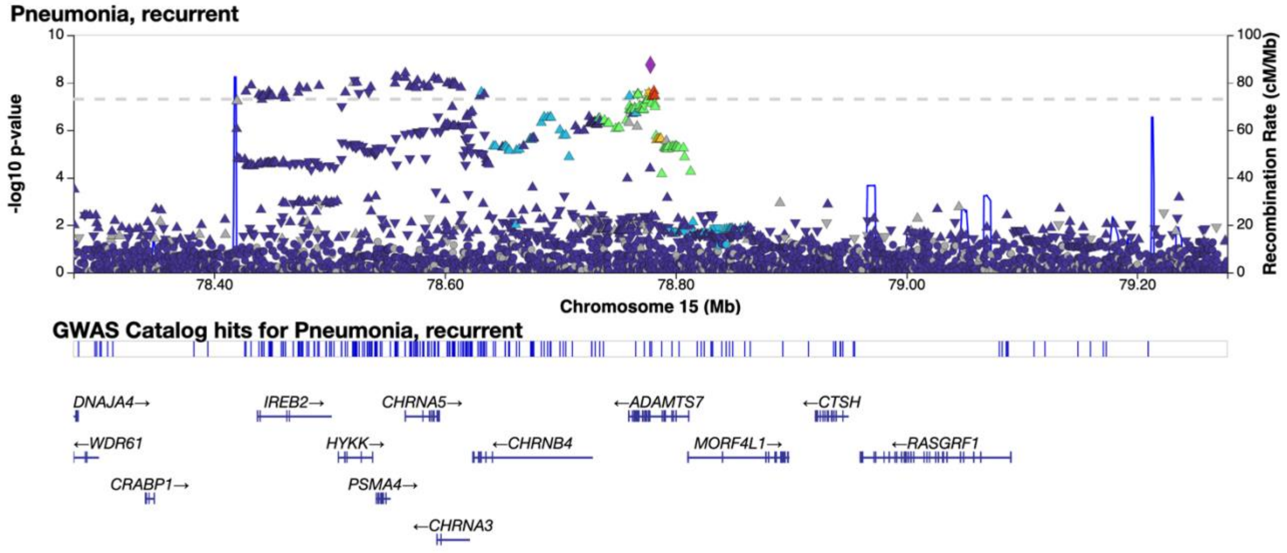

**Figure S26. Regional association plot of recurrent pneumonia risk loci at 15q25.1 (lead variant rs12907764).**

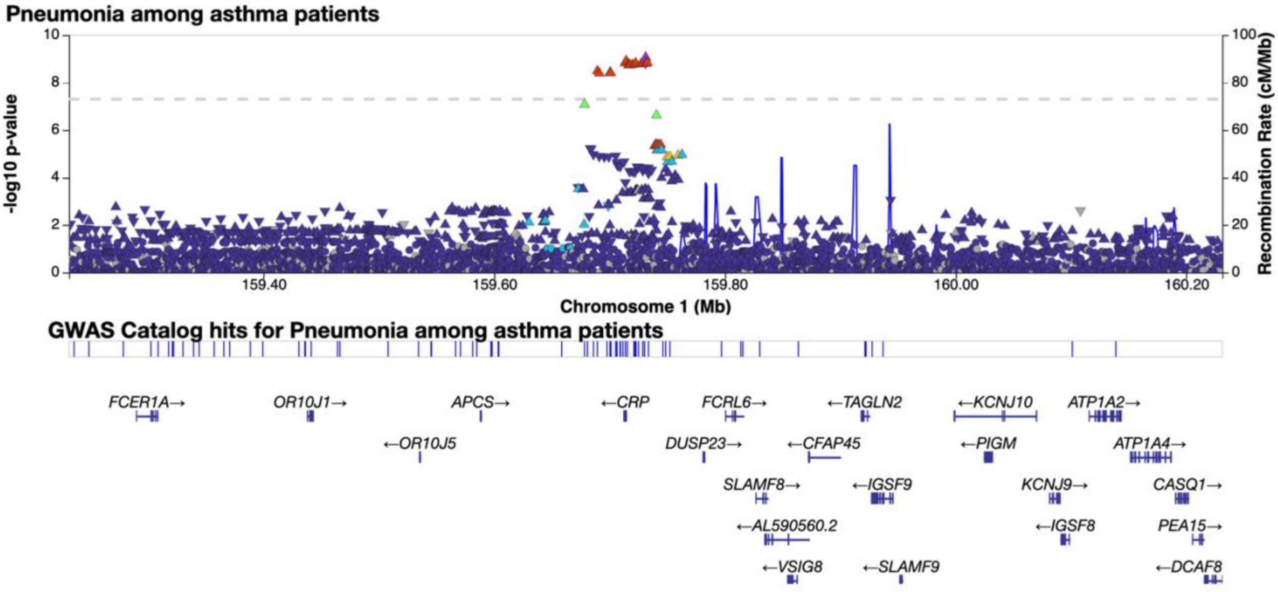

**Figure S27. Regional association plot of pneumonia among asthma patients risk loci at 1q23.2 (lead variant rs4261114).**

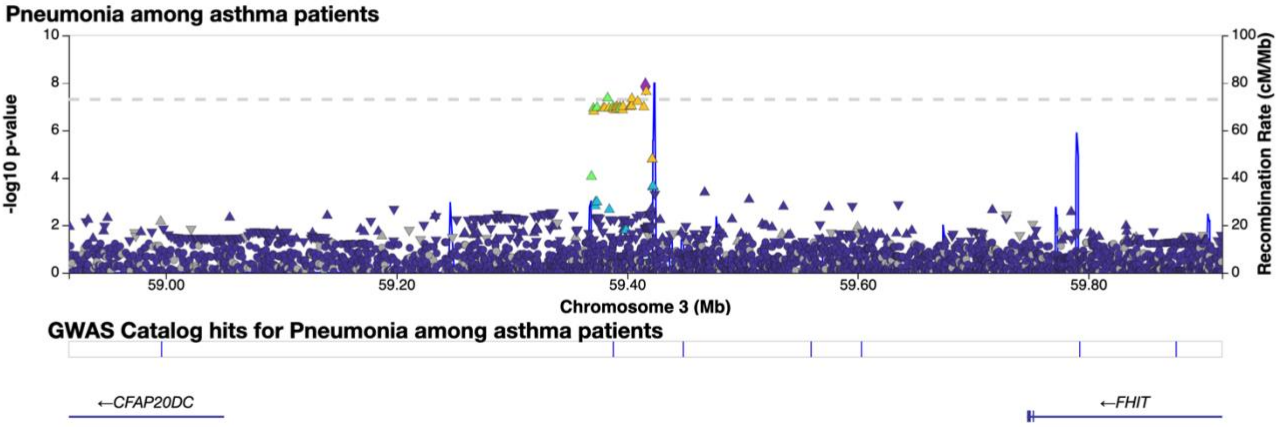

**Figure S28. Regional association plot of pneumonia among asthma patients risk loci at 3:59415853 (lead variant rs11130706).**

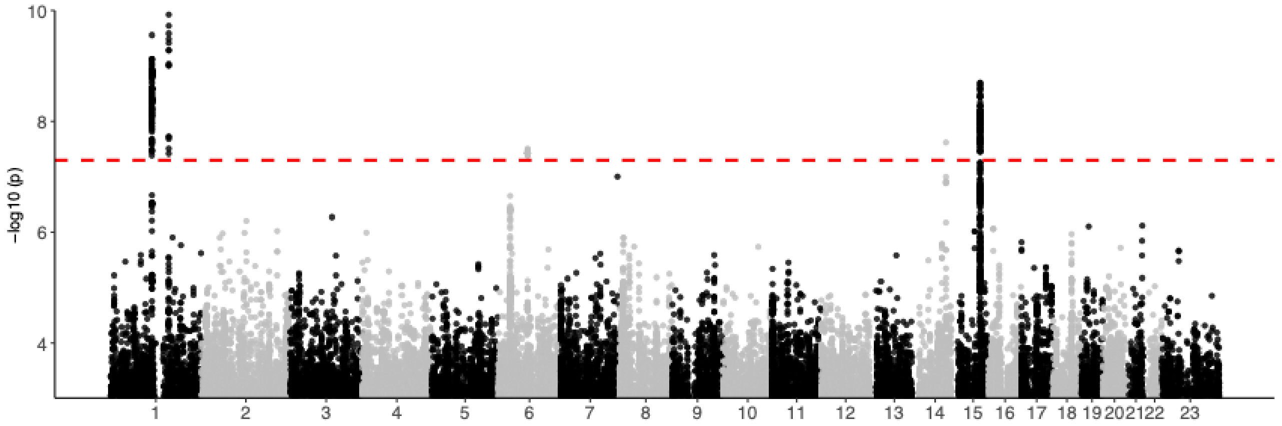

**Figure S29. Manhattan plot of the pneumonia inpatient analysis in FinnGen.** Y-axis represents the -log10 p-value. The genome-wide significance threshold is marked with black dashed horizontal line.

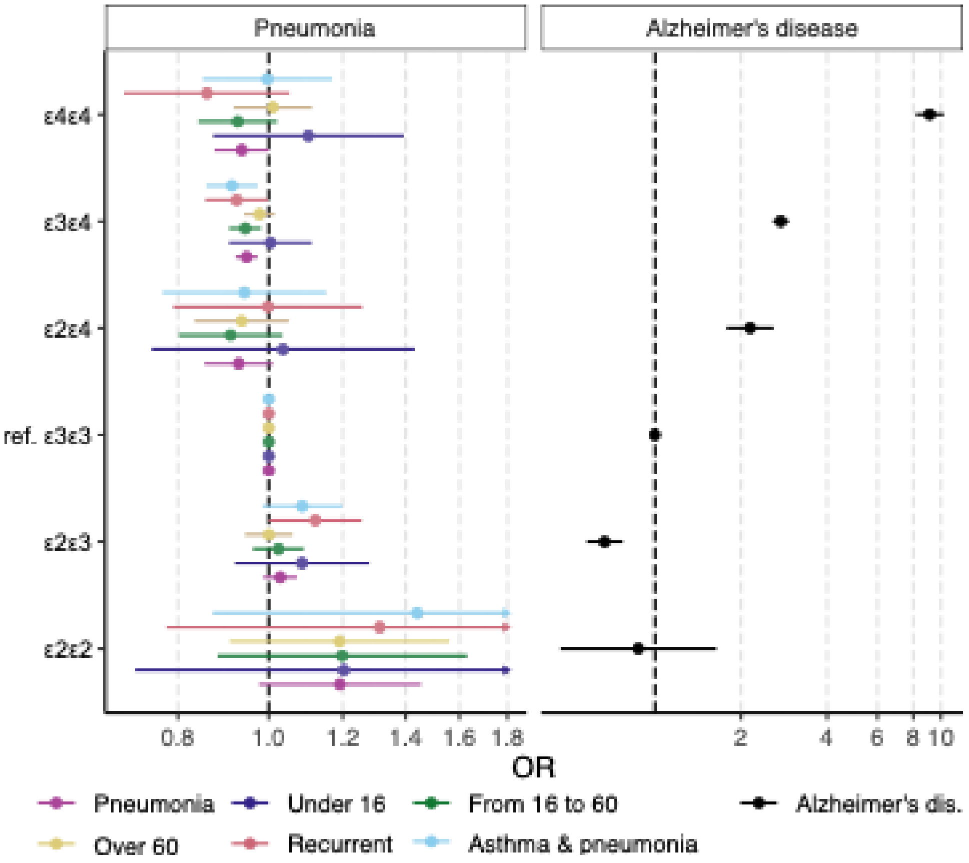

**Figure S30. Effect estimates of pneumonia subgroups in full APOE genotypes in FinnGen.** Left panel shows effect estimates in pneumonia groups in FinnGen population, right shows effect estimates for same alleles among FinnGen Alzheimer's disease cases for comparison. Y-axis shows APOE genotypes and x-axis shows odds ratio for the estimates. Vertical dashed line shows the place of value 1. Legend below the figure shows color coding for pneumonia subgroups.

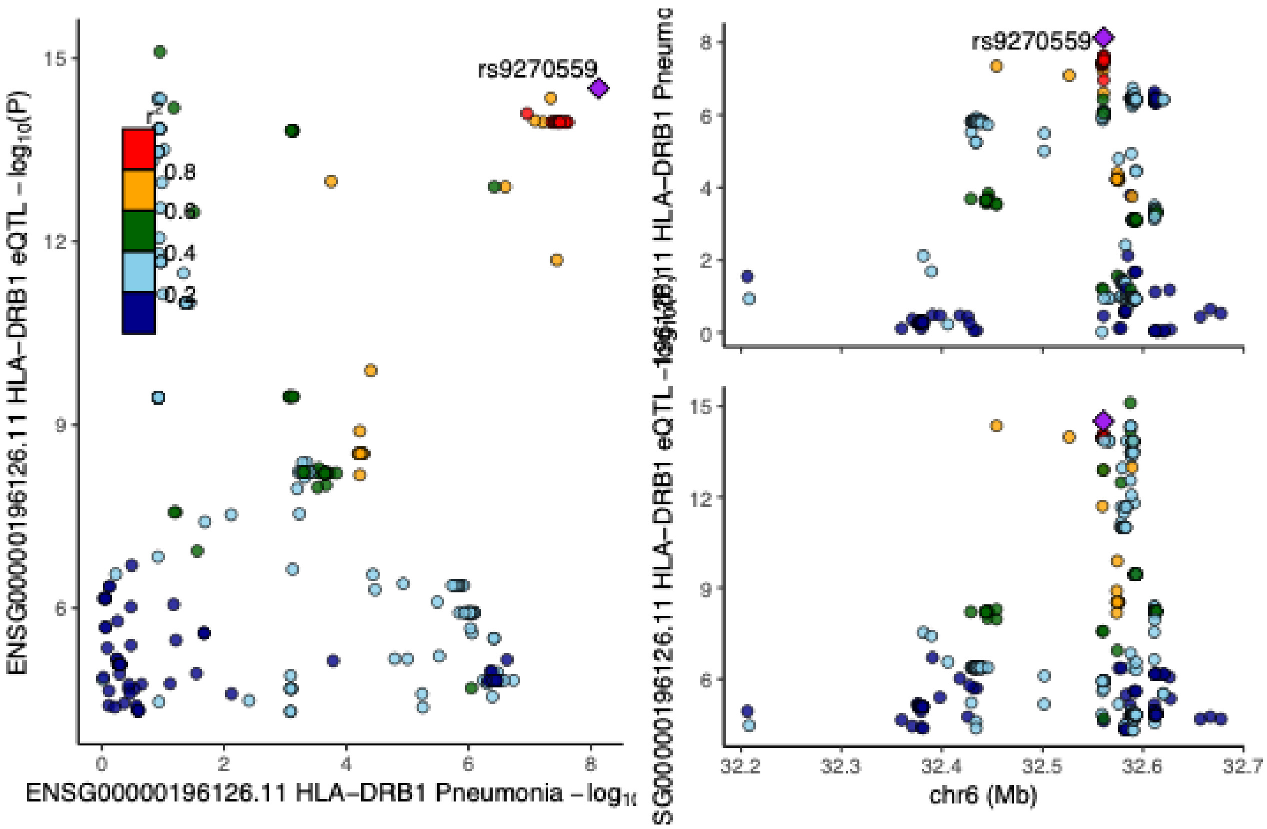

**Figure S31. LocusCompareR[19] plot of HLA-DRB1 gene colocalization between pneumonia and eQTL tissue Cells_EBV-transformed_lymphocytes.** In the left panel, y-axis shows tissue -log10 p-value and x-axis shows pneumonia -log10 p-value. In the right panel, the upper plot shows pneumonia -log10 p-values of variants near colocalized variant and the lower shows tissue -log10 p-values of variants near colocalized variant. Colocalization causal variant is marked with purple diamond and is labeled.

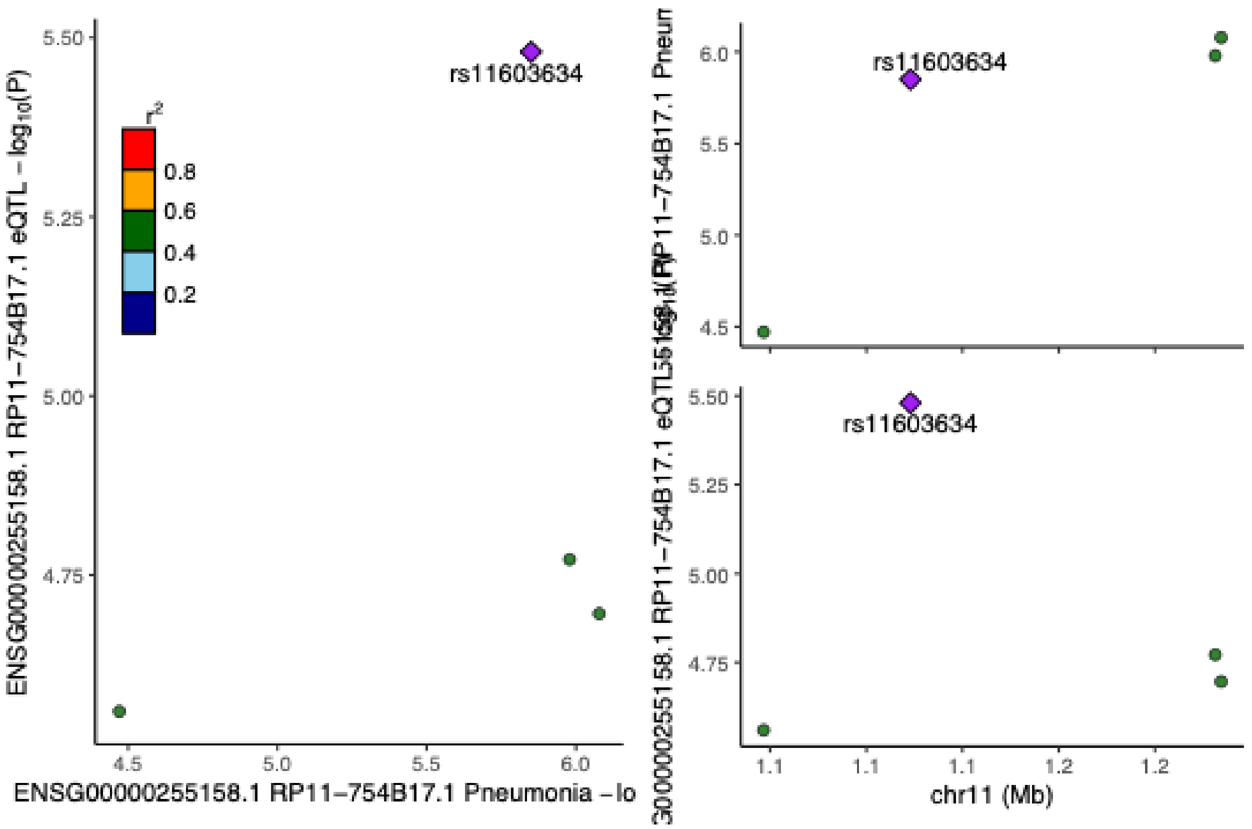

**Figure S32. LocusCompareR[19] plot of RP11-754B17.1 gene colocalization between pneumonia and eQTL tissue Cells_EBV-transformed_lymphocytes.** In the left panel, y-axis shows tissue -log10 p-value and x-axis shows pneumonia -log10 p-value. In the right panel, the upper plot shows pneumonia -log10 p-values of variants near colocalized variant and the lower shows tissue -log10 p-values of variants near colocalized variant. Colocalization causal variant is marked with purple diamond and is labeled.

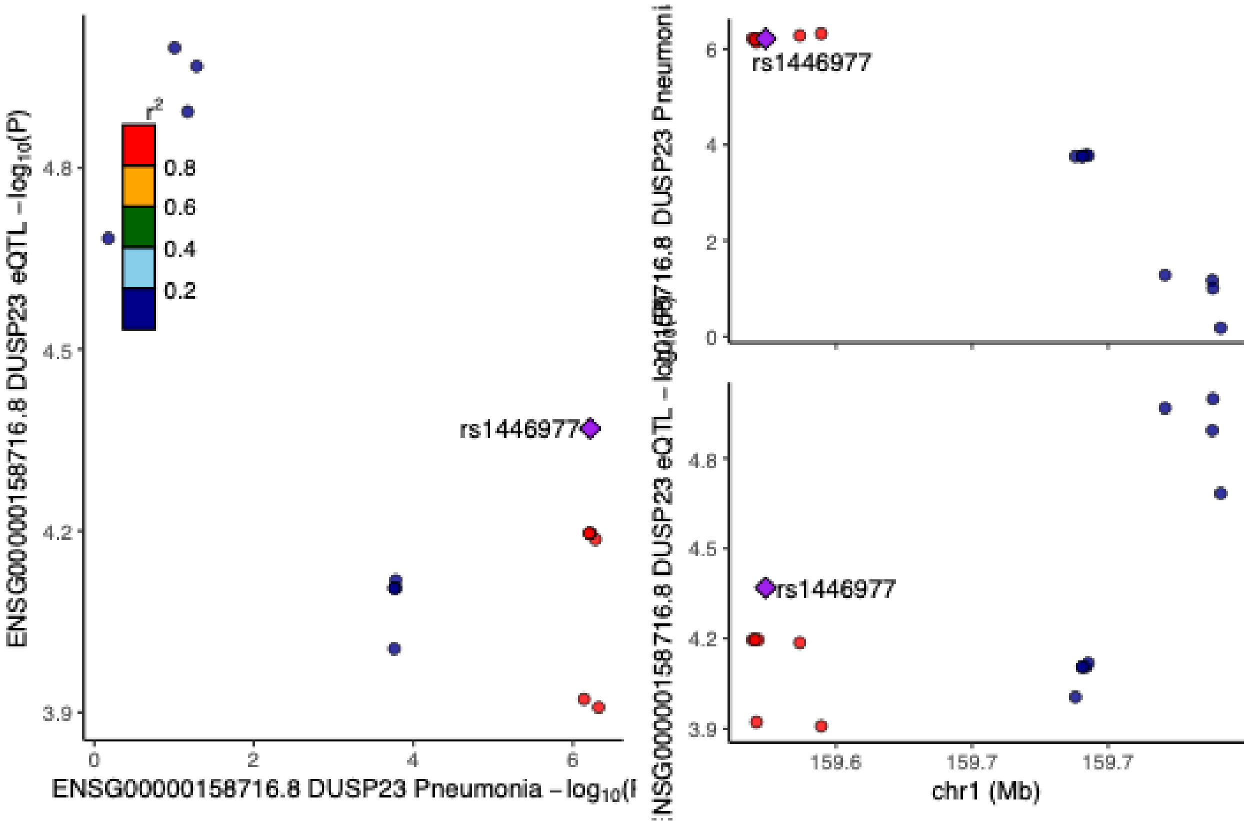

**Figure S33. LocusCompareR[19] plot of DUSP23 gene colocalization between pneumonia and eQTL tissue Whole_blood.** In the left panel, y-axis shows tissue -log10 p-value and x-axis shows pneumonia -log10 p-value. In the right panel, the upper plot shows pneumonia -log10 p-values of variants near colocalized variant and the lower shows tissue -log10 p-values of variants near colocalized variant. Colocalization causal variant is marked with purple diamond and is labeled.

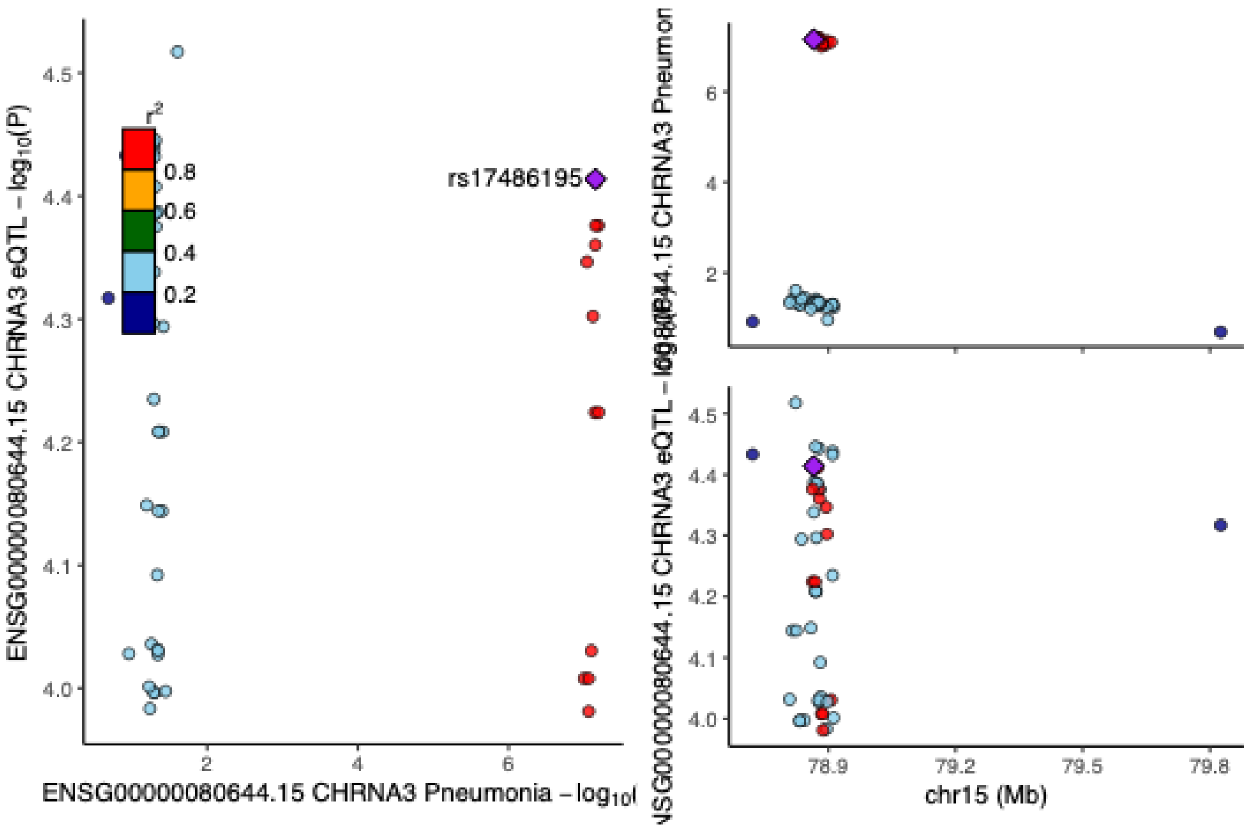

**Figure S34. LocusCompareR[19] plot of CHRNA3 gene colocalization between pneumonia and eQTL tissue Lung.** In the left panel, y-axis shows tissue -log10 p-value and x-axis shows pneumonia -log10 p-value. In the right panel, the upper plot shows pneumonia -log10 p-values of variants near colocalized variant and the lower shows tissue -log10 p-values of variants near colocalized variant. Colocalization causal variant is marked with purple diamond and is labeled.

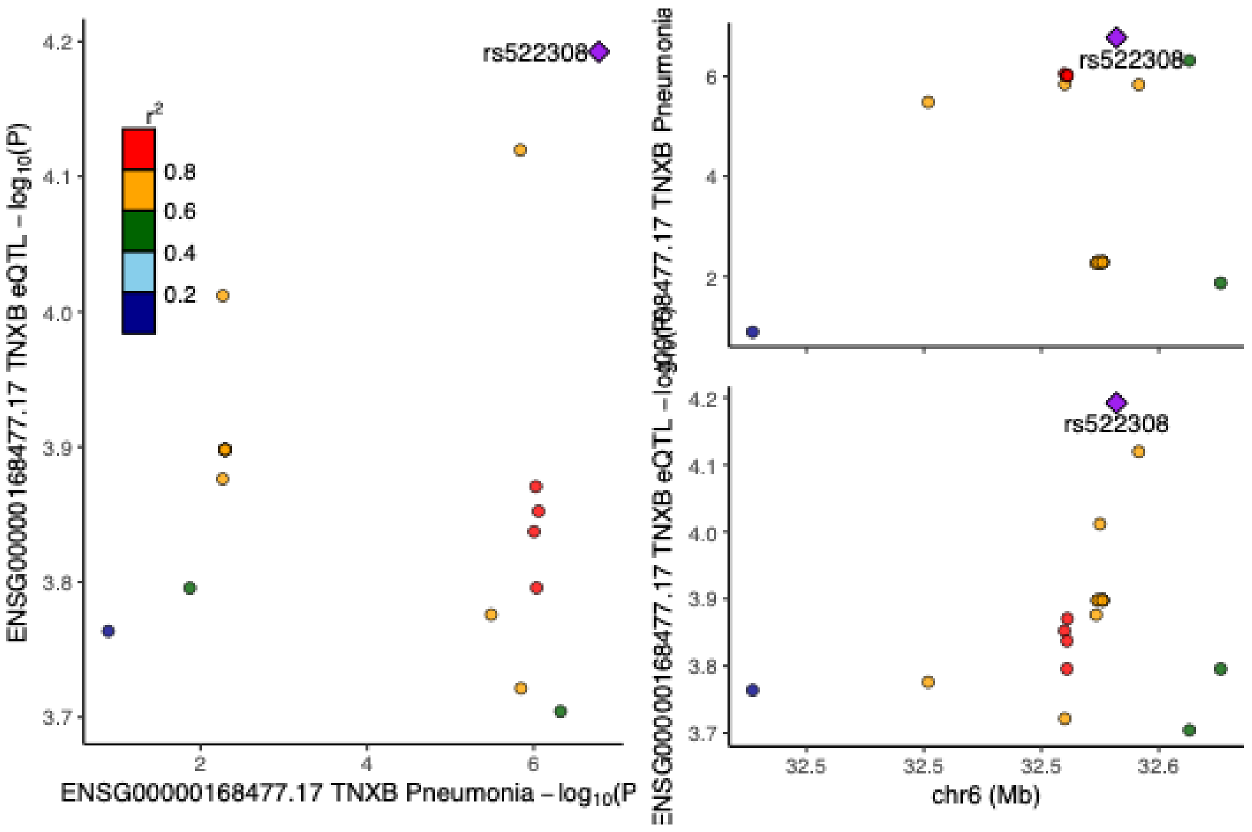

**Figure S35. LocusCompareR[19] plot of TNXB gene colocalization between pneumonia and eQTL tissue Whole_Blood.** In the left panel, y-axis shows tissue -log10 p-value and x-axis shows pneumonia -log10 p-value. In the right panel, the upper plot shows pneumonia -log10 p-values of variants near colocalized variant and the lower shows tissue -log10 p-values of variants near colocalized variant. Colocalization causal variant is marked with purple diamond and is labeled.

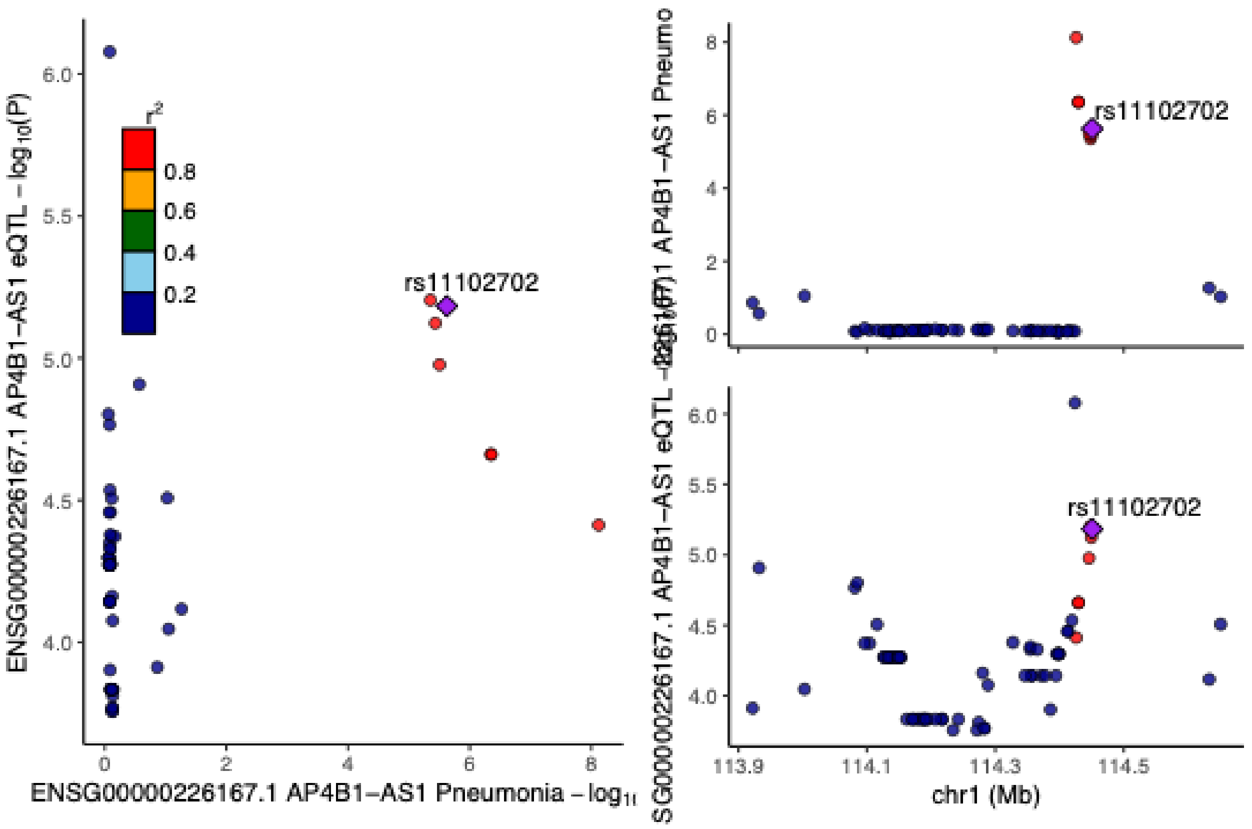

**Figure S36. LocusCompareR[19] plot of AP4B1-AS1 gene colocalization between pneumonia and eQTL tissue Lung.** In the left panel, y-axis shows tissue -log10 p-value and x-axis shows pneumonia -log10 p-value. In the right panel, the upper plot shows pneumonia -log10 p-values of variants near colocalized variant and the lower shows tissue -log10 p-values of variants near colocalized variant. Colocalization causal variant is marked with purple diamond and is labeled.

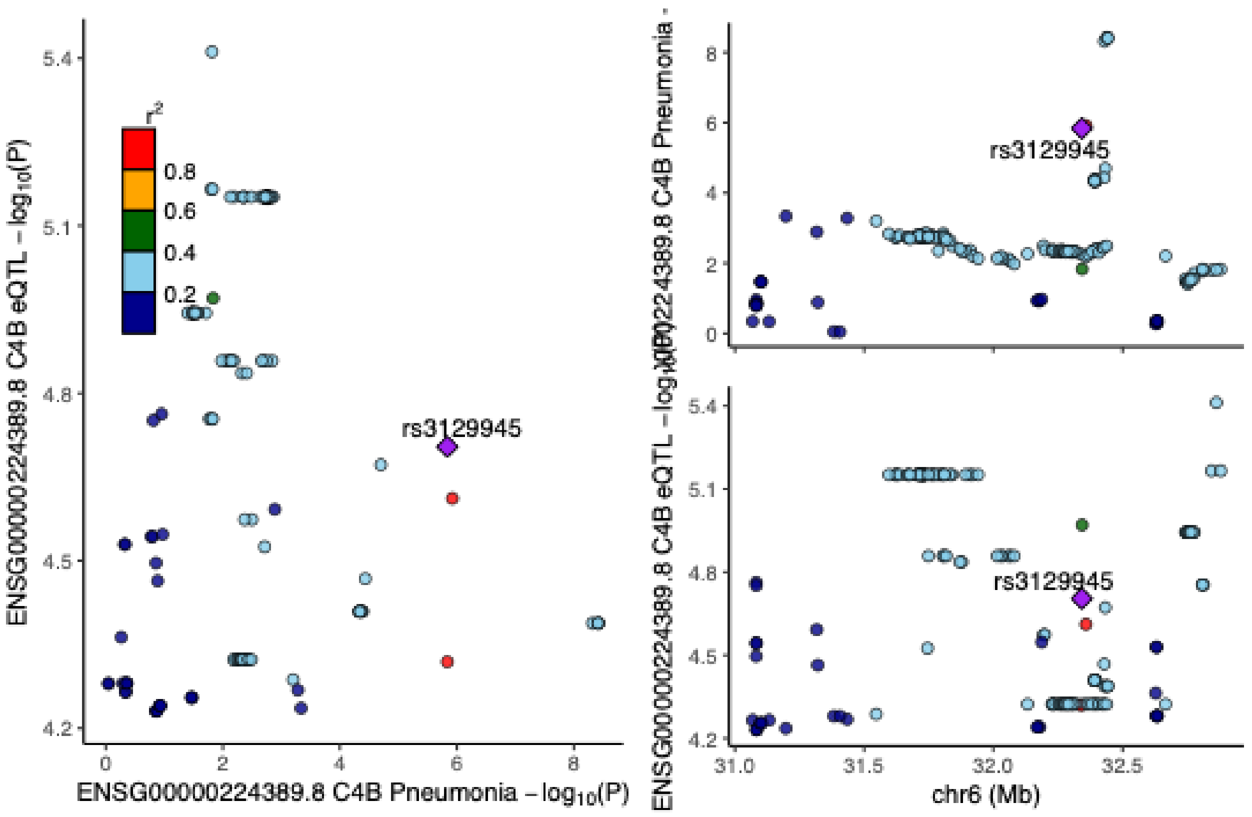

**Figure S37. LocusCompareR[19] plot of C4B gene colocalization between pneumonia and eQTL tissue Cells_EBV-transformed_lymphocytes.** In the left panel, y-axis shows tissue -log10 p-value and x-axis shows pneumonia -log10 p-value. In the right panel, the upper plot shows pneumonia -log10 p-values of variants near colocalized variant and the lower shows tissue -log10 p-values of variants near colocalized variant. Colocalization causal variant is marked with purple diamond and is labeled.

### Table S1. Full list of FinnGen consortium authors.

| Full Name | Affiliation | E-mail | Role 1 | Role 2 |
|---|---|---|---|---|
| Aarno Palotie | Institute for Molecular Medicine Finland (FIMM), HiLIFE, University of Helsinki, Helsinki, Finland; Broad Institute of MIT and Harvard; Massachusetts General Hospital | | Steering Committee | Steering Committee |
| Mark Daly | Institute for Molecular Medicine Finland (FIMM), HiLIFE, University of Helsinki, Helsinki, Finland; Broad Institute of MIT and Harvard; Massachusetts General Hospital | | Steering Committee | Steering Committee |
| Bridget Riley-Gills | Abbvie, Chicago, IL, United States | | Steering Committee | Pharmaceutical companies |
| Howard Jacob | Abbvie, Chicago, IL, United States | | Steering Committee | Pharmaceutical companies |
| Coralie Viollet | Astra Zeneca, Cambridge, United Kingdom | | Steering Committee | Pharmaceutical companies |
| Slavé Petrovski | Astra Zeneca, Cambridge, United Kingdom | | Steering Committee | Pharmaceutical companies |
| Chia-Yen Chen | Biogen, Cambridge, MA, United States | | Steering Committee | Pharmaceutical companies |
| Sally John | Biogen, Cambridge, MA, United States | | Steering Committee | Pharmaceutical companies |
| George Okafo | Boehringer Ingelheim, Ingelheim am Rhein, Germany | | Steering Committee | Pharmaceutical companies |
| Robert Plenge | Bristol Myers Squibb, New York, NY, United States | | Steering Committee | Pharmaceutical companies |
| Joseph Maranville | Bristol Myers Squibb, New York, NY, United States | | Steering Committee | Pharmaceutical companies |
| Mark McCarthy | Genentech, San Francisco, CA, United States | | Steering Committee | Pharmaceutical companies |
| Rion Pendergrass | Genentech, San Francisco, CA, United States | | Steering Committee | Pharmaceutical companies |
| Jonathan Davitte | GlaxoSmithKline, Collegeville, PA, United States | | Steering Committee | Pharmaceutical companies |
| Kirsi Auro | GlaxoSmithKline, Espoo, Finland | | Steering Committee | Pharmaceutical companies |
| Simonne Longerich | Merck, Kenilworth, NJ, United States | | Steering Committee | Pharmaceutical companies |
| Anders Mälarstig | Pfizer, New York, NY, United States | | Steering Committee | Pharmaceutical companies |
| Anna Vlahiotis | Pfizer, New York, NY, United States | | Steering Committee | Pharmaceutical companies |
| Katherine Klinger | Translational Sciences, Sanofi R&D, Framingham, MA, USA | | Steering Committee | Pharmaceutical companies |
| Clement Chatelain | Translational Sciences, Sanofi R&D, Framingham, MA, USA | | Steering Committee | Pharmaceutical companies |
| Matthias Gossel | Translational Sciences, Sanofi R&D, Framingham, MA, USA | | Steering Committee | Pharmaceutical companies |
| Karol Estrada | Maze Therapeutics, San Francisco, CA, United States | | Steering Committee | Pharmaceutical companies |
| Robert Graham | Maze Therapeutics, San Francisco, CA, United States | | Steering Committee | Pharmaceutical companies |
| Dawn Waterworth | Janssen Research & Development, LLC, Spring House, PA, United States | | Steering Committee | Pharmaceutical companies |
| Chris O´Donnell | Novartis Institutes for BioMedical Research, Cambridge, MA, United States | | Steering Committee | Pharmaceutical companies |
| Nicole Renaud | Novartis Institutes for BioMedical Research, Cambridge, MA, United States | | Steering Committee | Pharmaceutical companies |
| Tomi P. Mäkelä | HiLIFE, University of Helsinki, Finland, Finland | | Steering Committee | University of Helsinki & Biobanks |
| Jaakko Kaprio | Institute for Molecular Medicine Finland (FIMM), HiLIFE, University of Helsinki, Helsinki, Finland | | Steering Committee | University of Helsinki & Biobanks |
| Minna Ruddock | Arctic biobank / University of Oulu | | Steering Committee | University of Helsinki & Biobanks |
| Petri Virolainen | Auria Biobank / University of Turku / Hospital District of Southwest Finland, Turku, Finland | | Steering Committee | University of Helsinki & Biobanks |
| Antti Hakanen | Auria Biobank / University of Turku / Hospital District of Southwest Finland, Turku, Finland | | Steering Committee | University of Helsinki & Biobanks |
| Terhi Kilpi | THL Biobank / Finnish Institute for Health and Welfare (THL), Helsinki, Finland | | Steering Committee | University of Helsinki & Biobanks |
| Markus Perola | THL Biobank / Finnish Institute for Health and Welfare (THL), Helsinki, Finland | | Steering Committee | University of Helsinki & Biobanks |
| Jukka Partanen | Finnish Red Cross Blood Service / Finnish Hematology Registry and Clinical Biobank, Helsinki, Finland | | Steering Committee | University of Helsinki & Biobanks |
| Taneli Raivio | Helsinki Biobank / Helsinki University and Hospital District of Helsinki and Uusimaa, Helsinki | | Steering Committee | University of Helsinki & Biobanks |

| | | | | |
|---|---|---|---|---|
| Jani Tikkanen | Northern Finland Biobank Borealis / University of Oulu / Northern Ostrobothnia Hospital District, Oulu, Finland | | Steering Committee | **University of Helsinki & Biobanks** |
| Raisa Serpi | Northern Finland Biobank Borealis / University of Oulu / Northern Ostrobothnia Hospital District, Oulu, Finland | | Steering Committee | **University of Helsinki & Biobanks** |
| Kati Kristiansson | Finnish Clinical Biobank Tampere / University of Tampere / Pirkanmaa Hospital District, Tampere, Finland | | Steering Committee | **University of Helsinki & Biobanks** |
| Veli-Matti Kosma | Biobank of Eastern Finland / University of Eastern Finland / Northern Savo Hospital District, Kuopio, Finland | | Steering Committee | **University of Helsinki & Biobanks** |
| Jari Laukkanen | Central Finland Biobank / University of Jyväskylä / Central Finland Health Care District, Jyväskylä, Finland | | Steering Committee | **University of Helsinki & Biobanks** |
| Marco Hautalahti | FINBB - Finnish biobank cooperative | | Steering Committee | **University of Helsinki & Biobanks** |
| Outi Tuovila | Business Finland, Helsinki, Finland | | Steering Committee | **Other Experts/ Non-Voting Members** |
| Jeffrey Waring | Abbvie, Chicago, IL, United States | | Scientific Committee | **Pharmaceutical companies** |
| Bridget Riley-Gillis | Abbvie, Chicago, IL, United States | | Scientific Committee | **Pharmaceutical companies** |
| Fedik Rahimov | Abbvie, Chicago, IL, United States | | Scientific Committee | **Pharmaceutical companies** |
| Ioanna Tachmazidou | Astra Zeneca, Cambridge, United Kingdom | | Scientific Committee | **Pharmaceutical companies** |
| Chia-Yen Chen | Biogen, Cambridge, MA, United States | | Scientific Committee | **Pharmaceutical companies** |
| Zhihao Ding | Boehringer Ingelheim, Ingelheim am Rhein, Germany | | Scientific Committee | **Pharmaceutical companies** |
| Marc Jung | Boehringer Ingelheim, Ingelheim am Rhein, Germany | | Scientific Committee | **Pharmaceutical companies** |
| Hanati Tuoken | Boehringer Ingelheim, Ingelheim am Rhein, Germany | | Scientific Committee | **Pharmaceutical companies** |
| Shameek Biswas | Bristol Myers Squibb, New York, NY, United States | | Scientific Committee | **Pharmaceutical companies** |
| Rion Pendergrass | Genentech, San Francisco, CA, United States | | Scientific Committee | **Pharmaceutical companies** |
| Jonathan Davitte | GlaxoSmithKline, Collegeville, PA, United States | | Scientific Committee | **Pharmaceutical companies** |
| Neha Raghavan | Merck, Kenilworth, NJ, United States | | Scientific Committee | **Pharmaceutical companies** |
| Adriana Huertas-Vazquez | Merck, Kenilworth, NJ, United States | | Scientific Committee | **Pharmaceutical companies** |
| Jae-Hoon Sul | Merck, Kenilworth, NJ, United States | | Scientific Committee | **Pharmaceutical companies** |
| Anders Mälarstig | Pfizer, New York, NY, United States | | Scientific Committee | **Pharmaceutical companies** |
| Xinli Hu | Pfizer, New York, NY, United States | | Scientific Committee | **Pharmaceutical companies** |
| Åsa Hedman | Pfizer, New York, NY, United States | | Scientific Committee | **Pharmaceutical companies** |
| Katherine Klinger | Translational Sciences, Sanofi R&D, Framingham, MA, USA | | Scientific Committee | **Pharmaceutical companies** |
| Robert Graham | Maze Therapeutics, San Francisco, CA, United States | | Scientific Committee | **Pharmaceutical companies** |
| Dawn Waterworth | Janssen Research & Development, LLC, Spring House, PA, United States | | Scientific Committee | **Pharmaceutical companies** |
| Nicole Renaud | Novartis Institutes for BioMedical Research, Cambridge, MA, United States | | Scientific Committee | **Pharmaceutical companies** |
| Ma´en Obeidat | Novartis Institutes for BioMedical Research, Cambridge, MA, United States | | Scientific Committee | **Pharmaceutical companies** |
| Jonathan Chung | Novartis Institutes for BioMedical Research, Cambridge, MA, United States | | Scientific Committee | **Pharmaceutical companies** |
| Jonas Zierer | Novartis Institutes for BioMedical Research, Cambridge, MA, United States | | Scientific Committee | **Pharmaceutical companies** |
| Mari Niemi | Novartis Institutes for BioMedical Research, Cambridge, MA, United States | | Scientific Committee | **Pharmaceutical companies** |
| Samuli Ripatti | Institute for Molecular Medicine Finland (FIMM), HiLIFE, University of Helsinki, Helsinki, Finland | | Scientific Committee | **University of Helsinki & Biobanks** |
| Johanna Schleutker | Auria Biobank / Univ. of Turku / Hospital District of Southwest Finland, Turku, Finland | | Scientific Committee | **University of Helsinki & Biobanks** |
| Markus Perola | THL Biobank / Finnish Institute for Health and Welfare (THL), Helsinki, Finland | | Scientific Committee | **University of Helsinki & Biobanks** |
| Mikko Arvas | Finnish Red Cross Blood Service / Finnish Hematology Registry and Clinical Biobank, Helsinki, Finland | | Scientific Committee | **University of Helsinki & Biobanks** |
| Olli Carpén | Helsinki Biobank / Helsinki University and Hospital District of Helsinki and Uusimaa, Helsinki | | Scientific Committee | **University of Helsinki & Biobanks** |
| Reetta Hinttala | Northern Finland Biobank Borealis / University of Oulu / Northern Ostrobothnia Hospital District, Oulu, Finland | | Scientific Committee | **University of Helsinki & Biobanks** |

| | | | | |
|---|---|---|---|---|
| Johannes Kettunen | Northern Finland Biobank Borealis / University of Oulu / Northern Ostrobothnia Hospital District, Oulu, Finland | | Scientific Committee | **University of Helsinki & Biobanks** |
| Arto Mannermaa | Biobank of Eastern Finland / University of Eastern Finland / Northern Savo Hospital District, Kuopio, Finland | | Scientific Committee | **University of Helsinki & Biobanks** |
| Katriina Aalto-Setälä | Faculty of Medicine and Health Technology, Tampere University, Tampere, Finland | | Scientific Committee | **University of Helsinki & Biobanks** |
| Mika Kähönen | Finnish Clinical Biobank Tampere / University of Tampere / Pirkanmaa Hospital District, Tampere, Finland | | Scientific Committee | **University of Helsinki & Biobanks** |
| Jari Laukkanen | Central Finland Biobank / University of Jyväskylä / Central Finland Health Care District, Jyväskylä, Finland | | Scientific Committee | **University of Helsinki & Biobanks** |
| Johanna Mäkelä | FINBB - Finnish biobank cooperative | | Scientific Committee | **University of Helsinki & Biobanks** |
| Reetta Kälviäinen | Northern Savo Hospital District, Kuopio, Finland | | Clinical Groups | **Neurology Group** |
| Valtteri Julkunen | Northern Savo Hospital District, Kuopio, Finland | | Clinical Groups | **Neurology Group** |
| Hilkka Soininen | Northern Savo Hospital District, Kuopio, Finland | | Clinical Groups | **Neurology Group** |
| Anne Remes | Northern Ostrobothnia Hospital District, Oulu, Finland | | Clinical Groups | **Neurology Group** |
| Mikko Hiltunen | University of Eastern Finland, Kuopio, Finland | | Clinical Groups | **Neurology Group** |
| Jukka Peltola | Pirkanmaa Hospital District, Tampere, Finland | | Clinical Groups | **Neurology Group** |
| Minna Raivio | Hospital District of Helsinki and Uusimaa, Helsinki, Finland | | Clinical Groups | **Neurology Group** |
| Pentti Tienari | Hospital District of Helsinki and Uusimaa, Helsinki, Finland | | Clinical Groups | **Neurology Group** |
| Juha Rinne | Hospital District of Southwest Finland, Turku, Finland | | Clinical Groups | **Neurology Group** |
| Roosa Kallionpää | Hospital District of Southwest Finland, Turku, Finland | | Clinical Groups | **Neurology Group** |
| Juulia Partanen | Institute for Molecular Medicine Finland, HiLIFE, University of Helsinki, Finland | | Clinical Groups | **Neurology Group** |
| Adam Ziemann | Abbvie, Chicago, IL, United States | | Clinical Groups | **Neurology Group** |
| Nizar Smaoui | Abbvie, Chicago, IL, United States | | Clinical Groups | **Neurology Group** |
| Anne Lehtonen | Abbvie, Chicago, IL, United States | | Clinical Groups | **Neurology Group** |
| Susan Eaton | Biogen, Cambridge, MA, United States | | Clinical Groups | **Neurology Group** |
| Heiko Runz | Biogen, Cambridge, MA, United States | | Clinical Groups | **Neurology Group** |
| Sanni Lahdenperä | Biogen, Cambridge, MA, United States | | Clinical Groups | **Neurology Group** |
| Shameek Biswas | Bristol Myers Squibb, New York, NY, United States | | Clinical Groups | **Neurology Group** |
| Natalie Bowers | Genentech, San Francisco, CA, United States | | Clinical Groups | **Neurology Group** |
| Edmond Teng | Genentech, San Francisco, CA, United States | | Clinical Groups | **Neurology Group** |
| Rion Pendergrass | Genentech, San Francisco, CA, United States | | Clinical Groups | **Neurology Group** |
| Fanli Xu | GlaxoSmithKline, Brentford, United Kingdom | | Clinical Groups | **Neurology Group** |
| Kirsi Auro | GlaxoSmithKline, Espoo, Finland | | Clinical Groups | **Neurology Group** |
| Laura Addis | GlaxoSmithKline, Brentford, United Kingdom | | Clinical Groups | **Neurology Group** |
| John Eicher | GlaxoSmithKline, Brentford, United Kingdom | | Clinical Groups | **Neurology Group** |
| Qingqin S Li | Janssen Research & Development, LLC, Titusville, NJ 08560, United States | | Clinical Groups | **Neurology Group** |
| Karen He | Janssen Research & Development, LLC, Spring House, PA, United States | | Clinical Groups | **Neurology Group** |
| Ekaterina Khramtsova | Janssen Research & Development, LLC, Spring House, PA, United States | | Clinical Groups | **Neurology Group** |
| Neha Raghavan | Merck, Kenilworth, NJ, United States | | Clinical Groups | **Neurology Group** |
| Martti Färkkilä | Hospital District of Helsinki and Uusimaa, Helsinki, Finland | | Clinical Groups | **Gastroenterology Group** |
| Jukka Koskela | Hospital District of Helsinki and Uusimaa, Helsinki, Finland | | Clinical Groups | **Gastroenterology Group** |
| Sampsa Pikkarainen | Hospital District of Helsinki and Uusimaa, Helsinki, Finland | | Clinical Groups | **Gastroenterology Group** |
| Airi Jussila | Pirkanmaa Hospital District, Tampere, Finland | | Clinical Groups | **Gastroenterology Group** |
| Katri Kaukinen | Pirkanmaa Hospital District, Tampere, Finland | | Clinical Groups | **Gastroenterology Group** |
| Timo Blomster | Northern Ostrobothnia Hospital District, Oulu, Finland | | Clinical Groups | **Gastroenterology Group** |
| Mikko Kiviniemi | Northern Savo Hospital District, Kuopio, Finland | | Clinical Groups | **Gastroenterology Group** |
| Markku Voutilainen | Hospital District of Southwest Finland, Turku, Finland | | Clinical Groups | **Gastroenterology Group** |

| | | | | |
|---|---|---|---|---|
| Mark Daly | Institute for Molecular Medicine, Finland (FIMM), HiLIFE, University of Helsinki, Helsinki, Finland; Broad Institute of MIT and Harvard; Massachusetts General Hospital | | Clinical Groups | **Gastroenterology Group** |
| Jeffrey Waring | Abbvie, Chicago, IL, United States | | Clinical Groups | **Gastroenterology Group** |
| Nizar Smaoui | Abbvie, Chicago, IL, United States | | Clinical Groups | **Gastroenterology Group** |
| Fedik Rahimov | Abbvie, Chicago, IL, United States | | Clinical Groups | **Gastroenterology Group** |
| Anne Lehtonen | Abbvie, Chicago, IL, United States | | Clinical Groups | **Gastroenterology Group** |
| Tim Lu | Genentech, San Francisco, CA, United States | | Clinical Groups | **Gastroenterology Group** |
| Natalie Bowers | Genentech, San Francisco, CA, United States | | Clinical Groups | **Gastroenterology Group** |
| Rion Pendergrass | Genentech, San Francisco, CA, United States | | Clinical Groups | **Gastroenterology Group** |
| Linda McCarthy | GlaxoSmithKline, Brentford, United Kingdom | | Clinical Groups | **Gastroenterology Group** |
| Amy Hart | Janssen Research & Development, LLC, Spring House, PA, United States | | Clinical Groups | **Gastroenterology Group** |
| Meijian Guan | Janssen Research & Development, LLC, Spring House, PA, United States | | Clinical Groups | **Gastroenterology Group** |
| Jason Miller | Merck, Kenilworth, NJ, United States | | Clinical Groups | **Gastroenterology Group** |
| Kirsi Kalpala | Pfizer, New York, NY, United States | | Clinical Groups | **Gastroenterology Group** |
| Melissa Miller | Pfizer, New York, NY, United States | | Clinical Groups | **Gastroenterology Group** |
| Xinli Hu | Pfizer, New York, NY, United States | | Clinical Groups | **Gastroenterology Group** |
| Kari Eklund | Hospital District of Helsinki and Uusimaa, Helsinki, Finland | | Clinical Groups | **Rheumatology Group** |
| Antti Palomäki | Hospital District of Southwest Finland, Turku, Finland | | Clinical Groups | **Rheumatology Group** |
| Pia Isomäki | Pirkanmaa Hospital District, Tampere, Finland | | Clinical Groups | **Rheumatology Group** |
| Laura Pirilä | Hospital District of Southwest Finland, Turku, Finland | | Clinical Groups | **Rheumatology Group** |
| Oili Kaipiainen-Seppänen | Northern Savo Hospital District, Kuopio, Finland | | Clinical Groups | **Rheumatology Group** |
| Johanna Huhtakangas | Northern Ostrobothnia Hospital District, Oulu, Finland | | Clinical Groups | **Rheumatology Group** |
| Nina Mars | Institute for Molecular Medicine Finland (FIMM), HiLIFE, University of Helsinki, Helsinki, Finland | | Clinical Groups | **Rheumatology Group** |
| Jeffrey Waring | Abbvie, Chicago, IL, United States | | Clinical Groups | **Rheumatology Group** |
| Fedik Rahimov | Abbvie, Chicago, IL, United States | | Clinical Groups | **Rheumatology Group** |
| Apinya Lertratanakul | Abbvie, Chicago, IL, United States | | Clinical Groups | **Rheumatology Group** |
| Nizar Smaoui | Abbvie, Chicago, IL, United States | | Clinical Groups | **Rheumatology Group** |
| Anne Lehtonen | Abbvie, Chicago, IL, United States | | Clinical Groups | **Rheumatology Group** |
| Coralie Viollet | AstraZeneca, Cambridge, United Kingdom | | Clinical Groups | **Rheumatology Group** |
| Marla Hochfeld | Bristol Myers Squibb, New York, NY, United States | | Clinical Groups | **Rheumatology Group** |
| Natalie Bowers | Genentech, San Francisco, CA, United States | | Clinical Groups | **Rheumatology Group** |
| Rion Pendergrass | Genentech, San Francisco, CA, United States | | Clinical Groups | **Rheumatology Group** |
| Jorge Esparza Gordillo | GlaxoSmithKline, Brentford, United Kingdom | | Clinical Groups | **Rheumatology Group** |
| Kirsi Auro | GlaxoSmithKline, Espoo, Finland | | Clinical Groups | **Rheumatology Group** |
| Dawn Waterworth | Janssen Research & Development, LLC, Spring House, PA, United States | | Clinical Groups | **Rheumatology Group** |
| Fabiana Farias | Merck, Kenilworth, NJ, United States | | Clinical Groups | **Rheumatology Group** |
| Kirsi Kalpala | Pfizer, New York, NY, United States | | Clinical Groups | **Rheumatology Group** |
| Nan Bing | Pfizer, New York, NY, United States | | Clinical Groups | **Rheumatology Group** |
| Xinli Hu | Pfizer, New York, NY, United States | | Clinical Groups | **Rheumatology Group** |
| Tarja Laitinen | Pirkanmaa Hospital District, Tampere, Finland | | Clinical Groups | **Pulmonology Group** |
| Margit Pelkonen | Northern Savo Hospital District, Kuopio, Finland | | Clinical Groups | **Pulmonology Group** |
| Paula Kauppi | Hospital District of Helsinki and Uusimaa, Helsinki, Finland | | Clinical Groups | **Pulmonology Group** |
| Hannu Kankaanranta | University of Gothenburg, Gothenburg, Sweden/ Seinäjoki Central Hospital, Seinäjoki, Finland/ Tampere University, Tampere, Finland | | Clinical Groups | **Pulmonology Group** |
| Terttu Harju | Northern Ostrobothnia Hospital District, Oulu, Finland | | Clinical Groups | **Pulmonology Group** |
| Riitta Lahesmaa | Hospital District of Southwest Finland, Turku, Finland | | Clinical Groups | **Pulmonology Group** |
| Nizar Smaoui | Abbvie, Chicago, IL, United States | | Clinical Groups | **Pulmonology Group** |
| Coralie Viollet | AstraZeneca, Cambridge, United Kingdom | | Clinical Groups | **Pulmonology Group** |

| | | | | |
|---|---|---|---|---|
| Susan Eaton | Biogen, Cambridge, MA, United States | | Clinical Groups | **Pulmonology Group** |
| Hubert Chen | Genentech, San Francisco, CA, United States | | Clinical Groups | **Pulmonology Group** |
| Rion Pendergrass | Genentech, San Francisco, CA, United States | | Clinical Groups | **Pulmonology Group** |
| Natalie Bowers | Genentech, San Francisco, CA, United States | | Clinical Groups | **Pulmonology Group** |
| Joanna Betts | GlaxoSmithKline, Brentford, United Kingdom | | Clinical Groups | **Pulmonology Group** |
| Kirsi Auro | GlaxoSmithKline, Espoo, Finland | | Clinical Groups | **Pulmonology Group** |
| Rajashree Mishra | GlaxoSmithKline, Brentford, United Kingdom | | Clinical Groups | **Pulmonology Group** |
| Majd Mouded | Novartis, Basel, Switzerland | | Clinical Groups | **Pulmonology Group** |
| Debby Ngo | Novartis, Basel, Switzerland | | Clinical Groups | **Pulmonology Group** |
| Teemu Niiranen | Finnish Institute for Health and Welfare (THL), Helsinki, Finland | | Clinical Groups | **Cardiometabolic Diseases Group** |
| Felix Vaura | Finnish Institute for Health and Welfare (THL), Helsinki, Finland | | Clinical Groups | **Cardiometabolic Diseases Group** |
| Veikko Salomaa | Finnish Institute for Health and Welfare (THL), Helsinki, Finland | | Clinical Groups | **Cardiometabolic Diseases Group** |
| Kaj Metsärinne | Hospital District of Southwest Finland, Turku, Finland | | Clinical Groups | **Cardiometabolic Diseases Group** |
| Jenni Aittokallio | Hospital District of Southwest Finland, Turku, Finland | | Clinical Groups | **Cardiometabolic Diseases Group** |
| Mika Kähönen | Pirkanmaa Hospital District, Tampere, Finland | | Clinical Groups | **Cardiometabolic Diseases Group** |
| Jussi Hernesniemi | Pirkanmaa Hospital District, Tampere, Finland | | Clinical Groups | **Cardiometabolic Diseases Group** |
| Daniel Gordin | Hospital District of Helsinki and Uusimaa, Helsinki, Finland | | Clinical Groups | **Cardiometabolic Diseases Group** |
| Juha Sinisalo | Hospital District of Helsinki and Uusimaa, Helsinki, Finland | | Clinical Groups | **Cardiometabolic Diseases Group** |
| Marja-Riitta Taskinen | Hospital District of Helsinki and Uusimaa, Helsinki, Finland | | Clinical Groups | **Cardiometabolic Diseases Group** |
| Tiinamaija Tuomi | Hospital District of Helsinki and Uusimaa, Helsinki, Finland | | Clinical Groups | **Cardiometabolic Diseases Group** |
| Timo Hiltunen | Hospital District of Helsinki and Uusimaa, Helsinki, Finland | | Clinical Groups | **Cardiometabolic Diseases Group** |
| Jari Laukkanen | Central Finland Health Care District, Jyväskylä, Finland | | Clinical Groups | **Cardiometabolic Diseases Group** |
| Amanda Elliott | Institute for Molecular Medicine Finland (FIMM), HiLIFE, University of Helsinki, Helsinki, Finland; Broad Institute, Cambridge, MA, USA and Massachusetts General Hospital, Boston, MA, USA | | Clinical Groups | **Cardiometabolic Diseases Group** |
| Mary Pat Reeve | Institute for Molecular Medicine Finland (FIMM), HiLIFE, University of Helsinki, Helsinki, Finland | | Clinical Groups | **Cardiometabolic Diseases Group** |
| Sanni Ruotsalainen | Institute for Molecular Medicine Finland (FIMM), HiLIFE, University of Helsinki, Helsinki, Finland | | Clinical Groups | **Cardiometabolic Diseases Group** |
| Dirk Paul | Astra Zeneca, Cambridge, United Kingdom | | Clinical Groups | **Cardiometabolic Diseases Group** |
| Natalie Bowers | Genentech, San Francisco, CA, United States | | Clinical Groups | **Cardiometabolic Diseases Group** |
| Rion Pendergrass | Genentech, San Francisco, CA, United States | | Clinical Groups | **Cardiometabolic Diseases Group** |
| Audrey Chu | GlaxoSmithKline, Brentford, United Kingdom | | Clinical Groups | **Cardiometabolic Diseases Group** |
| Kirsi Auro | GlaxoSmithKline, Espoo, Finland | | Clinical Groups | **Cardiometabolic Diseases Group** |
| Dermot Reilly | Janssen Research & Development, LLC, Boston, MA, United States | | Clinical Groups | **Cardiometabolic Diseases Group** |
| Mike Mendelson | Novartis, Boston, MA, United States | | Clinical Groups | **Cardiometabolic Diseases Group** |
| Jaakko Parkkinen | Pfizer, New York, NY, United States | | Clinical Groups | **Cardiometabolic Diseases Group** |
| Melissa Miller | Pfizer, New York, NY, United States | | Clinical Groups | **Cardiometabolic Diseases Group** |
| Tuomo Meretoja | Department of Breast Surgery, Helsinki University Hospital Comprehensive Cancer Center and University of Helsinki, Helsinki, Finland | | Clinical Groups | **Oncology Group** |
| Heikki Joensuu | Department of Oncology, Helsinki University Hospital Comprehensive Cancer Center and University of Helsinki, Helsinki, Finland | | Clinical Groups | **Oncology Group** |
| Olli Carpén | Hospital District of Helsinki and Uusimaa, Helsinki, Finland | | Clinical Groups | **Oncology Group** |
| Johanna Mattson | Hospital District of Helsinki and Uusimaa, Helsinki, Finland | | Clinical Groups | **Oncology Group** |
| Eveliina Salminen | Hospital District of Helsinki and Uusimaa, Helsinki, Finland | | Clinical Groups | **Oncology Group** |
| Annika Auranen | Pirkanmaa Hospital District , Tampere, Finland | | Clinical Groups | **Oncology Group** |
| Peeter Karihtala | Department of Oncology, Helsinki University Hospital Comprehensive Cancer Center and University of Helsinki, Helsinki, Finland | | Clinical Groups | **Oncology Group** |

| | | | | |
|---|---|---|---|---|
| Päivi Auvinen | Northern Savo Hospital District, Kuopio, Finland | | Clinical Groups | **Oncology Group** |
| Klaus Elenius | Hospital District of Southwest Finland, Turku, Finland | | Clinical Groups | **Oncology Group** |
| Johanna Schleutker | Hospital District of Southwest Finland, Turku, Finland | | Clinical Groups | **Oncology Group** |
| Esa Pitkänen | Institute for Molecular Medicine Finland (FIMM), HiLIFE, University of Helsinki, Helsinki, Finland | | Clinical Groups | **Oncology Group** |
| Nina Mars | Institute for Molecular Medicine Finland (FIMM), HiLIFE, University of Helsinki, Helsinki, Finland | | Clinical Groups | **Oncology Group** |
| Mark Daly | Institute for Molecular Medicine Finland (FIMM), HiLIFE, University of Helsinki, Helsinki, Finland; Broad Institute of MIT and Harvard; Massachusetts General Hospital | | Clinical Groups | **Oncology Group** |
| Relja Popovic | Abbvie, Chicago, IL, United States | | Clinical Groups | **Oncology Group** |
| Jeffrey Waring | Abbvie, Chicago, IL, United States | | Clinical Groups | **Oncology Group** |
| Bridget Riley-Gillis | Abbvie, Chicago, IL, United States | | Clinical Groups | **Oncology Group** |
| Anne Lehtonen | Abbvie, Chicago, IL, United States | | Clinical Groups | **Oncology Group** |
| Margarete Fabre | AstraZeneca, Cambridge, United Kingdom | | Clinical Groups | **Oncology Group** |
| Jennifer Schutzman | Genentech, San Francisco, CA, United States | | Clinical Groups | **Oncology Group** |
| Natalie Bowers | Genentech, San Francisco, CA, United States | | Clinical Groups | **Oncology Group** |
| Rion Pendergrass | Genentech, San Francisco, CA, United States | | Clinical Groups | **Oncology Group** |
| Diptee Kulkarni | GlaxoSmithKline, Brentford, United Kingdom | | Clinical Groups | **Oncology Group** |
| Kirsi Auro | GlaxoSmithKline, Espoo, Finland | | Clinical Groups | **Oncology Group** |
| Alessandro Porello | Janssen Research & Development, LLC, Spring House, PA, United States | | Clinical Groups | **Oncology Group** |
| Andrey Loboda | Merck, Kenilworth, NJ, United States | | Clinical Groups | **Oncology Group** |
| Heli Lehtonen | Pfizer, New York, NY, United States | | Clinical Groups | **Oncology Group** |
| Stefan McDonough | Pfizer, New York, NY, United States | | Clinical Groups | **Oncology Group** |
| Sauli Vuoti | Janssen-Cilag Oy, Espoo, Finland | | Clinical Groups | **Oncology Group** |
| Kai Kaarniranta | Northern Savo Hospital District, Kuopio, Finland; Department of Molecular Genetics, University of Lodz, Lodz, Poland | | Clinical Groups | **Opthalmology Group** |
| Joni A Turunen | Helsinki University Hospital and University of Helsinki, Helsinki, Finland; Eye Genetics Group, Folkhälsan Research Center, Helsinki, Finland | | Clinical Groups | **Opthalmology Group** |
| Terhi Ollila | Hospital District of Helsinki and Uusimaa, Helsinki, Finland | | Clinical Groups | **Opthalmology Group** |
| Hannu Uusitalo | Pirkanmaa Hospital District, Tampere, Finland | | Clinical Groups | **Opthalmology Group** |
| Juha Karjalainen | Institute for Molecular Medicine Finland (FIMM), HiLIFE, University of Helsinki, Helsinki, Finland | | Clinical Groups | **Opthalmology Group** |
| Esa Pitkänen | Institute for Molecular Medicine Finland (FIMM), HiLIFE, University of Helsinki, Helsinki, Finland | | Clinical Groups | **Opthalmology Group** |
| Mengzhen Liu | Abbvie, Chicago, IL, United States | | Clinical Groups | **Opthalmology Group** |
| Heiko Runz | Biogen, Cambridge, MA, United States | | Clinical Groups | **Opthalmology Group** |
| Stephanie Loomis | Biogen, Cambridge, MA, United States | | Clinical Groups | **Opthalmology Group** |
| Erich Strauss | Genentech, San Francisco, CA, United States | | Clinical Groups | **Opthalmology Group** |
| Natalie Bowers | Genentech, San Francisco, CA, United States | | Clinical Groups | **Opthalmology Group** |
| Hao Chen | Genentech, San Francisco, CA, United States | | Clinical Groups | **Opthalmology Group** |
| Rion Pendergrass | Genentech, San Francisco, CA, United States | | Clinical Groups | **Opthalmology Group** |
| Kaisa Tasanen | Northern Ostrobothnia Hospital District, Oulu, Finland | | Clinical Groups | **Dermatology Group** |
| Laura Huilaja | Northern Ostrobothnia Hospital District, Oulu, Finland | | Clinical Groups | **Dermatology Group** |
| Katariina Hannula-Jouppi | Hospital District of Helsinki and Uusimaa, Helsinki, Finland | | Clinical Groups | **Dermatology Group** |
| Teea Salmi | Pirkanmaa Hospital District, Tampere, Finland | | Clinical Groups | **Dermatology Group** |
| Sirkku Peltonen | Hospital District of Southwest Finland, Turku, Finland | | Clinical Groups | **Dermatology Group** |
| Leena Koulu | Hospital District of Southwest Finland, Turku, Finland | | Clinical Groups | **Dermatology Group** |
| Nizar Smaoui | Abbvie, Chicago, IL, United States | | Clinical Groups | **Dermatology Group** |
| Fedik Rahimov | Abbvie, Chicago, IL, United States | | Clinical Groups | **Dermatology Group** |
| Anne Lehtonen | Abbvie, Chicago, IL, United States | | Clinical Groups | **Dermatology Group** |
| David Choy | Genentech, San Francisco, CA, United States | | Clinical Groups | **Dermatology Group** |
| Rion Pendergrass | Genentech, San Francisco, CA, United States | | Clinical Groups | **Dermatology Group** |

| | | | | |
|---|---|---|---|---|
| Dawn Waterworth | Janssen Research & Development, LLC, Spring House, PA, United States | | Clinical Groups | **Dermatology Group** |
| Kirsi Kalpala | Pfizer, New York, NY, United States | | Clinical Groups | **Dermatology Group** |
| Ying Wu | Pfizer, New York, NY, United States | | Clinical Groups | **Dermatology Group** |
| Pirkko Pussinen | Hospital District of Helsinki and Uusimaa, Helsinki, Finland | | Clinical Groups | **Odontology Group** |
| Aino Salminen | Hospital District of Helsinki and Uusimaa, Helsinki, Finland | | Clinical Groups | **Odontology Group** |
| Tuula Salo | Hospital District of Helsinki and Uusimaa, Helsinki, Finland | | Clinical Groups | **Odontology Group** |
| David Rice | Hospital District of Helsinki and Uusimaa, Helsinki, Finland | | Clinical Groups | **Odontology Group** |
| Pekka Nieminen | Hospital District of Helsinki and Uusimaa, Helsinki, Finland | | Clinical Groups | **Odontology Group** |
| Ulla Palotie | Hospital District of Helsinki and Uusimaa, Helsinki, Finland | | Clinical Groups | **Odontology Group** |
| Maria Siponen | Northern Savo Hospital District, Kuopio, Finland | | Clinical Groups | **Odontology Group** |
| Liisa Suominen | Northern Savo Hospital District, Kuopio, Finland | | Clinical Groups | **Odontology Group** |
| Päivi Mäntylä | Northern Savo Hospital District, Kuopio, Finland | | Clinical Groups | **Odontology Group** |
| Ulvi Gursoy | Hospital District of Southwest Finland, Turku, Finland | | Clinical Groups | **Odontology Group** |
| Vuokko Anttonen | Northern Ostrobothnia Hospital District, Oulu, Finland | | Clinical Groups | **Odontology Group** |
| Kirsi Sipilä | Research Unit of Oral Health Sciences Faculty of Medicine, University of Oulu, Oulu, Finland; Medical Research Center, Oulu, Oulu University Hospital and University of Oulu, Oulu, Finland | | Clinical Groups | **Odontology Group** |
| Rion Pendergrass | Genentech, San Francisco, CA, United States | | Clinical Groups | **Odontology Group** |
| Hannele Laivuori | Institute for Molecular Medicine Finland (FIMM), HiLIFE, University of Helsinki, Helsinki, Finland | | Clinical Groups | **Women's Health and Reproduction Group** |
| Venla Kurra | Pirkanmaa Hospital District, Tampere, Finland | | Clinical Groups | **Women's Health and Reproduction Group** |
| Laura Kotaniemi-Talonen | Pirkanmaa Hospital District, Tampere, Finland | | Clinical Groups | **Women's Health and Reproduction Group** |
| Oskari Heikinheimo | Hospital District of Helsinki and Uusimaa, Helsinki, Finland | | Clinical Groups | **Women's Health and Reproduction Group** |
| Ilkka Kalliala | Hospital District of Helsinki and Uusimaa, Helsinki, Finland | | Clinical Groups | **Women's Health and Reproduction Group** |
| Lauri Aaltonen | Hospital District of Helsinki and Uusimaa, Helsinki, Finland | | Clinical Groups | **Women's Health and Reproduction Group** |
| Varpu Jokimaa | Hospital District of Southwest Finland, Turku, Finland | | Clinical Groups | **Women's Health and Reproduction Group** |
| Johannes Kettunen | Northern Ostrobothnia Hospital District, Oulu, Finland | | Clinical Groups | **Women's Health and Reproduction Group** |
| Marja Vääräsmäki | Northern Ostrobothnia Hospital District, Oulu, Finland | | Clinical Groups | **Women's Health and Reproduction Group** |
| Outi Uimari | Northern Ostrobothnia Hospital District, Oulu, Finland | | Clinical Groups | **Women's Health and Reproduction Group** |
| Laure Morin-Papunen | Northern Ostrobothnia Hospital District, Oulu, Finland | | Clinical Groups | **Women's Health and Reproduction Group** |
| Maarit Niinimäki | Northern Ostrobothnia Hospital District, Oulu, Finland | | Clinical Groups | **Women's Health and Reproduction Group** |
| Terhi Piltonen | Northern Ostrobothnia Hospital District, Oulu, Finland | | Clinical Groups | **Women's Health and Reproduction Group** |
| Katja Kivinen | Institute for Molecular Medicine Finland (FIMM), HiLIFE, University of Helsinki, Helsinki, Finland | | Clinical Groups | **Women's Health and Reproduction Group** |
| Elisabeth Widen | Institute for Molecular Medicine Finland (FIMM), HiLIFE, University of Helsinki, Helsinki, Finland | | Clinical Groups | **Women's Health and Reproduction Group** |
| Taru Tukiainen | Institute for Molecular Medicine Finland (FIMM), HiLIFE, University of Helsinki, Helsinki, Finland | | Clinical Groups | **Women's Health and Reproduction Group** |
| Mary Pat Reeve | Institute for Molecular Medicine Finland (FIMM), HiLIFE, University of Helsinki, Helsinki, Finland | | Clinical Groups | **Women's Health and Reproduction Group** |
| Mark Daly | Institute for Molecular Medicine Finland (FIMM), HiLIFE, University of Helsinki, Helsinki, Finland; Broad Institute of MIT and Harvard; Massachusetts General Hospital | | Clinical Groups | **Women's Health and Reproduction Group** |
| Niko Välimäki | University of Helsinki, Helsinki, Finland | | Clinical Groups | **Women's Health and Reproduction Group** |
| Eija Laakkonen | University of Jyväskylä, Jyväskylä, Finland | | Clinical Groups | **Women's Health and Reproduction Group** |
| Jaakko Tyrmi | University of Oulu, Oulu, Finland / University of Tampere, Tampere, Finland | | Clinical Groups | **Women's Health and Reproduction Group** |
| Heidi Silven | University of Oulu, Oulu, Finland | | Clinical Groups | **Women's Health and Reproduction Group** |
| Eeva Sliz | University of Oulu, Oulu, Finland | | Clinical Groups | **Women's Health and Reproduction Group** |
| Riikka Arffman | University of Oulu, Oulu, Finland | | Clinical Groups | **Women's Health and Reproduction Group** |
| Susanna Savukoski | University of Oulu, Oulu, Finland | | Clinical Groups | **Women's Health and Reproduction Group** |
| Triin Laisk | Estonian biobank, Tartu, Estonia | | Clinical Groups | **Women's Health and Reproduction Group** |
| Natalia Pujol | Estonian biobank, Tartu, Estonia | | Clinical Groups | **Women's Health and Reproduction Group** |

| | | | | |
|---|---|---|---|---|
| Mengzhen Liu | Abbvie, Chicago, IL, United States | | Clinical Groups | **Women's Health and Reproduction Group** |
| Bridget Riley-Gillis | Abbvie, Chicago, IL, United States | | Clinical Groups | **Women's Health and Reproduction Group** |
| Rion Pendergrass | Genentech, San Francisco, CA, United States | | Clinical Groups | **Women's Health and Reproduction Group** |
| Janet Kumar | GlaxoSmithKline, Collegeville, PA, United States | | Clinical Groups | **Women's Health and Reproduction Group** |
| Kirsi Auro | GlaxoSmithKline, Espoo, Finland | | Clinical Groups | **Women's Health and Reproduction Group** |
| Iiris Hovatta | University of Helsinki, Finland | | Clinical Groups | **Depression group** |
| Chia-Yen Chen | Biogen, Cambridge, MA, United States | | Clinical Groups | **Depression group** |
| Erkki Isometsä | Hospital District of Helsinki and Uusimaa, Helsinki, Finland | | Clinical Groups | **Depression group** |
| Hanna Ollila | Institute for Molecular Medicine Finland (FIMM), HiLIFE, University of Helsinki, Helsinki, Finland | | Clinical Groups | **Depression group** |
| Jaana Suvisaari | Finnish Institute for Health and Welfare (THL), Helsinki, Finland | | Clinical Groups | **Depression group** |
| Antti Mäkitie | Department of Otorhinolaryngology - Head and Neck Surgery, University of Helsinki and Helsinki University Hospital, Helsinki, Finland | | Clinical Groups | **ENT (ear, nose and throath) Group** |
| Argyro Bizaki-Vallaskangas | Pirkanmaa Hospital District, Tampere, Finland | | Clinical Groups | **ENT (ear, nose and throath) Group** |
| Sanna Toppila-Salmi | University of Eastern Finland and Kuopio University Hospital, Department of Otorhinolaryngology, Kuopio, Finland and Department of Allergy, Helsinki University Hospital and University of Helsinki, Finland | | Clinical Groups | **ENT (ear, nose and throath) Group** |
| Tytti Willberg | Hospital District of Southwest Finland, Turku, Finland | | Clinical Groups | **ENT (ear, nose and throath) Group** |
| Elmo Saarentaus | Institute for Molecular Medicine Finland (FIMM), HiLIFE, University of Helsinki, Helsinki, Finland | | Clinical Groups | **ENT (ear, nose and throath) Group** |
| Antti Aarnisalo | Hospital District of Helsinki and Uusimaa, Helsinki, Finland | | Clinical Groups | **ENT (ear, nose and throath) Group** |
| Eveliina Salminen | Hospital District of Helsinki and Uusimaa, Helsinki, Finland | | Clinical Groups | **ENT (ear, nose and throath) Group** |
| Elisa Rahikkala | Northern Ostrobothnia Hospital District, Oulu, Finland | | Clinical Groups | **ENT (ear, nose and throath) Group** |
| Johannes Kettunen | Northern Ostrobothnia Hospital District, Oulu, Finland | | Clinical Groups | **ENT (ear, nose and throath) Group** |
| Kristiina Aittomäki | Department of Medical Genetics, Helsinki University Central Hospital, Helsinki, Finland | | Clinical Groups | **POI (premature ovarian failure) Group** |
| Fredrik Åberg | Transplantation and Liver Surgery Clinic, Helsinki University Hospital, Helsinki University, Helsinki, Finland | | Clinical Groups | **LiverScore Group** |
| Mitja Kurki | Institute for Molecular Medicine Finland (FIMM), HiLIFE, University of Helsinki, Helsinki, Finland; Broad Institute, Cambridge, MA, United States | | FinnGen Analysis working group | FinnGen Analysis working group |
| Samuli Ripatti | Institute for Molecular Medicine Finland (FIMM), HiLIFE, University of Helsinki, Helsinki, Finland | | FinnGen Analysis working group | FinnGen Analysis working group |
| Mark Daly | Institute for Molecular Medicine, Finland (FIMM), HiLIFE, University of Helsinki, Helsinki, Finland; Broad Institute of MIT and Harvard; Massachusetts General Hospital | | FinnGen Analysis working group | FinnGen Analysis working group |
| Juha Karjalainen | Institute for Molecular Medicine Finland (FIMM), HiLIFE, University of Helsinki, Helsinki, Finland | | FinnGen Analysis working group | FinnGen Analysis working group |
| Aki Havulinna | Institute for Molecular Medicine Finland (FIMM), HiLIFE, University of Helsinki, Helsinki, Finland; Finnish Institute for Health and Welfare (THL), Helsinki, Finland | | FinnGen Analysis working group | FinnGen Analysis working group |
| Juha Mehtonen | Institute for Molecular Medicine Finland (FIMM), HiLIFE, University of Helsinki, Helsinki, Finland | | FinnGen Analysis working group | FinnGen Analysis working group |
| Priit Palta | Institute for Molecular Medicine Finland (FIMM), HiLIFE, University of Helsinki, Helsinki, Finland | | FinnGen Analysis working group | FinnGen Analysis working group |
| Shabbeer Hassan | Institute for Molecular Medicine Finland (FIMM), HiLIFE, University of Helsinki, Helsinki, Finland | | FinnGen Analysis working group | FinnGen Analysis working group |
| Pietro Della Briotta Parolo | Institute for Molecular Medicine Finland (FIMM), HiLIFE, University of Helsinki, Helsinki, Finland | | FinnGen Analysis working group | FinnGen Analysis working group |
| Wei Zhou | Broad Institute, Cambridge, MA, United States | | FinnGen Analysis working group | FinnGen Analysis working group |
| Mutaamba Maasha | Broad Institute, Cambridge, MA, United States | | FinnGen Analysis working group | FinnGen Analysis working group |
| Shabbeer Hassan | Institute for Molecular Medicine Finland (FIMM), HiLIFE, University of Helsinki, Helsinki, Finland | | FinnGen Analysis working group | FinnGen Analysis working group |
| Susanna Lemmelä | Institute for Molecular Medicine Finland (FIMM), HiLIFE, University of Helsinki, Helsinki, Finland | | FinnGen Analysis working group | FinnGen Analysis working group |
| Manuel Rivas | University of Stanford, Stanford, CA, United States | | FinnGen Analysis working group | FinnGen Analysis working group |
| Aarno Palotie | Institute for Molecular Medicine Finland (FIMM), HiLIFE, University of Helsinki, Helsinki, Finland | | FinnGen Analysis working group | FinnGen Analysis working group |
| Aoxing Liu | Institute for Molecular Medicine Finland (FIMM), HiLIFE, University of Helsinki, Helsinki, Finland | | FinnGen Analysis working group | FinnGen Analysis working group |
| Arto Lehisto | Institute for Molecular Medicine Finland (FIMM), HiLIFE, University of Helsinki, Helsinki, Finland | | FinnGen Analysis working group | FinnGen Analysis working group |
| Andrea Ganna | Institute for Molecular Medicine Finland (FIMM), HiLIFE, University of Helsinki, Helsinki, Finland | | FinnGen Analysis working group | FinnGen Analysis working group |

| | | | | |
|---|---|---|---|---|
| Vincent Llorens | Institute for Molecular Medicine Finland (FIMM), HiLIFE, University of Helsinki, Helsinki, Finland | | FinnGen Analysis working group | FinnGen Analysis working group |
| Hannele Laivuori | Institute for Molecular Medicine Finland (FIMM), HiLIFE, University of Helsinki, Helsinki, Finland | | FinnGen Analysis working group | FinnGen Analysis working group |
| Taru Tukiainen | Institute for Molecular Medicine Finland (FIMM), HiLIFE, University of Helsinki, Helsinki, Finland | | FinnGen Analysis working group | FinnGen Analysis working group |
| Mary Pat Reeve | Institute for Molecular Medicine Finland (FIMM), HiLIFE, University of Helsinki, Helsinki, Finland | | FinnGen Analysis working group | FinnGen Analysis working group |
| Henrike Heyne | Institute for Molecular Medicine Finland (FIMM), HiLIFE, University of Helsinki, Helsinki, Finland | | FinnGen Analysis working group | FinnGen Analysis working group |
| Nina Mars | Institute for Molecular Medicine Finland (FIMM), HiLIFE, University of Helsinki, Helsinki, Finland | | FinnGen Analysis working group | FinnGen Analysis working group |
| Joel Rämö | Institute for Molecular Medicine Finland (FIMM), HiLIFE, University of Helsinki, Helsinki, Finland | | FinnGen Analysis working group | FinnGen Analysis working group |
| Elmo Saarentaus | Institute for Molecular Medicine Finland (FIMM), HiLIFE, University of Helsinki, Helsinki, Finland | | FinnGen Analysis working group | FinnGen Analysis working group |
| Hanna Ollila | Institute for Molecular Medicine Finland (FIMM), HiLIFE, University of Helsinki, Helsinki, Finland | | FinnGen Analysis working group | FinnGen Analysis working group |
| Rodos Rodosthenous | Institute for Molecular Medicine Finland (FIMM), HiLIFE, University of Helsinki, Helsinki, Finland | | FinnGen Analysis working group | FinnGen Analysis working group |
| Satu Strausz | Institute for Molecular Medicine Finland (FIMM), HiLIFE, University of Helsinki, Helsinki, Finland | | FinnGen Analysis working group | FinnGen Analysis working group |
| Tuula Palotie | University of Helsinki and Hospital District of Helsinki and Uusimaa, Helsinki, Finland | | FinnGen Analysis working group | FinnGen Analysis working group |
| Kimmo Palin | University of Helsinki, Helsinki, Finland | | FinnGen Analysis working group | FinnGen Analysis working group |
| Javier Garcia-Tabuenca | University of Tampere, Tampere, Finland | | FinnGen Analysis working group | FinnGen Analysis working group |
| Harri Siirtola | University of Tampere, Tampere, Finland | | FinnGen Analysis working group | FinnGen Analysis working group |
| Tuomo Kiiskinen | Institute for Molecular Medicine Finland (FIMM), HiLIFE, University of Helsinki, Helsinki, Finland | | FinnGen Analysis working group | FinnGen Analysis working group |
| Jiwoo Lee | Institute for Molecular Medicine Finland (FIMM), HiLIFE, University of Helsinki, Helsinki, Finland; Broad Institute, Cambridge, MA, United States | | FinnGen Analysis working group | FinnGen Analysis working group |
| Kristin Tsuo | Institute for Molecular Medicine Finland (FIMM), HiLIFE, University of Helsinki, Helsinki, Finland; Broad Institute, Cambridge, MA, United States | | FinnGen Analysis working group | FinnGen Analysis working group |
| Amanda Elliott | Institute for Molecular Medicine Finland (FIMM), HiLIFE, University of Helsinki, Helsinki, Finland; Broad Institute, Cambridge, MA, USA and Massachusetts General Hospital, Boston, MA, USA | | FinnGen Analysis working group | FinnGen Analysis working group |
| Kati Kristiansson | THL Biobank / Finnish Institute for Health and Welfare (THL), Helsinki, Finland | | FinnGen Analysis working group | FinnGen Analysis working group |
| Mikko Arvas | Finnish Red Cross Blood Service / Finnish Hematology Registry and Clinical Biobank, Helsinki, Finland | | FinnGen Analysis working group | FinnGen Analysis working group |
| Kati Hyvärinen | Finnish Red Cross Blood Service, Helsinki, Finland | | FinnGen Analysis working group | FinnGen Analysis working group |
| Jarmo Ritari | Finnish Red Cross Blood Service, Helsinki, Finland | | FinnGen Analysis working group | FinnGen Analysis working group |
| Olli Carpén | Helsinki Biobank / Helsinki University and Hospital District of Helsinki and Uusimaa, Helsinki | | FinnGen Analysis working group | FinnGen Analysis working group |
| Johannes Kettunen | Northern Finland Biobank Borealis / University of Oulu / Northern Ostrobothnia Hospital District, Oulu, Finland | | FinnGen Analysis working group | FinnGen Analysis working group |
| Katri Pylkäs | University of Oulu, Oulu, Finland | | FinnGen Analysis working group | FinnGen Analysis working group |
| Eeva Sliz | University of Oulu, Oulu, Finland | | FinnGen Analysis working group | FinnGen Analysis working group |
| Minna Karjalainen | University of Oulu, Oulu, Finland | | FinnGen Analysis working group | FinnGen Analysis working group |
| Tuomo Mantere | Northern Finland Biobank Borealis / University of Oulu / Northern Ostrobothnia Hospital District, Oulu, Finland | | FinnGen Analysis working group | FinnGen Analysis working group |
| Eeva Kangasniemi | Finnish Clinical Biobank Tampere / University of Tampere / Pirkanmaa Hospital District, Tampere, Finland | | FinnGen Analysis working group | FinnGen Analysis working group |
| Sami Heikkinen | University of Eastern Finland, Kuopio, Finland | | FinnGen Analysis working group | FinnGen Analysis working group |
| Arto Mannermaa | Biobank of Eastern Finland / University of Eastern Finland / Northern Savo Hospital District, Kuopio, Finland | | FinnGen Analysis working group | FinnGen Analysis working group |
| Eija Laakkonen | University of Jyväskylä, Jyväskylä, Finland | | FinnGen Analysis working group | FinnGen Analysis working group |
| Nina Pitkänen | Auria Biobank / University of Turku / Hospital District of Southwest Finland, Turku, Finland | | FinnGen Analysis working group | FinnGen Analysis working group |
| Samuel Lessard | Translational Sciences, Sanofi R&D, Framingham, MA, USA | | FinnGen Analysis working group | FinnGen Analysis working group |

| | | | | |
|---|---|---|---|---|
| Clément Chatelain | Translational Sciences, Sanofi R&D, Framingham, MA, USA | | FinnGen Analysis working group | FinnGen Analysis working group |
| Lila Kallio | Auria Biobank / University of Turku / Hospital District of Southwest Finland, Turku, Finland | | Biobank directors | Biobank directors |
| Tiina Wahlfors | THL Biobank / Finnish Institute for Health and Welfare (THL), Helsinki, Finland | | Biobank directors | Biobank directors |
| Jukka Partanen | Finnish Red Cross Blood Service / Finnish Hematology Registry and Clinical Biobank, Helsinki, Finland | | Biobank directors | Biobank directors |
| Eero Punkka | Helsinki Biobank / Helsinki University and Hospital District of Helsinki and Uusimaa, Helsinki | | Biobank directors | Biobank directors |
| Raisa Serpi | Northern Finland Biobank Borealis / University of Oulu / Northern Ostrobothnia Hospital District, Oulu, Finland | | Biobank directors | Biobank directors |
| Sanna Siltanen | Finnish Clinical Biobank Tampere / University of Tampere / Pirkanmaa Hospital District, Tampere, Finland | | Biobank directors | Biobank directors |
| Veli-Matti Kosma | Biobank of Eastern Finland / University of Eastern Finland / Northern Savo Hospital District, Kuopio, Finland | | Biobank directors | Biobank directors |
| Teijo Kuopio | Central Finland Biobank / University of Jyväskylä / Central Finland Health Care District, Jyväskylä, Finland | | Biobank directors | Biobank directors |
| Anu Jalanko | Institute for Molecular Medicine Finland (FIMM), HiLIFE, University of Helsinki, Helsinki, Finland | | FinnGen Teams | **Administration** |
| Huei-Yi Shen | Institute for Molecular Medicine Finland (FIMM), HiLIFE, University of Helsinki, Helsinki, Finland | | FinnGen Teams | **Administration** |
| Risto Kajanne | Institute for Molecular Medicine Finland (FIMM), HiLIFE, University of Helsinki, Helsinki, Finland | | FinnGen Teams | **Administration** |
| Mervi Aavikko | Institute for Molecular Medicine Finland (FIMM), HiLIFE, University of Helsinki, Helsinki, Finland | | FinnGen Teams | **Administration** |
| Helen Cooper | Institute for Molecular Medicine Finland (FIMM), HiLIFE, University of Helsinki, Helsinki, Finland | | FinnGen Teams | **Administration** |
| Denise Öller | Institute for Molecular Medicine Finland (FIMM), HiLIFE, University of Helsinki, Helsinki, Finland | | FinnGen Teams | **Administration** |
| Rasko Leinonen | Institute for Molecular Medicine Finland (FIMM), HiLIFE, University of Helsinki, Helsinki, Finland; European Molecular Biology Laboratory, European Bioinformatics Institute, Cambridge, UK | | FinnGen Teams | **Administration** |
| Henna Palin | Finnish Clinical Biobank Tampere / University of Tampere / Pirkanmaa Hospital District, Tampere, Finland | | FinnGen Teams | **Administration** |
| Malla-Maria Linna | Helsinki Biobank / Helsinki University and Hospital District of Helsinki and Uusimaa, Helsinki | | FinnGen Teams | **Administration** |
| Mitja Kurki | Institute for Molecular Medicine Finland (FIMM), HiLIFE, University of Helsinki, Helsinki, Finland; Broad Institute, Cambridge, MA, United States | | FinnGen Teams | **Analysis** |
| Juha Karjalainen | Institute for Molecular Medicine Finland (FIMM), HiLIFE, University of Helsinki, Helsinki, Finland | | FinnGen Teams | **Analysis** |
| Pietro Della Briotta Parolo | Institute for Molecular Medicine Finland (FIMM), HiLIFE, University of Helsinki, Helsinki, Finland | | FinnGen Teams | **Analysis** |
| Arto Lehisto | Institute for Molecular Medicine Finland (FIMM), HiLIFE, University of Helsinki, Helsinki, Finland | | FinnGen Teams | **Analysis** |
| Juha Mehtonen | Institute for Molecular Medicine Finland (FIMM), HiLIFE, University of Helsinki, Helsinki, Finland | | FinnGen Teams | **Analysis** |
| Wei Zhou | Broad Institute, Cambridge, MA, United States | | FinnGen Teams | **Analysis** |
| Masahiro Kanai | Broad Institute, Cambridge, MA, United States | | FinnGen Teams | **Analysis** |
| Mutaamba Maasha | Broad Institute, Cambridge, MA, United States | | FinnGen Teams | **Analysis** |
| Zhili Zheng | Broad Institute, Cambridge, MA, United States | | FinnGen Teams | **Analysis** |
| Hannele Laivuori | Institute for Molecular Medicine Finland (FIMM), HiLIFE, University of Helsinki, Helsinki, Finland | | FinnGen Teams | **Clinical Endpoint Development** |
| Aki Havulinna | Institute for Molecular Medicine Finland (FIMM), HiLIFE, University of Helsinki, Helsinki, Finland; Finnish Institute for Health and Welfare (THL), Helsinki, Finland | | FinnGen Teams | **Clinical Endpoint Development** |
| Susanna Lemmelä | Institute for Molecular Medicine Finland (FIMM), HiLIFE, University of Helsinki, Helsinki, Finland | | FinnGen Teams | **Clinical Endpoint Development** |
| Tuomo Kiiskinen | Institute for Molecular Medicine Finland (FIMM), HiLIFE, University of Helsinki, Helsinki, Finland | | FinnGen Teams | **Clinical Endpoint Development** |
| L. Elisa Lahtela | Institute for Molecular Medicine Finland (FIMM), HiLIFE, University of Helsinki, Helsinki, Finland | | FinnGen Teams | **Clinical Endpoint Development** |
| Mari Kaunisto | Institute for Molecular Medicine Finland (FIMM), HiLIFE, University of Helsinki, Helsinki, Finland | | FinnGen Teams | **Communication** |
| Elina Kilpeläinen | Institute for Molecular Medicine Finland (FIMM), HiLIFE, University of Helsinki, Helsinki, Finland | | FinnGen Teams | **E-Science** |
| Timo P. Sipilä | Institute for Molecular Medicine Finland (FIMM), HiLIFE, University of Helsinki, Helsinki, Finland | | FinnGen Teams | **E-Science** |

| | | | | |
|---|---|---|---|---|
| Oluwaseun Alexander Dada | Institute for Molecular Medicine Finland (FIMM), HiLIFE, University of Helsinki, Helsinki, Finland | | FinnGen Teams | **E-Science** |
| Awaisa Ghazal | Institute for Molecular Medicine Finland (FIMM), HiLIFE, University of Helsinki, Helsinki, Finland | | FinnGen Teams | **E-Science** |
| Anastasia Kytölä | Institute for Molecular Medicine Finland (FIMM), HiLIFE, University of Helsinki, Helsinki, Finland | | FinnGen Teams | **E-Science** |
| Rigbe Weldatsadik | Institute for Molecular Medicine Finland (FIMM), HiLIFE, University of Helsinki, Helsinki, Finland | | FinnGen Teams | **E-Science** |
| Sanni Ruotsalainen | Institute for Molecular Medicine Finland (FIMM), HiLIFE, University of Helsinki, Helsinki, Finland | | FinnGen Teams | **E-Science** |
| Kati Donner | Institute for Molecular Medicine Finland (FIMM), HiLIFE, University of Helsinki, Helsinki, Finland | | FinnGen Teams | **Genotyping** |
| Timo P. Sipilä | Institute for Molecular Medicine Finland (FIMM), HiLIFE, University of Helsinki, Helsinki, Finland | | FinnGen Teams | **Genotyping** |
| Anu Loukola | Helsinki Biobank / Helsinki University and Hospital District of Helsinki and Uusimaa, Helsinki | | FinnGen Teams | **Sample Collection Coordination** |
| Päivi Laiho | THL Biobank / Finnish Institute for Health and Welfare (THL), Helsinki, Finland | | FinnGen Teams | **Sample Logistics** |
| Tuuli Sistonen | THL Biobank / Finnish Institute for Health and Welfare (THL), Helsinki, Finland | | FinnGen Teams | **Sample Logistics** |
| Essi Kaiharju | THL Biobank / Finnish Institute for Health and Welfare (THL), Helsinki, Finland | | FinnGen Teams | **Sample Logistics** |
| Markku Laukkanen | THL Biobank / Finnish Institute for Health and Welfare (THL), Helsinki, Finland | | FinnGen Teams | **Sample Logistics** |
| Elina Järvensivu | THL Biobank / Finnish Institute for Health and Welfare (THL), Helsinki, Finland | | FinnGen Teams | **Sample Logistics** |
| Sini Lähteenmäki | THL Biobank / Finnish Institute for Health and Welfare (THL), Helsinki, Finland | | FinnGen Teams | **Sample Logistics** |
| Lotta Männikkö | THL Biobank / Finnish Institute for Health and Welfare (THL), Helsinki, Finland | | FinnGen Teams | **Sample Logistics** |
| Regis Wong | THL Biobank / Finnish Institute for Health and Welfare (THL), Helsinki, Finland | | FinnGen Teams | **Sample Logistics** |
| Auli Toivola | THL Biobank / Finnish Institute for Health and Welfare (THL), Helsinki, Finland | | FinnGen Teams | **Sample Logistics** |
| Minna Brunfeldt | THL Biobank / Finnish Institute for Health and Welfare (THL), Helsinki, Finland | | FinnGen Teams | **Registry Data Operations** |
| Hannele Mattsson | THL Biobank / Finnish Institute for Health and Welfare (THL), Helsinki, Finland | | FinnGen Teams | **Registry Data Operations** |
| Kati Kristiansson | THL Biobank / Finnish Institute for Health and Welfare (THL), Helsinki, Finland | | FinnGen Teams | **Registry Data Operations** |
| Susanna Lemmelä | Institute for Molecular Medicine Finland (FIMM), HiLIFE, University of Helsinki, Helsinki, Finland | | FinnGen Teams | **Registry Data Operations** |
| Sami Koskelainen | THL Biobank / Finnish Institute for Health and Welfare (THL), Helsinki, Finland | | FinnGen Teams | **Registry Data Operations** |
| Tero Hiekkalinna | THL Biobank / Finnish Institute for Health and Welfare (THL), Helsinki, Finland | | FinnGen Teams | **Registry Data Operations** |
| Teemu Paajanen | THL Biobank / Finnish Institute for Health and Welfare (THL), Helsinki, Finland | | FinnGen Teams | **Registry Data Operations** |
| Priit Palta | Institute for Molecular Medicine Finland (FIMM), HiLIFE, University of Helsinki, Helsinki, Finland | | FinnGen Teams | **Sequencing Informatics** |
| Shuang Luo | Institute for Molecular Medicine Finland (FIMM), HiLIFE, University of Helsinki, Helsinki, Finland | | FinnGen Teams | **Sequencing Informatics** |
| Tarja Laitinen | Pirkanmaa Hospital District, Tampere, Finland | | FinnGen Teams | **Trajectory** |
| Mary Pat Reeve | Institute for Molecular Medicine Finland (FIMM), HiLIFE, University of Helsinki, Helsinki, Finland | | FinnGen Teams | **Trajectory** |
| Shanmukha Sampath Padmanabhuni | Institute for Molecular Medicine Finland (FIMM), HiLIFE, University of Helsinki, Helsinki, Finland | | FinnGen Teams | **Trajectory** |
| Marianna Niemi | University of Tampere, Tampere, Finland | | FinnGen Teams | **Trajectory** |
| Harri Siirtola | University of Tampere, Tampere, Finland | | FinnGen Teams | **Trajectory** |
| Javier Gracia-Tabuenca | University of Tampere, Tampere, Finland | | FinnGen Teams | **Trajectory** |
| Mika Helminen | University of Tampere, Tampere, Finland | | FinnGen Teams | **Trajectory** |
| Tiina Luukkaala | University of Tampere, Tampere, Finland | | FinnGen Teams | **Trajectory** |
| Iida Vähätalo | University of Tampere, Tampere, Finland | | FinnGen Teams | **Trajectory** |
| Jyrki Tammerluoto | Institute for Molecular Medicine Finland (FIMM), HiLIFE, University of Helsinki, Helsinki, Finland | | FinnGen Teams | **Data protection officer** |
| Marco Hautalahti | Finnish Biobank Cooperative - FINBB | | FinnGen Teams | **FINBB - Finnish biobank cooperative** |
| Johanna Mäkelä | Finnish Biobank Cooperative - FINBB | | FinnGen Teams | **FINBB - Finnish biobank cooperative** |
| Sarah Smith | Finnish Biobank Cooperative - FINBB | | FinnGen Teams | **FINBB - Finnish biobank cooperative** |
| Tom Southerington | Finnish Biobank Cooperative - FINBB | | FinnGen Teams | **FINBB - Finnish biobank cooperative** |
| Petri Lehto | Finnish Biobank Cooperative - FINBB | | FinnGen Teams | **FINBB - Finnish biobank cooperative** |

**Table S2. Analysis subgroup definitions.**

Table shows case and control definitions, possible analysis exclusions, as well as amount of cases and controls both in FinnGen and Estonian Biobank (EstBB) populations as well as in meta-analysis.

*Pneumonia diagnosis codes:

**ICD-10:** J10.0, J11.0, J12, J12.0, J12.1, J12.2, J12.3, J12.8, J12.9, J13, J14, J15.0, J15.1, J15.2, J15.3, J15.4, J15.5, J15.6, J15.7, J15.8, J15.9, J16.0, J16.8, J18.0, J18.1, J18.2, J18.8, J18.9

**ICD-9:** 480, 481, 482, 483, 485, 4822, 4810A, 4820A, 4821A, 4822A, 4823A, 4824A, 4828X, 4870A

**ICD-8:** 482, 486, 47101, 47109, 48099, 48199, 48201, 48210, 48220, 48230, 48298, 48399, 48502, 48509

**Asthma diagnosis codes:

**ICD-10:** J45.0, J45.1, J45.8, J45.9, J46

**ICD-9:** 493

**ICD-8:** 493

| Subgroup | Case definition | Control definition | Exclusions | Cases, FinnGen | Cases, EstBB | Cases, meta | Controls, FinnGen | Controls, EstBB | Controls, meta |
|---|---|---|---|---|---|---|---|---|---|
| Pneumonia, unselected | Individuals with pneumonia diagnosis* at least once during lifetime | Individuals without pneumonia diagnosis * | None | 64 635 | 26 427 | 91 062 | 347 546 | 172 573 | 520 119 |
| Age under 16 | Individuals with pneumonia diagnosis* < 16 years old. In case of multiple diagnoses during lifetime, age group is defined as earliest diagnosis made. | Individuals without pneumonia diagnosis * | Individuals who have had pneumonia diagnosis* at ≥ 16 years of age. | 3193 | 3 448 | 6 641 | 347 546 | 172 894 | 520 440 |
| Age 16-60 | Individuals with pneumonia diagnosis* at 16 ≤ diagnosis age < 60. | Individuals without pneumonia diagnosis * | Individuals who have had pneumonia diagnosis* at < 16 or ≥ 60 years of age. | 24 032 | 16 404 | 40 436 | 347 546 | 172 894 | 520 440 |
| Age over 60 | Individuals with pneumonia diagnosis* at ≥ 60 years old. | Individuals without pneumonia diagnosis * | Individuals who have had pneumonia diagnosis* at < 60 years of age. | 37 410 | 6 643 | 44 053 | 347 546 | 172 894 | 520 440 |
| Recurrent pneumonia | Individuals with 3 or more pneumonia episodes during lifetime. To consider two given pneumonia diagnoses* as separate episodes, there must have been a minimum of 3 months (90 days) between diagnoses. | Individuals without pneumonia diagnosis * | Individuals with 1 or 2 pneumonia diagnoses* | 6 903 | 1 296 | 8 199 | 347 546 | 176 625 | 524 171 |
| Pneumonia in asthma patients | Individuals with asthma diagnosis**, who have also had at least one pneumonia diagnosis*. | Individuals with asthma diagnosis**, but without pneumonia diagnosis*. | Individuals without asthma diagnosis**. | 14 234 | 5 286 | 19 520 | 36 695 | 16 517 | 53 212 |

**Table S3. Lead variants in FinnGen pneumonia inpatient analysis.**

Column header abbreviations: chrom: chromosome, pos: position, ref: reference allele, alt: alternate allele (effect allele), rsid: rsid-identifier for the variant, nearest_gene: neareset gene to the variant, pval: p-value of the association, mlogp: -log10(p-value), beta: effect size, sebeta: beta standard error, af_alt: alternate allele frequency, af_alt_cases: alternate allele frequency in cases, af_alt_controls: alternate allele frequency in controls.

| chrom | pos | ref | alt | rsid | nearest_gene | pval | mlogp | beta | sebeta | af_alt | af_alt_cases | af_alt_controls |
|---|---|---|---|---|---|---|---|---|---|---|---|---|
| 1 | 113817445 | T | G | rs11102694 | BCL2L15 | 2.78E-10 | 9.56 | -0.0484 | 0.0077 | 0.653 | 0.645 | 0.654 |
| 1 | 159714396 | T | A | rs3116636 | CRP | 5.83E-16 | 15.23 | 0.0616 | 0.0076 | 0.353 | 0.366 | 0.351 |
| 6 | 80321027 | C | T | rs700491 | BCKDHB | 3.15E-08 | 7.50 | -0.0528 | 0.0095 | 0.824 | 0.818 | 0.825 |
| 14 | 91922703 | C | T | rs2284340 | FBLN5 | 2.42E-08 | 7.62 | -0.0411 | 0.0074 | 0.466 | 0.458 | 0.467 |
| 15 | 78593885 | A | G | rs564585 | CHRNA3/CHRNA5 | 2.01E-09 | 8.70 | -0.0477 | 0.0079 | 0.311 | 0.301 | 0.312 |

**Table S4. Significance of effect estimate differences in non-HLA lead variants across analyses.** To determine if effect estimates in lead variants differ significantly between subgroups, we conducted a two-tailed significance test between all subgroups. Comparisons with p_diff < 0.05 were considered significant and are highlighted in green color.

| Lead variant | Subgroup 1 | Subgroup2 | Subgroup1_Effect | Subgroup1 _StdErr | Subgroup2_Effect | Subgroup2_ StdErr | z_diff | p_diff |
|---|---|---|---|---|---|---|---|---|
| PTPN22 rs11102694 - A | General | Under 16 year olds | 0.035 | 0.006 | 0.013 | 0.021 | 1.006 | 0.314 |
| PTPN22 rs11102694 - A | General | 16-60 year olds | 0.035 | 0.006 | 0.043 | 0.009 | -0.787 | 0.431 |
| PTPN22 rs11102694 - A | General | Over 60 year olds | 0.035 | 0.006 | 0.035 | 0.009 | 0.018 | 0.985 |
| PTPN22 rs11102694 - A | General | Recurrent | 0.035 | 0.006 | 0.073 | 0.019 | -1.951 | 0.051 |
| PTPN22 rs11102694 - A | General | Asthma | 0.035 | 0.006 | 0.053 | 0.014 | -1.119 | 0.263 |
| PTPN22 rs11102694 - A | Under 16 year olds | 16-60 year olds | 0.013 | 0.021 | 0.043 | 0.009 | -1.334 | 0.182 |
| PTPN22 rs11102694 - A | Under 16 year olds | Over 60 year olds | 0.013 | 0.021 | 0.035 | 0.009 | -0.953 | 0.341 |
| PTPN22 rs11102694 - A | Under 16 year olds | Recurrent | 0.013 | 0.021 | 0.073 | 0.019 | -2.144 | 0.032 |
| PTPN22 rs11102694 - A | Under 16 year olds | Asthma | 0.013 | 0.021 | 0.053 | 0.014 | -1.550 | 0.121 |
| PTPN22 rs11102694 - A | 16-60 year olds | Over 60 year olds | 0.043 | 0.009 | 0.035 | 0.009 | 0.679 | 0.497 |
| PTPN22 rs11102694 - A | 16-60 year olds | Recurrent | 0.043 | 0.009 | 0.073 | 0.019 | -1.459 | 0.145 |
| PTPN22 rs11102694 - A | 16-60 year olds | Asthma | 0.043 | 0.009 | 0.053 | 0.014 | -0.549 | 0.583 |
| PTPN22 rs11102694 - A | Over 60 year olds | Recurrent | 0.035 | 0.009 | 0.073 | 0.019 | -1.854 | 0.064 |
| PTPN22 rs11102694 - A | Over 60 year olds | Asthma | 0.035 | 0.009 | 0.053 | 0.014 | -1.039 | 0.299 |
| PTPN22 rs11102694 - A | Recurrent | Asthma | 0.073 | 0.019 | 0.053 | 0.014 | 0.880 | 0.379 |
| CRP rs3116636 - A | General | Under 16 year olds | 0.055 | 0.005 | 0.034 | 0.019 | 1.058 | 0.290 |
| CRP rs3116636 - A | General | 16-60 year olds | 0.055 | 0.005 | 0.061 | 0.008 | -0.606 | 0.544 |
| CRP rs3116636 - A | General | Over 60 year olds | 0.055 | 0.005 | 0.059 | 0.008 | -0.448 | 0.654 |
| CRP rs3116636 - A | General | Recurrent | 0.055 | 0.005 | 0.098 | 0.017 | -2.480 | 0.013 |
| CRP rs3116636 - A | General | Asthma | 0.055 | 0.005 | 0.079 | 0.013 | -1.673 | 0.094 |
| CRP rs3116636 - A | Under 16 year olds | 16-60 year olds | 0.034 | 0.019 | 0.061 | 0.008 | -1.299 | 0.194 |
| CRP rs3116636 - A | Under 16 year olds | Over 60 year olds | 0.034 | 0.019 | 0.059 | 0.008 | -1.224 | 0.221 |
| CRP rs3116636 - A | Under 16 year olds | Recurrent | 0.034 | 0.019 | 0.098 | 0.017 | -2.552 | 0.011 |
| CRP rs3116636 - A | Under 16 year olds | Asthma | 0.034 | 0.019 | 0.079 | 0.013 | -1.938 | 0.053 |
| CRP rs3116636 - A | 16-60 year olds | Over 60 year olds | 0.061 | 0.008 | 0.059 | 0.008 | 0.116 | 0.908 |
| CRP rs3116636 - A | 16-60 year olds | Recurrent | 0.061 | 0.008 | 0.098 | 0.017 | -2.055 | 0.040 |
| CRP rs3116636 - A | 16-60 year olds | Asthma | 0.061 | 0.008 | 0.079 | 0.013 | -1.185 | 0.236 |
| CRP rs3116636 - A | Over 60 year olds | Recurrent | 0.059 | 0.008 | 0.098 | 0.017 | -2.101 | 0.036 |
| CRP rs3116636 - A | Over 60 year olds | Asthma | 0.059 | 0.008 | 0.079 | 0.013 | -1.249 | 0.212 |
| CRP rs3116636 - A | Recurrent | Asthma | 0.098 | 0.017 | 0.079 | 0.013 | 0.927 | 0.354 |
| MAPKAPK2 rs61815610 - T | General | Under 16 year olds | 0.030 | 0.007 | -0.001 | 0.025 | 1.183 | 0.237 |

| | | | | | | | | |
|---|---|---|---|---|---|---|---|---|
| MAPKAPK2 rs61815610 - T | General | 16-60 year olds | 0.030 | 0.007 | 0.056 | 0.010 | -2.139 | 0.032 |
| MAPKAPK2 rs61815610 - T | General | Over 60 year olds | 0.030 | 0.007 | 0.012 | 0.011 | 1.341 | 0.180 |
| MAPKAPK2 rs61815610 - T | General | Recurrent | 0.030 | 0.007 | -0.004 | 0.022 | 1.447 | 0.148 |
| MAPKAPK2 rs61815610 - T | General | Asthma | 0.030 | 0.007 | 0.024 | 0.017 | 0.290 | 0.772 |
| MAPKAPK2 rs61815610 - T | Under 16 year olds | 16-60 year olds | -0.001 | 0.025 | 0.056 | 0.010 | -2.126 | 0.034 |
| MAPKAPK2 rs61815610 - T | Under 16 year olds | Over 60 year olds | -0.001 | 0.025 | 0.012 | 0.011 | -0.494 | 0.621 |
| MAPKAPK2 rs61815610 - T | Under 16 year olds | Recurrent | -0.001 | 0.025 | -0.004 | 0.022 | 0.089 | 0.929 |
| MAPKAPK2 rs61815610 - T | Under 16 year olds | Asthma | -0.001 | 0.025 | 0.024 | 0.017 | -0.838 | 0.402 |
| MAPKAPK2 rs61815610 - T | 16-60 year olds | Over 60 year olds | 0.056 | 0.010 | 0.012 | 0.011 | 2.969 | 0.003 |
| MAPKAPK2 rs61815610 - T | 16-60 year olds | Recurrent | 0.056 | 0.010 | -0.004 | 0.022 | 2.471 | 0.013 |
| MAPKAPK2 rs61815610 - T | 16-60 year olds | Asthma | 0.056 | 0.010 | 0.024 | 0.017 | 1.605 | 0.108 |
| MAPKAPK2 rs61815610 - T | Over 60 year olds | Recurrent | 0.012 | 0.011 | -0.004 | 0.022 | 0.666 | 0.505 |
| MAPKAPK2 rs61815610 - T | Over 60 year olds | Asthma | 0.012 | 0.011 | 0.024 | 0.017 | -0.591 | 0.555 |
| MAPKAPK2 rs61815610 - T | Recurrent | Asthma | -0.004 | 0.022 | 0.024 | 0.017 | -1.012 | 0.312 |
| FHIT rs11130706 - T | General | Under 16 year olds | -0.011 | 0.007 | -0.019 | 0.025 | 0.321 | 0.748 |
| FHIT rs11130706 - T | General | 16-60 year olds | -0.011 | 0.007 | -0.011 | 0.010 | 0.016 | 0.987 |
| FHIT rs11130706 - T | General | Over 60 year olds | -0.011 | 0.007 | -0.010 | 0.011 | -0.008 | 0.994 |
| FHIT rs11130706 - T | General | Recurrent | -0.011 | 0.007 | -0.045 | 0.022 | 1.489 | 0.137 |
| FHIT rs11130706 - T | General | Asthma | -0.011 | 0.007 | -0.096 | 0.017 | 4.663 | 3.12E-06 |
| FHIT rs11130706 - T | Under 16 year olds | 16-60 year olds | -0.019 | 0.025 | -0.011 | 0.010 | -0.302 | 0.763 |
| FHIT rs11130706 - T | Under 16 year olds | Over 60 year olds | -0.019 | 0.025 | -0.010 | 0.011 | -0.310 | 0.756 |
| FHIT rs11130706 - T | Under 16 year olds | Recurrent | -0.019 | 0.025 | -0.045 | 0.022 | 0.802 | 0.423 |
| FHIT rs11130706 - T | Under 16 year olds | Asthma | -0.019 | 0.025 | -0.096 | 0.017 | 2.606 | 0.009 |
| FHIT rs11130706 - T | 16-60 year olds | Over 60 year olds | -0.011 | 0.010 | -0.010 | 0.011 | -0.020 | 0.984 |
| FHIT rs11130706 - T | 16-60 year olds | Recurrent | -0.011 | 0.010 | -0.045 | 0.022 | 1.415 | 0.157 |
| FHIT rs11130706 - T | 16-60 year olds | Asthma | -0.011 | 0.010 | -0.096 | 0.017 | 4.334 | 1.46E-05 |
| FHIT rs11130706 - T | Over 60 year olds | Recurrent | -0.010 | 0.011 | -0.045 | 0.022 | 1.412 | 0.158 |
| FHIT rs11130706 - T | Over 60 year olds | Asthma | -0.010 | 0.011 | -0.096 | 0.017 | 4.281 | 1.86E-05 |
| FHIT rs11130706 - T | Recurrent | Asthma | -0.045 | 0.022 | -0.096 | 0.017 | 1.831 | 0.067 |
| PTGER4 rs7725052 - T | General | Under 16 year olds | 0.029 | 0.005 | 0.056 | 0.018 | -1.422 | 0.155 |
| PTGER4 rs7725052 - T | General | 16-60 year olds | 0.029 | 0.005 | 0.034 | 0.007 | -0.520 | 0.603 |
| PTGER4 rs7725052 - T | General | Over 60 year olds | 0.029 | 0.005 | 0.018 | 0.008 | 1.141 | 0.254 |
| PTGER4 rs7725052 - T | General | Recurrent | 0.029 | 0.005 | 0.044 | 0.016 | -0.892 | 0.373 |
| PTGER4 rs7725052 - T | General | Asthma | 0.029 | 0.005 | 0.025 | 0.012 | 0.283 | 0.777 |
| PTGER4 rs7725052 - T | Under 16 year olds | 16-60 year olds | 0.056 | 0.018 | 0.034 | 0.007 | 1.125 | 0.260 |
| PTGER4 rs7725052 - T | Under 16 year olds | Over 60 year olds | 0.056 | 0.018 | 0.018 | 0.008 | 1.905 | 0.057 |

| | | | | | | | | |
|---|---|---|---|---|---|---|---|---|
| PTGER4 rs7725052 - T | Under 16 year olds | Recurrent | 0.056 | 0.018 | 0.044 | 0.016 | 0.479 | 0.632 |
| PTGER4 rs7725052 - T | Under 16 year olds | Asthma | 0.056 | 0.018 | 0.025 | 0.012 | 1.391 | 0.164 |
| PTGER4 rs7725052 - T | 16-60 year olds | Over 60 year olds | 0.034 | 0.007 | 0.018 | 0.008 | 1.432 | 0.152 |
| PTGER4 rs7725052 - T | 16-60 year olds | Recurrent | 0.034 | 0.007 | 0.044 | 0.016 | -0.584 | 0.559 |
| PTGER4 rs7725052 - T | 16-60 year olds | Asthma | 0.034 | 0.007 | 0.025 | 0.012 | 0.589 | 0.556 |
| PTGER4 rs7725052 - T | Over 60 year olds | Recurrent | 0.018 | 0.008 | 0.044 | 0.016 | -1.444 | 0.149 |
| PTGER4 rs7725052 - T | Over 60 year olds | Asthma | 0.018 | 0.008 | 0.025 | 0.012 | -0.471 | 0.638 |
| PTGER4 rs7725052 - T | Recurrent | Asthma | 0.044 | 0.016 | 0.025 | 0.012 | 0.929 | 0.353 |
| TNFSF15 rs112284124 - A | General | Under 16 year olds | -0.033 | 0.006 | -0.074 | 0.020 | 1.935 | 0.053 |
| TNFSF15 rs112284124 - A | General | 16-60 year olds | -0.033 | 0.006 | -0.039 | 0.008 | 0.546 | 0.585 |
| TNFSF15 rs112284124 - A | General | Over 60 year olds | -0.033 | 0.006 | -0.022 | 0.009 | -1.096 | 0.273 |
| TNFSF15 rs112284124 - A | General | Recurrent | -0.033 | 0.006 | -0.055 | 0.018 | 1.149 | 0.250 |
| TNFSF15 rs112284124 - A | General | Asthma | -0.033 | 0.006 | -0.023 | 0.014 | -0.637 | 0.524 |
| TNFSF15 rs112284124 - A | Under 16 year olds | 16-60 year olds | -0.074 | 0.020 | -0.039 | 0.008 | -1.607 | 0.108 |
| TNFSF15 rs112284124 - A | Under 16 year olds | Over 60 year olds | -0.074 | 0.020 | -0.022 | 0.009 | -2.373 | 0.018 |
| TNFSF15 rs112284124 - A | Under 16 year olds | Recurrent | -0.074 | 0.020 | -0.055 | 0.018 | -0.688 | 0.492 |
| TNFSF15 rs112284124 - A | Under 16 year olds | Asthma | -0.074 | 0.020 | -0.023 | 0.014 | -2.047 | 0.041 |
| TNFSF15 rs112284124 - A | 16-60 year olds | Over 60 year olds | -0.039 | 0.008 | -0.022 | 0.009 | -1.414 | 0.157 |
| TNFSF15 rs112284124 - A | 16-60 year olds | Recurrent | -0.039 | 0.008 | -0.055 | 0.018 | 0.819 | 0.413 |
| TNFSF15 rs112284124 - A | 16-60 year olds | Asthma | -0.039 | 0.008 | -0.023 | 0.014 | -0.933 | 0.351 |
| TNFSF15 rs112284124 - A | Over 60 year olds | Recurrent | -0.022 | 0.009 | -0.055 | 0.018 | 1.661 | 0.097 |
| TNFSF15 rs112284124 - A | Over 60 year olds | Asthma | -0.022 | 0.009 | -0.023 | 0.014 | 0.113 | 0.910 |
| TNFSF15 rs112284124 - A | Recurrent | Asthma | -0.055 | 0.018 | -0.023 | 0.014 | -1.377 | 0.168 |
| MUC5AC** rs28415845 - T | General | Under 16 year olds | 0.037 | 0.006 | 0.057 | 0.020 | -0.967 | 0.333 |
| MUC5AC** rs28415845 - T | General | 16-60 year olds | 0.037 | 0.006 | 0.041 | 0.008 | -0.420 | 0.675 |
| MUC5AC** rs28415845 - T | General | Over 60 year olds | 0.037 | 0.006 | 0.033 | 0.008 | 0.406 | 0.685 |
| MUC5AC** rs28415845 - T | General | Recurrent | 0.037 | 0.006 | 0.101 | 0.017 | -3.585 | 3.38E-04 |
| MUC5AC** rs28415845 - T | General | Asthma | 0.037 | 0.006 | 0.049 | 0.014 | -0.828 | 0.408 |
| MUC5AC** rs28415845 - T | Under 16 year olds | 16-60 year olds | 0.057 | 0.020 | 0.041 | 0.008 | 0.740 | 0.459 |
| MUC5AC** rs28415845 - T | Under 16 year olds | Over 60 year olds | 0.057 | 0.020 | 0.033 | 0.008 | 1.116 | 0.264 |
| MUC5AC** rs28415845 - T | Under 16 year olds | Recurrent | 0.057 | 0.020 | 0.101 | 0.017 | -1.705 | 0.088 |
| MUC5AC** rs28415845 - T | Under 16 year olds | Asthma | 0.057 | 0.020 | 0.049 | 0.014 | 0.325 | 0.745 |
| MUC5AC** rs28415845 - T | 16-60 year olds | Over 60 year olds | 0.041 | 0.008 | 0.033 | 0.008 | 0.707 | 0.480 |
| MUC5AC** rs28415845 - T | 16-60 year olds | Recurrent | 0.041 | 0.008 | 0.101 | 0.017 | -3.199 | 0.001 |
| MUC5AC** rs28415845 - T | 16-60 year olds | Asthma | 0.041 | 0.008 | 0.049 | 0.014 | -0.510 | 0.610 |

| | | | | | | | | |
|---|---|---|---|---|---|---|---|---|
| MUC5AC** rs28415845 - T | Over 60 year olds | Recurrent | 0.033 | 0.008 | 0.101 | 0.017 | -3.601 | 3.17E-04 |
| MUC5AC** rs28415845 - T | Over 60 year olds | Asthma | 0.033 | 0.008 | 0.049 | 0.014 | -1.019 | 0.308 |
| MUC5AC** rs28415845 - T | Recurrent | Asthma | 0.101 | 0.017 | 0.049 | 0.014 | 2.405 | 0.016 |
| TNFRSF1A rs4149581 - T | General | Under 16 year olds | 0.029 | 0.005 | 0.047 | 0.018 | -0.977 | 0.329 |
| TNFRSF1A rs4149581 - T | General | 16-60 year olds | 0.029 | 0.005 | 0.033 | 0.007 | -0.442 | 0.658 |
| TNFRSF1A rs4149581 - T | General | Over 60 year olds | 0.029 | 0.005 | 0.023 | 0.008 | 0.613 | 0.540 |
| TNFRSF1A rs4149581 - T | General | Recurrent | 0.029 | 0.005 | 0.030 | 0.016 | -0.094 | 0.925 |
| TNFRSF1A rs4149581 - T | General | Asthma | 0.029 | 0.005 | 0.031 | 0.013 | -0.170 | 0.865 |
| TNFRSF1A rs4149581 - T | Under 16 year olds | 16-60 year olds | 0.047 | 0.018 | 0.033 | 0.007 | 0.735 | 0.462 |
| TNFRSF1A rs4149581 - T | Under 16 year olds | Over 60 year olds | 0.047 | 0.018 | 0.023 | 0.008 | 1.226 | 0.220 |
| TNFRSF1A rs4149581 - T | Under 16 year olds | Recurrent | 0.047 | 0.018 | 0.030 | 0.016 | 0.690 | 0.490 |
| TNFRSF1A rs4149581 - T | Under 16 year olds | Asthma | 0.047 | 0.018 | 0.031 | 0.013 | 0.730 | 0.465 |
| TNFRSF1A rs4149581 - T | 16-60 year olds | Over 60 year olds | 0.033 | 0.007 | 0.023 | 0.008 | 0.905 | 0.365 |
| TNFRSF1A rs4149581 - T | 16-60 year olds | Recurrent | 0.033 | 0.007 | 0.030 | 0.016 | 0.135 | 0.893 |
| TNFRSF1A rs4149581 - T | 16-60 year olds | Asthma | 0.033 | 0.007 | 0.031 | 0.013 | 0.117 | 0.907 |
| TNFRSF1A rs4149581 - T | Over 60 year olds | Recurrent | 0.023 | 0.008 | 0.030 | 0.016 | -0.411 | 0.681 |
| TNFRSF1A rs4149581 - T | Over 60 year olds | Asthma | 0.023 | 0.008 | 0.031 | 0.013 | -0.548 | 0.584 |
| TNFRSF1A rs4149581 - T | Recurrent | Asthma | 0.030 | 0.016 | 0.031 | 0.013 | -0.034 | 0.973 |
| HNF1A rs58367757 - T | General | Under 16 year olds | 0.031 | 0.005 | 0.030 | 0.018 | 0.047 | 0.963 |
| HNF1A rs58367757 - T | General | 16-60 year olds | 0.031 | 0.005 | 0.037 | 0.008 | -0.664 | 0.507 |
| HNF1A rs58367757 - T | General | Over 60 year olds | 0.031 | 0.005 | 0.018 | 0.008 | 1.386 | 0.166 |
| HNF1A rs58367757 - T | General | Recurrent | 0.031 | 0.005 | 0.052 | 0.016 | -1.189 | 0.234 |
| HNF1A rs58367757 - T | General | Asthma | 0.031 | 0.005 | 0.039 | 0.013 | -0.567 | 0.571 |
| HNF1A rs58367757 - T | Under 16 year olds | 16-60 year olds | 0.030 | 0.018 | 0.037 | 0.008 | -0.352 | 0.725 |
| HNF1A rs58367757 - T | Under 16 year olds | Over 60 year olds | 0.030 | 0.018 | 0.018 | 0.008 | 0.618 | 0.537 |
| HNF1A rs58367757 - T | Under 16 year olds | Recurrent | 0.030 | 0.018 | 0.052 | 0.016 | -0.868 | 0.385 |
| HNF1A rs58367757 - T | Under 16 year olds | Asthma | 0.030 | 0.018 | 0.039 | 0.013 | -0.389 | 0.697 |
| HNF1A rs58367757 - T | 16-60 year olds | Over 60 year olds | 0.037 | 0.008 | 0.018 | 0.008 | 1.769 | 0.077 |
| HNF1A rs58367757 - T | 16-60 year olds | Recurrent | 0.037 | 0.008 | 0.052 | 0.016 | -0.799 | 0.425 |
| HNF1A rs58367757 - T | 16-60 year olds | Asthma | 0.037 | 0.008 | 0.039 | 0.013 | -0.115 | 0.908 |
| HNF1A rs58367757 - T | Over 60 year olds | Recurrent | 0.018 | 0.008 | 0.052 | 0.016 | -1.852 | 0.064 |
| HNF1A rs58367757 - T | Over 60 year olds | Asthma | 0.018 | 0.008 | 0.039 | 0.013 | -1.406 | 0.160 |
| HNF1A rs58367757 - T | Recurrent | Asthma | 0.052 | 0.016 | 0.039 | 0.013 | 0.612 | 0.540 |
| RIN3 rs59985723 - A | General | Under 16 year olds | 0.033 | 0.005 | 0.037 | 0.019 | -0.232 | 0.816 |
| RIN3 rs59985723 - A | General | 16-60 year olds | 0.033 | 0.005 | 0.044 | 0.008 | -1.212 | 0.225 |

| | | | | | | | | |
|---|---|---|---|---|---|---|---|---|
| RIN3 rs59985723 - A | General | Over 60 year olds | 0.033 | 0.005 | 0.023 | 0.008 | 0.996 | 0.319 |
| RIN3 rs59985723 - A | General | Recurrent | 0.033 | 0.005 | 0.021 | 0.017 | 0.651 | 0.515 |
| RIN3 rs59985723 - A | General | Asthma | 0.033 | 0.005 | 0.024 | 0.013 | 0.636 | 0.525 |
| RIN3 rs59985723 - A | Under 16 year olds | 16-60 year olds | 0.037 | 0.019 | 0.044 | 0.008 | -0.338 | 0.735 |
| RIN3 rs59985723 - A | Under 16 year olds | Over 60 year olds | 0.037 | 0.019 | 0.023 | 0.008 | 0.700 | 0.484 |
| RIN3 rs59985723 - A | Under 16 year olds | Recurrent | 0.037 | 0.019 | 0.021 | 0.017 | 0.635 | 0.525 |
| RIN3 rs59985723 - A | Under 16 year olds | Asthma | 0.037 | 0.019 | 0.024 | 0.013 | 0.592 | 0.554 |
| RIN3 rs59985723 - A | 16-60 year olds | Over 60 year olds | 0.044 | 0.008 | 0.023 | 0.008 | 1.891 | 0.059 |
| RIN3 rs59985723 - A | 16-60 year olds | Recurrent | 0.044 | 0.008 | 0.021 | 0.017 | 1.244 | 0.213 |
| RIN3 rs59985723 - A | 16-60 year olds | Asthma | 0.044 | 0.008 | 0.024 | 0.013 | 1.349 | 0.177 |
| RIN3 rs59985723 - A | Over 60 year olds | Recurrent | 0.023 | 0.008 | 0.021 | 0.017 | 0.087 | 0.931 |
| RIN3 rs59985723 - A | Over 60 year olds | Asthma | 0.023 | 0.008 | 0.024 | 0.013 | -0.053 | 0.958 |
| RIN3 rs59985723 - A | Recurrent | Asthma | 0.021 | 0.017 | 0.024 | 0.013 | -0.115 | 0.909 |
| CHRNA5* rs2036527 - A | General | Under 16 year olds | 0.031 | 0.006 | -0.037 | 0.019 | 3.441 | 0.001 |
| CHRNA5* rs2036527 - A | General | 16-60 year olds | 0.031 | 0.006 | 0.030 | 0.008 | 0.189 | 0.850 |
| CHRNA5* rs2036527 - A | General | Over 60 year olds | 0.031 | 0.006 | 0.053 | 0.008 | -2.210 | 0.027 |
| CHRNA5* rs2036527 - A | General | Recurrent | 0.031 | 0.006 | 0.098 | 0.017 | -3.773 | 1.61E-04 |
| CHRNA5* rs2036527 - A | General | Asthma | 0.031 | 0.006 | 0.034 | 0.013 | -0.169 | 0.866 |
| CHRNA5* rs2036527 - A | Under 16 year olds | 16-60 year olds | -0.037 | 0.019 | 0.030 | 0.008 | -3.228 | 0.001 |
| CHRNA5* rs2036527 - A | Under 16 year olds | Over 60 year olds | -0.037 | 0.019 | 0.053 | 0.008 | -4.341 | 1.42E-05 |
| CHRNA5* rs2036527 - A | Under 16 year olds | Recurrent | -0.037 | 0.019 | 0.098 | 0.017 | -5.311 | 1.09E-07 |
| CHRNA5* rs2036527 - A | Under 16 year olds | Asthma | -0.037 | 0.019 | 0.034 | 0.013 | -3.057 | 0.002 |
| CHRNA5* rs2036527 - A | 16-60 year olds | Over 60 year olds | 0.030 | 0.008 | 0.053 | 0.008 | -2.090 | 0.037 |
| CHRNA5* rs2036527 - A | 16-60 year olds | Recurrent | 0.030 | 0.008 | 0.098 | 0.017 | -3.698 | 2.17E-04 |
| CHRNA5* rs2036527 - A | 16-60 year olds | Asthma | 0.030 | 0.008 | 0.034 | 0.013 | -0.275 | 0.783 |
| CHRNA5* rs2036527 - A | Over 60 year olds | Recurrent | 0.053 | 0.008 | 0.098 | 0.017 | -2.385 | 0.017 |
| CHRNA5* rs2036527 - A | Over 60 year olds | Asthma | 0.053 | 0.008 | 0.034 | 0.013 | 1.264 | 0.206 |
| CHRNA5* rs2036527 - A | Recurrent | Asthma | 0.098 | 0.017 | 0.034 | 0.013 | 3.018 | 0.003 |
| APOE rs769449 - A | General | Under 16 year olds | -0.042 | 0.008 | -0.037 | 0.019 | -1.572 | 0.116 |
| APOE rs769449 - A | General | 16-60 year olds | -0.042 | 0.008 | -0.032 | 0.011 | -0.689 | 0.491 |
| APOE rs769449 - A | General | Over 60 year olds | -0.042 | 0.008 | -0.021 | 0.011 | -1.507 | 0.132 |
| APOE rs769449 - A | General | Recurrent | -0.042 | 0.008 | -0.087 | 0.023 | 1.872 | 0.061 |
| APOE rs769449 - A | General | Asthma | -0.042 | 0.008 | -0.019 | 0.018 | -1.151 | 0.250 |
| APOE rs769449 - A | Under 16 year olds | 16-60 year olds | -0.037 | 0.019 | -0.032 | 0.011 | 1.201 | 0.230 |
| APOE rs769449 - A | Under 16 year olds | Over 60 year olds | -0.037 | 0.019 | -0.021 | 0.011 | 0.806 | 0.420 |
| APOE rs769449 - A | Under 16 year olds | Recurrent | -0.037 | 0.019 | -0.087 | 0.023 | 2.525 | 0.012 |

| | | | | | | | | |
|---|---|---|---|---|---|---|---|---|
| APOE rs769449 - A | Under 16 year olds | Asthma | -0.037 | 0.019 | -0.019 | 0.018 | 0.656 | 0.512 |
| APOE rs769449 - A | 16-60 year olds | Over 60 year olds | -0.032 | 0.011 | -0.021 | 0.011 | -0.726 | 0.468 |
| APOE rs769449 - A | 16-60 year olds | Recurrent | -0.032 | 0.011 | -0.087 | 0.023 | 2.141 | 0.032 |
| APOE rs769449 - A | 16-60 year olds | Asthma | -0.032 | 0.011 | -0.019 | 0.018 | -0.641 | 0.522 |
| APOE rs769449 - A | Over 60 year olds | Recurrent | -0.021 | 0.011 | -0.087 | 0.023 | 2.566 | 0.010 |
| APOE rs769449 - A | Over 60 year olds | Asthma | -0.021 | 0.011 | -0.019 | 0.018 | -0.103 | 0.918 |
| APOE rs769449 - A | Recurrent | Asthma | -0.087 | 0.023 | -0.019 | 0.018 | -2.321 | 0.020 |

**Table S5. HLA association analysis results.**

Column headers: chrom: chromosome, pos: position, alt: HLA effect allele, pval: p-value, beta: effect estimate, sebeta: standard error of beta, af_alt: effect allele frequency, af_alt_cases: effect allele frequency among cases, af_alt_controls: effect allele frequency among controls, p_fdr: false discovery rate (FDR)-corrected p-value, OR: odds ratio, OR_l: 95 % confidence interval lower OR; OR_u: 95 % confidence interval upper OR, abbr: abbreviation for pneumonia subgroup. Statistically significant associations are bolded.

| chrom | pos | alt | pval | beta | sebeta | af_alt | af_alt_cases | af_alt_controls | p_fdr | OR | OR_l | OR_u | abbr |
|---|---|---|---|---|---|---|---|---|---|---|---|---|---|
| 6 | 29941260 | A*01:01 | 1.15E-01 | -0.052 | 0.033 | 0.085 | 0.081 | 0.086 | 5.74E-01 | 0.949 | 0.890 | 1.013 | pneu-rec |
| 6 | 32500000 | DRB3*01:01 | 9.42E-01 | 0.002 | 0.023 | 0.182 | 0.183 | 0.182 | 9.89E-01 | 1.002 | 0.957 | 1.049 | pneu-rec |
| **6** | **32500000** | **DRB4*01:03** | **1.71E-04** | **0.082** | **0.022** | **0.217** | **0.227** | **0.217** | **1.13E-02** | **1.085** | **1.040** | **1.132** | **pneu-rec** |
| 6 | 32578770 | DRB1*03:01 | 4.17E-01 | 0.024 | 0.030 | 0.100 | 0.101 | 0.100 | 8.45E-01 | 1.024 | 0.966 | 1.086 | pneu-rec |
| 6 | 32578770 | DRB1*04:03 | 4.70E-01 | 0.281 | 0.389 | 0.003 | 0.003 | 0.003 | 8.82E-01 | 1.324 | 0.618 | 2.838 | pneu-rec |
| 6 | 32578770 | DRB1*04:04 | 1.31E-03 | 0.152 | 0.047 | 0.042 | 0.046 | 0.041 | 6.08E-02 | 1.165 | 1.061 | 1.278 | pneu-rec |
| **6** | **32628180** | **DQA1*03:01** | **7.35E-05** | **0.116** | **0.029** | **0.110** | **0.118** | **0.110** | **7.47E-03** | **1.123** | **1.060** | **1.189** | **pneu-rec** |
| 6 | 32628180 | DQA1*05:01 | 4.99E-01 | 0.020 | 0.030 | 0.106 | 0.106 | 0.106 | 8.97E-01 | 1.020 | 0.963 | 1.081 | pneu-rec |
| 6 | 32659470 | DQB1*02:01 | 4.49E-01 | 0.023 | 0.030 | 0.100 | 0.101 | 0.100 | 8.68E-01 | 1.023 | 0.965 | 1.084 | pneu-rec |
| **6** | **32659470** | **DQB1*03:02** | **1.04E-04** | **0.105** | **0.027** | **0.118** | **0.126** | **0.118** | **8.93E-03** | **1.111** | **1.053** | **1.171** | **pneu-rec** |
| 6 | 29941260 | A*01:01 | 2.62E-02 | -0.061 | 0.028 | 0.084 | 0.080 | 0.086 | 3.54E-01 | 0.941 | 0.891 | 0.993 | pneu-asthma |
| 6 | 32500000 | DRB3*01:01 | 2.28E-01 | -0.024 | 0.020 | 0.180 | 0.177 | 0.181 | 7.04E-01 | 0.977 | 0.940 | 1.015 | pneu-asthma |
| 6 | 32500000 | DRB4*01:03 | 7.61E-01 | 0.005 | 0.018 | 0.231 | 0.232 | 0.230 | 9.57E-01 | 1.005 | 0.971 | 1.042 | pneu-asthma |
| 6 | 32578770 | DRB1*03:01 | 1.37E-01 | -0.037 | 0.025 | 0.101 | 0.098 | 0.102 | 5.98E-01 | 0.964 | 0.918 | 1.012 | pneu-asthma |
| 6 | 32578770 | DRB1*04:03 | 8.94E-01 | -0.040 | 0.303 | 0.003 | 0.003 | 0.003 | 9.72E-01 | 0.960 | 0.530 | 1.741 | pneu-asthma |
| 6 | 32578770 | DRB1*04:04 | 1.52E-01 | 0.055 | 0.039 | 0.046 | 0.048 | 0.045 | 6.21E-01 | 1.057 | 0.980 | 1.140 | pneu-asthma |
| 6 | 32628180 | DQA1*03:01 | 1.04E-01 | 0.039 | 0.024 | 0.120 | 0.123 | 0.118 | 5.74E-01 | 1.040 | 0.992 | 1.089 | pneu-asthma |
| 6 | 32628180 | DQA1*05:01 | 1.59E-01 | -0.035 | 0.025 | 0.106 | 0.104 | 0.108 | 6.22E-01 | 0.966 | 0.920 | 1.014 | pneu-asthma |
| 6 | 32659470 | DQB1*02:01 | 1.56E-01 | -0.035 | 0.025 | 0.101 | 0.098 | 0.102 | 6.21E-01 | 0.966 | 0.920 | 1.013 | pneu-asthma |
| 6 | 32659470 | DQB1*03:02 | 1.02E-01 | 0.036 | 0.022 | 0.128 | 0.132 | 0.127 | 5.74E-01 | 1.037 | 0.993 | 1.083 | pneu-asthma |
| **6** | **29941260** | **A*01:01** | **1.25E-04** | **-0.070** | **0.018** | **0.085** | **0.079** | **0.086** | **9.72E-03** | **0.932** | **0.900** | **0.966** | **pneu-16-60** |
| **6** | **32500000** | **DRB3*01:01** | **5.44E-06** | **-0.058** | **0.013** | **0.181** | **0.173** | **0.182** | **1.22E-03** | **0.943** | **0.920** | **0.967** | **pneu-16-60** |
| **6** | **32500000** | **DRB4*01:03** | **1.37E-05** | **0.051** | **0.012** | **0.217** | **0.226** | **0.217** | **2.18E-03** | **1.053** | **1.029** | **1.077** | **pneu-16-60** |
| **6** | **32578770** | **DRB1*03:01** | **4.01E-04** | **-0.058** | **0.016** | **0.100** | **0.094** | **0.100** | **2.49E-02** | **0.944** | **0.914** | **0.975** | **pneu-16-60** |
| 6 | 32578770 | DRB1*04:03 | 1.83E-01 | 0.278 | 0.209 | 0.003 | 0.003 | 0.003 | 6.52E-01 | 1.320 | 0.877 | 1.988 | pneu-16-60 |
| **6** | **32578770** | **DRB1*04:04** | **8.54E-07** | **0.126** | **0.026** | **0.042** | **0.046** | **0.041** | **4.78E-04** | **1.134** | **1.079** | **1.192** | **pneu-16-60** |
| **6** | **32628180** | **DQA1*03:01** | **3.19E-05** | **0.066** | **0.016** | **0.110** | **0.117** | **0.110** | **3.97E-03** | **1.068** | **1.035** | **1.102** | **pneu-16-60** |
| **6** | **32628180** | **DQA1*05:01** | **4.88E-04** | **-0.057** | **0.016** | **0.105** | **0.100** | **0.106** | **2.73E-02** | **0.945** | **0.915** | **0.975** | **pneu-16-60** |
| **6** | **32659470** | **DQB1*02:01** | **4.42E-04** | **-0.057** | **0.016** | **0.100** | **0.094** | **0.100** | **2.60E-02** | **0.944** | **0.914** | **0.975** | **pneu-16-60** |
| **6** | **32659470** | **DQB1*03:02** | **3.81E-05** | **0.060** | **0.015** | **0.119** | **0.126** | **0.118** | **4.26E-03** | **1.062** | **1.032** | **1.093** | **pneu-16-60** |
| 6 | 29941260 | A*01:01 | 4.58E-01 | -0.012 | 0.016 | 0.085 | 0.085 | 0.086 | 8.73E-01 | 0.988 | 0.958 | 1.020 | pneu-o60 |
| 6 | 32500000 | DRB3*01:01 | 7.47E-01 | 0.004 | 0.011 | 0.182 | 0.183 | 0.182 | 9.56E-01 | 1.004 | 0.982 | 1.026 | pneu-o60 |
| 6 | 32500000 | DRB4*01:03 | 1.79E-02 | 0.025 | 0.011 | 0.217 | 0.216 | 0.217 | 2.78E-01 | 1.026 | 1.004 | 1.048 | pneu-o60 |

| | | | | | | | | | | | | | |
|---|---|---|---|---|---|---|---|---|---|---|---|---|---|
| 6 | 32578770 | DRB1*03:01 | 6.06E-01 | 0.007 | 0.014 | 0.100 | 0.100 | 0.100 | 9.14E-01 | 1.007 | 0.979 | 1.036 | pneu-o60 |
| 6 | 32578770 | DRB1*04:03 | 2.50E-01 | -0.222 | 0.193 | 0.003 | 0.003 | 0.003 | 7.23E-01 | 0.801 | 0.549 | 1.169 | pneu-o60 |
| 6 | 32578770 | DRB1*04:04 | 5.30E-02 | 0.047 | 0.024 | 0.041 | 0.042 | 0.041 | 4.44E-01 | 1.048 | 0.999 | 1.098 | pneu-o60 |
| 6 | 32628180 | DQA1*03:01 | 3.46E-02 | 0.031 | 0.015 | 0.110 | 0.108 | 0.110 | 3.87E-01 | 1.031 | 1.002 | 1.061 | pneu-o60 |
| 6 | 32628180 | DQA1*05:01 | 6.65E-01 | 0.006 | 0.014 | 0.106 | 0.106 | 0.106 | 9.33E-01 | 1.006 | 0.978 | 1.035 | pneu-o60 |
| 6 | 32659470 | DQB1*02:01 | 6.36E-01 | 0.007 | 0.014 | 0.100 | 0.100 | 0.100 | 9.22E-01 | 1.007 | 0.979 | 1.036 | pneu-o60 |
| 6 | 32659470 | DQB1*03:02 | 5.74E-02 | 0.026 | 0.014 | 0.118 | 0.116 | 0.118 | 4.49E-01 | 1.026 | 0.999 | 1.054 | pneu-o60 |
| 6 | 29941260 | A*01:01 | 1.85E-03 | -0.036 | 0.012 | 0.085 | 0.083 | 0.086 | 7.41E-02 | 0.964 | 0.942 | 0.987 | pneu-all |
| 6 | 32500000 | DRB3*01:01 | 7.57E-03 | -0.022 | 0.008 | 0.181 | 0.179 | 0.182 | 1.84E-01 | 0.978 | 0.962 | 0.994 | pneu-all |
| **6** | **32500000** | **DRB4*01:03** | **2.22E-06** | **0.037** | **0.008** | **0.218** | **0.221** | **0.217** | **8.29E-04** | **1.037** | **1.022** | **1.053** | **pneu-all** |
| 6 | 32578770 | DRB1*03:01 | 4.32E-02 | -0.021 | 0.011 | 0.100 | 0.098 | 0.100 | 4.31E-01 | 0.979 | 0.959 | 0.999 | pneu-all |
| 6 | 32578770 | DRB1*04:03 | 3.52E-01 | 0.128 | 0.138 | 0.003 | 0.003 | 0.003 | 8.14E-01 | 1.137 | 0.868 | 1.490 | pneu-all |
| **6** | **32578770** | **DRB1*04:04** | **4.07E-07** | **0.087** | **0.017** | **0.042** | **0.044** | **0.041** | **4.55E-04** | **1.090** | **1.055** | **1.128** | **pneu-all** |
| **6** | **32628180** | **DQA1*03:01** | **3.46E-06** | **0.049** | **0.010** | **0.110** | **0.113** | **0.110** | **9.66E-04** | **1.050** | **1.028** | **1.072** | **pneu-all** |
| 6 | 32628180 | DQA1*05:01 | 4.55E-02 | -0.021 | 0.010 | 0.105 | 0.104 | 0.106 | 4.40E-01 | 0.979 | 0.959 | 1.000 | pneu-all |
| 6 | 32659470 | DQB1*02:01 | 4.59E-02 | -0.021 | 0.011 | 0.100 | 0.098 | 0.100 | 4.40E-01 | 0.979 | 0.959 | 1.000 | pneu-all |
| **6** | **32659470** | **DQB1*03:02** | **8.77E-06** | **0.043** | **0.010** | **0.119** | **0.121** | **0.118** | **1.63E-03** | **1.044** | **1.024** | **1.064** | **pneu-all** |
| 6 | 29941260 | A*01:01 | 5.42E-01 | 0.029 | 0.048 | 0.086 | 0.088 | 0.086 | 9.00E-01 | 1.030 | 0.937 | 1.132 | pneu-u16 |
| 6 | 32500000 | DRB3*01:01 | 4.46E-01 | -0.026 | 0.034 | 0.182 | 0.178 | 0.182 | 8.68E-01 | 0.974 | 0.910 | 1.042 | pneu-u16 |
| **6** | **32500000** | **DRB4*01:03** | **2.89E-05** | **0.131** | **0.031** | **0.217** | **0.241** | **0.217** | **3.97E-03** | **1.140** | **1.072** | **1.213** | **pneu-u16** |
| 6 | 32578770 | DRB1*03:01 | 9.64E-01 | 0.002 | 0.043 | 0.100 | 0.102 | 0.100 | 9.91E-01 | 1.002 | 0.920 | 1.091 | pneu-u16 |
| **6** | **32578770** | **DRB1*04:03** | **8.60E-05** | **1.699** | **0.433** | **0.003** | **0.004** | **0.003** | **8.02E-03** | **5.470** | **2.342** | **12.774** | **pneu-u16** |
| 6 | 32578770 | DRB1*04:04 | 3.76E-02 | 0.147 | 0.071 | 0.042 | 0.048 | 0.041 | 4.00E-01 | 1.158 | 1.008 | 1.330 | pneu-u16 |
| **6** | **32628180** | **DQA1*03:01** | **1.30E-04** | **0.160** | **0.042** | **0.110** | **0.128** | **0.110** | **9.72E-03** | **1.173** | **1.081** | **1.273** | **pneu-u16** |
| 6 | 32628180 | DQA1*05:01 | 8.67E-01 | 0.007 | 0.043 | 0.106 | 0.107 | 0.106 | 9.67E-01 | 1.007 | 0.925 | 1.097 | pneu-u16 |
| 6 | 32659470 | DQB1*02:01 | 8.81E-01 | 0.007 | 0.043 | 0.100 | 0.102 | 0.100 | 9.68E-01 | 1.007 | 0.924 | 1.096 | pneu-u16 |
| **6** | **32659470** | **DQB1*03:02** | **1.64E-04** | **0.146** | **0.039** | **0.118** | **0.138** | **0.118** | **1.13E-02** | **1.157** | **1.073** | **1.248** | **pneu-u16** |

**Table S6. Pneumonia subgroup associations with APOE full genotypes.** FinnGen Alzheimer phenotype (G6_ALZHEIMER) is included in the results for comparison. Group, tested subgroup; P, p-value; OR, odd's ratio; CI95lower, lower 95 % confidence interval of OR; CI95upper, upper 95 % confidence interval of OR; N, number of cases; apoe_variable, APOE full genotype.

| group | P | OR | CI95lower | CI95upper | N | apoe_variable |
|---|---|---|---|---|---|---|
| PNEUMONIA_CUSTOM_ALL | 8.43E-02 | 1.19 | 0.98 | 1.45 | 141014 | e2e2_vs_e3e3 |
| PNEUMONIA_UNDER_16 | 6.89E-01 | 1.20 | 0.49 | 2.97 | 120511 | e2e2_vs_e3e3 |
| PNEUMONIA_16_TO_60 | 2.48E-01 | 1.20 | 0.88 | 1.63 | 128007 | e2e2_vs_e3e3 |
| PNEUMONIA_60_AND_OVER | 2.05E-01 | 1.19 | 0.91 | 1.56 | 131174 | e2e2_vs_e3e3 |
| PNEUMO_3_OR_MORE | 3.08E-01 | 1.31 | 0.78 | 2.22 | 121600 | e2e2_vs_e3e3 |
| ASTHMA_CUSTOM | 3.53E-01 | 0.89 | 0.70 | 1.14 | 141014 | e2e2_vs_e3e3 |
| PNEUMO_ASTHMA_CUSTOM | 1.55E-01 | 1.44 | 0.87 | 2.38 | 17545 | e2e2_vs_e3e3 |
| G6_ALZHEIMER | 6.78E-01 | 0.88 | 0.47 | 1.64 | 141014 | e2e2_vs_e3e3 |
| PNEUMONIA_CUSTOM_ALL | 1.81E-01 | 1.03 | 0.99 | 1.07 | 160100 | e2e3_vs_e3e3 |
| PNEUMONIA_UNDER_16 | 3.29E-01 | 1.09 | 0.92 | 1.28 | 136744 | e2e3_vs_e3e3 |
| PNEUMONIA_16_TO_60 | 4.57E-01 | 1.02 | 0.96 | 1.09 | 145240 | e2e3_vs_e3e3 |
| PNEUMONIA_60_AND_OVER | 9.88E-01 | 1.00 | 0.94 | 1.06 | 148928 | e2e3_vs_e3e3 |
| PNEUMO_3_OR_MORE | 4.92E-02 | 1.12 | 1.00 | 1.26 | 138013 | e2e3_vs_e3e3 |
| ASTHMA_CUSTOM | 2.67E-01 | 0.97 | 0.93 | 1.02 | 160100 | e2e3_vs_e3e3 |
| PNEUMO_ASTHMA_CUSTOM | 9.69E-02 | 1.09 | 0.99 | 1.20 | 19844 | e2e3_vs_e3e3 |
| G6_ALZHEIMER | 5.31E-08 | 0.67 | 0.58 | 0.77 | 160100 | e2e3_vs_e3e3 |
| PNEUMONIA_CUSTOM_ALL | 8.36E-02 | 0.93 | 0.85 | 1.01 | 145109 | e2e4_vs_e3e3 |
| PNEUMONIA_UNDER_16 | 8.32E-01 | 1.04 | 0.75 | 1.43 | 124079 | e2e4_vs_e3e3 |
| PNEUMONIA_16_TO_60 | 1.47E-01 | 0.91 | 0.80 | 1.03 | 131760 | e2e4_vs_e3e3 |
| PNEUMONIA_60_AND_OVER | 2.65E-01 | 0.94 | 0.83 | 1.05 | 135014 | e2e4_vs_e3e3 |
| PNEUMO_3_OR_MORE | 9.87E-01 | 1.00 | 0.79 | 1.26 | 125195 | e2e4_vs_e3e3 |
| ASTHMA_CUSTOM | 1.06E-01 | 0.93 | 0.85 | 1.02 | 145109 | e2e4_vs_e3e3 |
| PNEUMO_ASTHMA_CUSTOM | 5.60E-01 | 0.94 | 0.77 | 1.15 | 18019 | e2e4_vs_e3e3 |
| G6_ALZHEIMER | 2.30E-15 | 2.15 | 1.78 | 2.60 | 145109 | e2e4_vs_e3e3 |
| PNEUMONIA_CUSTOM_ALL | 5.24E-05 | 0.95 | 0.92 | 0.97 | 207816 | e3e4_vs_e3e3 |
| PNEUMONIA_UNDER_16 | 9.33E-01 | 1.00 | 0.91 | 1.11 | 178257 | e3e4_vs_e3e3 |
| PNEUMONIA_16_TO_60 | 3.61E-03 | 0.94 | 0.91 | 0.98 | 189121 | e3e4_vs_e3e3 |
| PNEUMONIA_60_AND_OVER | 2.34E-01 | 0.98 | 0.94 | 1.01 | 193452 | e3e4_vs_e3e3 |
| PNEUMO_3_OR_MORE | 4.51E-02 | 0.92 | 0.86 | 1.00 | 179723 | e3e4_vs_e3e3 |
| ASTHMA_CUSTOM | 8.04E-06 | 0.94 | 0.91 | 0.96 | 207816 | e3e4_vs_e3e3 |
| PNEUMO_ASTHMA_CUSTOM | 4.56E-03 | 0.91 | 0.86 | 0.97 | 25394 | e3e4_vs_e3e3 |
| G6_ALZHEIMER | 2.14E-203 | 2.75 | 2.58 | 2.94 | 207816 | e3e4_vs_e3e3 |
| PNEUMONIA_CUSTOM_ALL | 5.26E-02 | 0.94 | 0.88 | 1.00 | 148182 | e4e4_vs_e3e3 |
| PNEUMONIA_UNDER_16 | 4.12E-01 | 1.10 | 0.87 | 1.39 | 126780 | e4e4_vs_e3e3 |
| PNEUMONIA_16_TO_60 | 1.25E-01 | 0.93 | 0.84 | 1.02 | 134622 | e4e4_vs_e3e3 |
| PNEUMONIA_60_AND_OVER | 8.33E-01 | 1.01 | 0.92 | 1.11 | 137848 | e4e4_vs_e3e3 |
| PNEUMO_3_OR_MORE | 1.43E-01 | 0.86 | 0.70 | 1.05 | 127881 | e4e4_vs_e3e3 |
| ASTHMA_CUSTOM | 1.57E-03 | 0.89 | 0.83 | 0.96 | 148182 | e4e4_vs_e3e3 |
| PNEUMO_ASTHMA_CUSTOM | 9.69E-01 | 1.00 | 0.85 | 1.17 | 18348 | e4e4_vs_e3e3 |
| G6_ALZHEIMER | 0.00E+00 | 9.13 | 8.16 | 10.23 | 148182 | e4e4_vs_e3e3 |

**Table S7. Pneumonia subgroup associations with APOE allele counts.** FinnGen Alzheimer phenotype (G6_ALZHEIMER) is included in the results for comparison. Group, tested subgroup; P, p-value; OR, odd's ratio; CI95lower, lower 95 % confidence interval of OR; CI95upper, upper 95 % confidence interval of OR; N, number of cases; apoe_variable, APOE allele count.

| group | P | OR | CI95lower | CI95upper | N | apoe_variable |
|---|---|---|---|---|---|---|
| PNEUMONIA_CUSTOM_ALL | 3.94E-02 | 1.04 | 1.00 | 1.07 | 240824 | e2_count |
| PNEUMONIA_UNDER_16 | 3.44E-01 | 1.07 | 0.93 | 1.23 | 206500 | e2_count |
| PNEUMONIA_16_TO_60 | 2.78E-01 | 1.03 | 0.98 | 1.09 | 219056 | e2_count |
| PNEUMONIA_60_AND_OVER | 7.79E-01 | 1.01 | 0.96 | 1.06 | 224178 | e2_count |
| PNEUMO_3_OR_MORE | 1.19E-02 | 1.13 | 1.03 | 1.25 | 208225 | e2_count |
| ASTHMA_CUSTOM | 4.97E-01 | 0.99 | 0.95 | 1.03 | 240824 | e2_count |
| PNEUMO_ASTHMA_CUSTOM | 3.17E-02 | 1.10 | 1.01 | 1.19 | 29263 | e2_count |
| G6_ALZHEIMER | 2.59E-24 | 0.57 | 0.51 | 0.64 | 240824 | e2_count |
| PNEUMONIA_CUSTOM_ALL | 1.20E-03 | 1.03 | 1.01 | 1.05 | 240824 | e3_count |
| PNEUMONIA_UNDER_16 | 4.16E-01 | 0.97 | 0.90 | 1.04 | 206500 | e3_count |
| PNEUMONIA_16_TO_60 | 9.47E-03 | 1.04 | 1.01 | 1.07 | 219056 | e3_count |
| PNEUMONIA_60_AND_OVER | 4.23E-01 | 1.01 | 0.98 | 1.04 | 224178 | e3_count |
| PNEUMO_3_OR_MORE | 2.47E-01 | 1.03 | 0.98 | 1.09 | 208225 | e3_count |
| ASTHMA_CUSTOM | 5.02E-07 | 1.05 | 1.03 | 1.08 | 240824 | e3_count |
| PNEUMO_ASTHMA_CUSTOM | 1.90E-01 | 1.03 | 0.99 | 1.08 | 29263 | e3_count |
| G6_ALZHEIMER | 1.33E-280 | 0.44 | 0.42 | 0.46 | 240824 | e3_count |
| PNEUMONIA_CUSTOM_ALL | 1.62E-06 | 0.95 | 0.93 | 0.97 | 240824 | e4_count |
| PNEUMONIA_UNDER_16 | 7.19E-01 | 1.01 | 0.94 | 1.10 | 206500 | e4_count |
| PNEUMONIA_16_TO_60 | 5.11E-04 | 0.95 | 0.92 | 0.98 | 219056 | e4_count |
| PNEUMONIA_60_AND_OVER | 2.86E-01 | 0.98 | 0.95 | 1.01 | 224178 | e4_count |
| PNEUMO_3_OR_MORE | 5.32E-03 | 0.92 | 0.86 | 0.97 | 208225 | e4_count |
| ASTHMA_CUSTOM | 2.86E-07 | 0.94 | 0.92 | 0.96 | 240824 | e4_count |
| PNEUMO_ASTHMA_CUSTOM | 6.88E-03 | 0.93 | 0.89 | 0.98 | 29263 | e4_count |
| G6_ALZHEIMER | 0.00E+00 | 3.00 | 2.87 | 3.15 | 240824 | e4_count |

**Table S8. Colocalisation results (PP4 > 0.8) of pneumonia and Genotype-Tissue Expression Project (GTEx) tissues.**

Column header abbreviations: PP0: probability that neither trait has a genetic association in the region, PP1: probability that only trait 1 has a genetic association in the region, PP2: probability that only trait 2 has a genetic association in the region, PP3: probability that both traits are associated, but with different causal variants, PP4: probability that both traits are associated and share a single causal variant.

| tissue | gene | gene_name | PP0 | PP1 | PP2 | PP3 | PP4 |
|---|---|---|---|---|---|---|---|
| Cells_EBV-transformed_lymphocytes | ENSG00000196126.11 | HLA-DRB1 | 1.42E-13 | 0.0000734 | 7.44E-11 | 0.038 | 0.962 |
| Cells_EBV-transformed_lymphocytes | ENSG00000255158.1 | RP11-754B17.1 | 0.02 | 0.003 | 0.024 | 0.003 | 0.95 |
| Whole_Blood | ENSG00000158716.8 | DUSP23 | 0.004 | 0.004 | 0.027 | 0.023 | 0.942 |
| Lung | ENSG00000080644.15 | CHRNA3 | 0.000268 | 0.000574 | 0.021 | 0.045 | 0.933 |
| Whole_Blood | ENSG00000168477.17 | TNXB | 0.002 | 0.000996 | 0.043 | 0.024 | 0.931 |
| Lung | ENSG00000226167.1 | AP4B1-AS1 | 0.000442 | 0.002 | 0.022 | 0.084 | 0.891 |
| Cells_EBV-transformed_lymphocytes | ENSG00000224389.8 | C4B | 0.000155 | 0.00011 | 0.104 | 0.073 | 0.822 |

**Table S9. Results table of traits showing significant genetic correlation ($p < 6.48 * 10^{-5}$) with pneumonia using linkage disequilibrium score regression (LDSC) method.**

Column header abbreviations: GWAS ID: ID of the trait in IEU Open GWAS Project website (https://gwas.mrcieu.ac.uk/); $r_g$: genetic correlation between pneumonia and trait; se: standard error; p: p-value of the correlation

| GWAS_ID | Trait | Group | $r_g$ | se | p |
|---|---|---|---|---|---|
| ukb-b-6306 | Overall health rating | activity_fitness_sleep | 0.4912 | 3.74E-02 | 2.12E-39 |
| ukb-b-18335 | Wheeze or whistling in the chest in last year | lung_function | 0.4957 | 3.80E-02 | 8.22E-39 |
| ukb-a-225 | Smoking status: Current | smoking | 0.4843 | 4.22E-02 | 1.64E-30 |
| ukb-b-223 | Current tobacco smoking | smoking | 0.4598 | 4.05E-02 | 6.98E-30 |
| ukb-b-929 | Frequency of tiredness / lethargy in last 2 weeks | mood_behaviour | 0.4098 | 3.73E-02 | 4.74E-28 |
| ukb-b-18096 | Leg fat mass (right) | anthropometry | 0.3153 | 2.93E-02 | 4.83E-27 |
| ukb-b-20292 | Taking other prescription medications | drugs_supplements | 0.5027 | 4.68E-02 | 6.43E-27 |
| ukb-b-13764 | Long-standing illness, disability or infirmity | diseases | 0.4672 | 4.36E-02 | 8.05E-27 |
| ukb-b-7212 | Leg fat mass (left) | anthropometry | 0.3127 | 2.92E-02 | 8.10E-27 |
| ukb-b-8338 | Arm fat mass (left) | anthropometry | 0.3111 | 2.91E-02 | 1.08E-26 |
| ukb-b-6704 | Arm fat mass (right) | anthropometry | 0.3119 | 2.92E-02 | 1.28E-26 |
| ukb-b-9405 | Waist circumference | anthropometry | 0.3209 | 3.01E-02 | 1.36E-26 |
| ukb-b-20531 | Leg fat percentage (right) | anthropometry | 0.3184 | 3.00E-02 | 2.28E-26 |
| ukb-b-19953 | Body mass index (BMI) | anthropometry | 0.3172 | 2.99E-02 | 2.96E-26 |
| ukb-b-18377 | Leg fat percentage (left) | anthropometry | 0.3164 | 2.99E-02 | 3.68E-26 |
| ukb-b-19393 | Whole body fat mass | anthropometry | 0.2984 | 2.83E-02 | 6.11E-26 |
| ukb-b-12854 | Arm fat percentage (right) | anthropometry | 0.3099 | 2.97E-02 | 1.80E-25 |
| ieu-a-1239 | Years of schooling | education_intelligence | -0.31 | 2.98E-02 | 2.57E-25 |
| ukb-b-20188 | Arm fat percentage (left) | anthropometry | 0.3057 | 2.95E-02 | 3.03E-25 |
| ukb-b-8909 | Body fat percentage | anthropometry | 0.2976 | 2.90E-02 | 9.26E-25 |
| ieu-b-4877 | smoking initiation | smoking | 0.3663 | 3.60E-02 | 2.45E-24 |
| ukb-b-20044 | Trunk fat mass | anthropometry | 0.2816 | 2.79E-02 | 5.54E-24 |
| ukb-b-12405 | Age at first live birth | gynecology_fertility | -0.3917 | 3.90E-02 | 8.90E-24 |
| ukb-b-11085 | Medication for pain relief, constipation, heartburn: None of the above | drugs_supplements | -0.3696 | 3.71E-02 | 2.35E-23 |
| ukb-b-4711 | Usual walking pace | activity_fitness_sleep | -0.3621 | 3.66E-02 | 4.31E-23 |
| ukb-b-9130 | Pain type(s) experienced in last month: None of the above | pain_skeletal | -0.3866 | 3.97E-02 | 1.99E-22 |
| ukb-b-16407 | Trunk fat percentage | anthropometry | 0.2802 | 2.89E-02 | 3.40E-22 |
| ukb-b-16489 | Qualifications: College or University degree | education_intelligence | -0.3155 | 3.28E-02 | 6.14E-22 |
| ukb-b-3656 | Number of treatments/medications taken | drugs_supplements | 0.4054 | 4.22E-02 | 8.06E-22 |
| ukb-d-20116_0 | Smoking status: Never | smoking | -0.3587 | 3.78E-02 | 2.28E-21 |

| GWAS_ID | Trait | Group | $r_g$ | se | p |
|---|---|---|---|---|---|
| ukb-b-2535 | Falls in the last year | activity_fitness_sleep | 0.3944 | 4.22E-02 | 8.80E-21 |
| ukb-b-11842 | Weight | anthropometry | 0.2528 | 2.72E-02 | 1.41E-20 |
| ukb-b-10591 | Chest pain or discomfort | pain_skeletal | 0.419 | 4.55E-02 | 3.47E-20 |
| ukb-b-9667 | Illness, injury, bereavement, stress in last 2 years: Financial difficulties | work_stress | 0.413 | 4.53E-02 | 7.77E-20 |
| ukb-b-2053 | Health satisfaction | activity_fitness_sleep | 0.4442 | 4.94E-02 | 2.34E-19 |
| ukb-b-6134 | Age completed full time education | education_intelligence | -0.3743 | 4.18E-02 | 3.27E-19 |
| ukb-b-15590 | Hip circumference | anthropometry | 0.2532 | 2.83E-02 | 3.65E-19 |
| ukb-b-17729 | Qualifications: None of the above | education_intelligence | 0.3216 | 3.60E-02 | 4.15E-19 |
| ukb-b-15869 | Types of physical activity in last 4 weeks: None of the above | activity_fitness_sleep | 0.4506 | 5.07E-02 | 5.72E-19 |
| ukb-b-9543 | Shortness of breath walking on level ground | activity_fitness_sleep | 0.5205 | 5.87E-02 | 7.01E-19 |
| ukb-b-16207 | Blood clot, DVT, bronchitis, emphysema, asthma, rhinitis, eczema, allergy diagnosed by doctor: Emphysema/chronic bronchitis | diseases | 0.6262 | 7.12E-02 | 1.53E-18 |
| ukb-b-12930 | Mouth/teeth dental problems: Dentures | dental | 0.349 | 4.12E-02 | 2.28E-17 |
| ieu-b-25 | Cigarettes per Day | smoking | 0.3553 | 4.22E-02 | 3.58E-17 |
| ieu-b-142 | Cigarettes smoked per day | smoking | 0.3553 | 4.22E-02 | 3.59E-17 |
| ukb-b-10831 | Pack years of smoking | smoking | 0.4328 | 5.14E-02 | 3.70E-17 |
| ukb-b-17685 | Maternal smoking around birth | smoking | 0.3411 | 4.06E-02 | 4.68E-17 |
| ukb-b-16254 | Pain type(s) experienced in last month: Knee pain | pain_skeletal | 0.3533 | 4.25E-02 | 8.56E-17 |
| ukb-b-11615 | Qualifications: A levels/AS levels or equivalent | education_intelligence | -0.3112 | 3.74E-02 | 9.26E-17 |
| ukb-a-581 | Diagnoses - main ICD10: R07 Pain in throat and chest | diseases | 0.5233 | 6.44E-02 | 4.26E-16 |
| ukb-b-2134 | Past tobacco smoking | smoking | -0.2954 | 3.72E-02 | 1.82E-15 |
| ukb-b-4733 | Number of operations, self-reported | diseases | 0.3889 | 4.92E-02 | 2.62E-15 |
| ukb-b-7289 | Pain type(s) experienced in last month: Hip pain | pain_skeletal | 0.4118 | 5.22E-02 | 3.22E-15 |
| ukb-b-1419 | Frequency of unenthusiasm / disinterest in last 2 weeks | mood_behaviour | 0.3551 | 4.58E-02 | 9.16E-15 |
| ukb-b-2002 | Job involves heavy manual or physical work | activity_fitness_sleep | 0.3112 | 4.04E-02 | 1.40E-14 |
| ukb-b-7337 | Types of physical activity in last 4 weeks: Walking for pleasure (not as a means of transport) | activity_fitness_sleep | -0.3463 | 4.54E-02 | 2.43E-14 |
| ukb-b-6991 | Seen doctor (GP) for nerves, anxiety, tension or depression | mood_behaviour | 0.3054 | 4.08E-02 | 7.42E-14 |
| ukb-b-12687 | Mother's age at death | family_memb_health | -0.4965 | 6.74E-02 | 1.82E-13 |
| ukb-b-17595 | Medication for pain relief, constipation, heartburn: Paracetamol | drugs_supplements | 0.313 | 4.30E-02 | 3.52E-13 |
| ukb-b-11411 | Blood clot, DVT, bronchitis, emphysema, asthma, rhinitis, eczema, allergy diagnosed by doctor: Blood clot in the leg (DVT) | diseases | 0.5015 | 6.91E-02 | 3.86E-13 |

| GWAS_ID | Trait | Group | $r_g$ | se | p |
|---|---|---|---|---|---|
| ukb-b-17601 | Mouth/teeth dental problems: None of the above | dental | -0.3201 | 4.42E-02 | 4.33E-13 |
| ukb-b-18596 | Pain type(s) experienced in last month: Neck or shoulder pain | pain_skeletal | 0.3314 | 4.63E-02 | 8.11E-13 |
| ukb-b-8764 | Types of physical activity in last 4 weeks: Other exercises (eg: swimming, cycling, keep fit, bowling) | activity_fitness_sleep | -0.3062 | 4.28E-02 | 8.53E-13 |
| ukb-b-9838 | Pain type(s) experienced in last month: Back pain | pain_skeletal | 0.3384 | 4.73E-02 | 8.54E-13 |
| ukb-b-20261 | Ever smoked | smoking | 0.2557 | 3.62E-02 | 1.51E-12 |
| ukb-b-16099 | Leg fat-free mass (left) | anthropometry | 0.1898 | 2.72E-02 | 3.10E-12 |
| ukb-b-16446 | Basal metabolic rate | anthropometry | 0.1826 | 2.63E-02 | 3.80E-12 |
| ukb-b-5664 | Frequency of tenseness / restlessness in last 2 weeks | mood_behaviour | 0.3097 | 4.46E-02 | 3.99E-12 |
| ukb-b-8727 | Age at last live birth | gynecology_fertility | -0.3114 | 4.52E-02 | 5.68E-12 |
| ukb-b-10387 | Leg pain on walking | pain_skeletal | 0.4129 | 6.05E-02 | 8.70E-12 |
| ukb-b-2830 | Financial situation satisfaction | work_stress | 0.353 | 5.21E-02 | 1.28E-11 |
| ukb-b-19925 | Arm fat-free mass (left) | anthropometry | 0.1809 | 2.67E-02 | 1.37E-11 |
| ebi-a-GCST006702 | Parental longevity (combined parental age at death) | family_memb_health | -0.4615 | 6.92E-02 | 2.51E-11 |
| ebi-a-GCST006250 | Intelligence | education_intelligence | -0.1996 | 3.01E-02 | 3.37E-11 |
| ukb-b-11303 | Father's age at death | family_memb_health | -0.3809 | 5.87E-02 | 8.73E-11 |
| ukb-a-227 | Alcohol drinker status: Previous | alcohol | 0.4305 | 6.66E-02 | 9.93E-11 |
| met-d-LA_pct | Ratio of linoleic acid to total fatty acids | NMR_metabo_UKBB | -0.3376 | 5.24E-02 | 1.16E-10 |
| ukb-b-14961 | Other serious medical condition/disability diagnosed by doctor | diseases | 0.3364 | 5.23E-02 | 1.25E-10 |
| ukb-b-12828 | Leg fat-free mass (right) | anthropometry | 0.1722 | 2.69E-02 | 1.59E-10 |
| ukb-b-18674 | Ever unenthusiastic/disinterested for a whole week | mood_behaviour | 0.3849 | 6.12E-02 | 3.28E-10 |
| ukb-b-14926 | Types of transport used (excluding work): Walk | activity_fitness_sleep | -0.3032 | 4.85E-02 | 3.91E-10 |
| ukb-b-12654 | Former alcohol drinker | alcohol | 0.5117 | 8.22E-02 | 4.85E-10 |
| ebi-a-GCST006867 | Type 2 diabetes | diseases | 0.2666 | 4.29E-02 | 4.95E-10 |
| ukb-b-17687 | Illness, injury, bereavement, stress in last 2 years: None of the above | work_stress | -0.3231 | 5.23E-02 | 6.41E-10 |
| ukb-b-9433 | Age started oral contraceptive pill | gynecology_fertility | -0.3465 | 5.61E-02 | 6.49E-10 |
| ukb-b-7953 | Forced vital capacity (FVC) | lung_function | -0.1866 | 3.04E-02 | 8.87E-10 |
| ukb-b-19520 | Arm fat-free mass (right) | anthropometry | 0.1602 | 2.62E-02 | 9.74E-10 |
| ukb-b-14713 | Forced vital capacity (FVC), Best measure | lung_function | -0.1882 | 3.10E-02 | 1.23E-09 |
| ukb-b-18336 | Seen a psychiatrist for nerves, anxiety, tension or depression | mood_behaviour | 0.2872 | 4.74E-02 | 1.42E-09 |
| ukb-b-15378 | Had other major operations | diseases | 0.3599 | 5.95E-02 | 1.51E-09 |
| ukb-b-10753 | Diabetes diagnosed by doctor | diseases | 0.2614 | 4.34E-02 | 1.75E-09 |
| ukb-b-5192 | Time spent watching television (TV) | activity_fitness_sleep | 0.2373 | 3.94E-02 | 1.79E-09 |

| GWAS_ID | Trait | Group | $r_g$ | se | p |
|---|---|---|---|---|---|
| ukb-b-11495 | Types of physical activity in last 4 weeks: Light DIY (eg: pruning, watering the lawn) | activity_fitness_sleep | -0.2746 | 4.57E-02 | 1.82E-09 |
| ukb-b-4263 | Number of full brothers | family_memb_health | 0.3825 | 6.37E-02 | 1.94E-09 |
| ukb-a-477 | Pain type(s) experienced in last month: Pain all over the body | pain_skeletal | 0.467 | 7.79E-02 | 1.99E-09 |
| ukb-b-18162 | Illness, injury, bereavement, stress in last 2 years: Serious illness, injury or assault to yourself | work_stress | 0.3991 | 6.68E-02 | 2.35E-09 |
| ukb-b-3957 | Sleeplessness / insomnia | work_stress | 0.2537 | 4.25E-02 | 2.47E-09 |
| ukb-b-11605 | Medication for pain relief, constipation, heartburn: Omeprazole (e.g. Zanprol) | drugs_supplements | 0.3194 | 5.37E-02 | 2.66E-09 |
| ukb-b-18099 | Qualifications: O levels/GCSEs or equivalent | education_intelligence | -0.3046 | 5.13E-02 | 2.84E-09 |
| ukb-b-4461 | Job involves mainly walking or standing | activity_fitness_sleep | 0.2498 | 4.20E-02 | 2.85E-09 |
| ieu-b-109 | HDL cholesterol | biomarker | -0.1966 | 3.32E-02 | 3.06E-09 |
| ukb-b-13354 | Whole body fat-free mass | anthropometry | 0.1548 | 2.62E-02 | 3.47E-09 |
| ieu-a-836 | College completion | education_intelligence | -0.3168 | 5.37E-02 | 3.64E-09 |
| ukb-b-14540 | Whole body water mass | anthropometry | 0.1531 | 2.61E-02 | 4.19E-09 |
| ukb-b-11461 | Medication for pain relief, constipation, heartburn: Laxatives (e.g. Dulcolax, Senokot) | drugs_supplements | 0.403 | 6.88E-02 | 4.75E-09 |
| ieu-b-24 | Age Of Smoking Initiation | smoking | -0.3092 | 5.33E-02 | 6.70E-09 |
| ukb-b-6019 | Number of cigarettes previously smoked daily | smoking | 0.3486 | 6.02E-02 | 7.19E-09 |
| ukb-b-10011 | Townsend deprivation index at recruitment | work_stress | 0.2997 | 5.19E-02 | 7.80E-09 |
| ukb-b-8463 | Back pain for 3+ months | pain_skeletal | 0.4472 | 7.75E-02 | 8.09E-09 |
| ukb-a-569 | Diagnoses - main ICD10: M54 Dorsalgia | diseases | 0.5167 | 8.97E-02 | 8.48E-09 |
| ukb-b-11578 | Chest pain or discomfort walking normally | pain_skeletal | 0.4529 | 7.93E-02 | 1.11E-08 |
| ukb-b-3822 | Frequency of depressed mood in last 2 weeks | mood_behaviour | 0.3084 | 5.41E-02 | 1.19E-08 |
| ukb-b-19657 | Forced expiratory volume in 1-second (FEV1) | lung_function | -0.1897 | 3.35E-02 | 1.47E-08 |
| ukb-b-11141 | Forced expiratory volume in 1-second (FEV1), Best measure | lung_function | -0.1883 | 3.36E-02 | 2.00E-08 |
| ieu-a-72 | Waist-to-hip ratio | anthropometry | 0.2802 | 0.05 | 2.08E-08 |
| ukb-b-19921 | Impedance of whole body | anthropometry | -0.1722 | 3.09E-02 | 2.42E-08 |
| ukb-b-10505 | Qualifications: CSEs or equivalent | education_intelligence | 0.3131 | 5.65E-02 | 3.05E-08 |
| ukb-b-5776 | Daytime dozing / sleeping (narcolepsy) | activity_fitness_sleep | 0.2197 | 3.97E-02 | 3.08E-08 |
| ukb-b-9937 | Had major operations | diseases | 0.4083 | 7.41E-02 | 3.63E-08 |
| ukb-b-14180 | Mood swings | mood_behaviour | 0.246 | 4.48E-02 | 3.90E-08 |
| ukb-b-13352 | Vascular/heart problems diagnosed by doctor: None of the above | diseases | -0.2095 | 3.83E-02 | 4.59E-08 |
| ukb-b-1209 | Number of live births | gynecology_fertility | 0.2394 | 4.40E-02 | 5.43E-08 |
| ukb-b-13405 | Forced expiratory volume in 1-second (FEV1), predicted percentage | lung_function | -0.2097 | 3.86E-02 | 5.49E-08 |

| GWAS_ID | Trait | Group | $r_g$ | se | p |
|---|---|---|---|---|---|
| ukb-b-2091 | Noisy workplace | work_stress | 0.3295 | 6.07E-02 | 5.65E-08 |
| ukb-b-13423 | Breastfed as a baby | infancy_puberty | -0.2981 | 5.51E-02 | 6.29E-08 |
| ukb-b-19809 | Fed-up feelings | mood_behaviour | 0.2606 | 4.86E-02 | 8.13E-08 |
| ukb-b-18103 | Pulse rate, automated reading | cardiovascular | 0.1692 | 3.18E-02 | 1.06E-07 |
| ukb-b-5238 | Fluid intelligence score | education_intelligence | -0.1984 | 3.73E-02 | 1.07E-07 |
| ukb-b-13799 | Qualifications: Other professional qualifications eg: nursing, teaching | education_intelligence | -0.2357 | 4.47E-02 | 1.31E-07 |
| ukb-b-5239 | Average weekly red wine intake | alcohol | -0.2335 | 4.44E-02 | 1.43E-07 |
| ukb-b-19379 | Impedance of arm (left) | anthropometry | -0.1634 | 3.13E-02 | 1.76E-07 |
| ukb-b-18994 | Miserableness | mood_behaviour | 0.2427 | 4.65E-02 | 1.76E-07 |
| ukb-b-7926 | Qualifications: NVQ or HND or HNC or equivalent | education_intelligence | 0.3158 | 6.15E-02 | 2.78E-07 |
| ukb-b-6244 | Exposure to tobacco smoke outside home | smoking | 0.2718 | 5.29E-02 | 2.86E-07 |
| ukb-b-16878 | Alcohol usually taken with meals | alcohol | -0.2282 | 4.47E-02 | 3.38E-07 |
| ukb-b-11413 | Pain type(s) experienced in last month: Stomach or abdominal pain | pain_skeletal | 0.3258 | 6.39E-02 | 3.44E-07 |
| ukb-b-14068 | Impedance of leg (left) | anthropometry | -0.1611 | 3.17E-02 | 3.60E-07 |
| met-d-L_HDL_PL_pct | Phospholipids to total lipids ratio in large HDL | NMR_metabo_UKBB | 0.27 | 5.32E-02 | 3.78E-07 |
| ebi-a-GCST005903 | Major depressive disorder (ICD-10 coded) | diseases | 0.4433 | 8.75E-02 | 4.07E-07 |
| ukb-b-960 | Smoking/smokers in household | smoking | 0.3704 | 7.32E-02 | 4.19E-07 |
| ukb-b-14177 | Vascular/heart problems diagnosed by doctor: High blood pressure | diseases | 0.1921 | 3.80E-02 | 4.29E-07 |
| ukb-b-4616 | Nap during day | activity_fitness_sleep | 0.1677 | 3.36E-02 | 5.77E-07 |
| ukb-b-8714 | Vascular/heart problems diagnosed by doctor: Stroke | diseases | 0.5241 | 0.1057 | 7.20E-07 |
| ukb-b-20379 | Medication for cholesterol, blood pressure, diabetes, or take exogenous hormones: None of the above | drugs_supplements | -0.2477 | 5.02E-02 | 7.88E-07 |
| met-d-PUFA_pct | Ratio of polyunsaturated fatty acids to total fatty acids | NMR_metabo_UKBB | -0.2917 | 5.93E-02 | 8.58E-07 |
| ukb-b-17557 | Types of transport used (excluding work): Cycle | activity_fitness_sleep | -0.2467 | 5.02E-02 | 8.83E-07 |
| ebi-a-GCST005195 | Coronary artery disease | diseases | 0.2224 | 4.54E-02 | 9.72E-07 |
| ukb-b-298 | Frequency of stair climbing in last 4 weeks | activity_fitness_sleep | -0.2269 | 4.66E-02 | 1.10E-06 |
| ukb-b-7137 | Medication for pain relief, constipation, heartburn: Aspirin | drugs_supplements | 0.2841 | 5.85E-02 | 1.22E-06 |
| ukb-b-17409 | Trunk fat-free mass | anthropometry | 0.123 | 2.56E-02 | 1.61E-06 |
| ukb-b-1712 | Job involves shift work | work_stress | 0.2888 | 6.02E-02 | 1.63E-06 |
| ukb-b-7376 | Impedance of leg (right) | anthropometry | -0.1501 | 3.13E-02 | 1.64E-06 |
| ukb-b-15787 | Transport type for commuting to job workplace: Cycle | activity_fitness_sleep | -0.2815 | 5.88E-02 | 1.71E-06 |
| ukb-b-5779 | Alcohol intake frequency. | alcohol | 0.1806 | 3.79E-02 | 1.88E-06 |

| GWAS_ID | Trait | Group | $r_g$ | se | p |
|---|---|---|---|---|---|
| ukb-a-582 | Diagnoses - main ICD10: R10 Abdominal and pelvic pain | diseases | 0.3726 | 7.84E-02 | 1.98E-06 |
| ukb-b-17155 | Transport type for commuting to job workplace: Public transport | activity_fitness_sleep | -0.3011 | 6.34E-02 | 2.02E-06 |
| met-d-L_LDL_TG | Triglycerides in large LDL | NMR_metabo_UKBB | 0.2027 | 4.27E-02 | 2.11E-06 |
| ukb-b-5593 | Number of full sisters | family_memb_health | 0.2936 | 6.20E-02 | 2.18E-06 |
| ukb-b-8476 | Loneliness, isolation | mood_behaviour | 0.2311 | 4.89E-02 | 2.32E-06 |
| met-d-L_VLDL_PL_pct | Phospholipids to total lipids ratio in large VLDL | NMR_metabo_UKBB | 0.2329 | 4.94E-02 | 2.44E-06 |
| ukb-b-17400 | Snoring | activity_fitness_sleep | -0.1778 | 3.77E-02 | 2.45E-06 |
| met-d-Albumin | Albumin | NMR_metabo_UKBB | -0.2702 | 5.73E-02 | 2.47E-06 |
| ukb-a-224 | Smoking status: Previous | smoking | 0.2177 | 4.63E-02 | 2.52E-06 |
| ukb-b-4462 | Exposure to tobacco smoke at home | smoking | 0.3407 | 7.26E-02 | 2.69E-06 |
| ukb-b-7551 | Maximum workload during fitness test | cardiovascular | -0.3984 | 8.52E-02 | 2.91E-06 |
| ukb-b-7663 | Types of physical activity in last 4 weeks: Strenuous sports | activity_fitness_sleep | -0.2588 | 5.56E-02 | 3.22E-06 |
| ukb-b-7859 | Impedance of arm (right) | anthropometry | -0.1478 | 3.18E-02 | 3.26E-06 |
| ukb-b-15957 | Types of transport used (excluding work): Public transport | activity_fitness_sleep | -0.2507 | 5.39E-02 | 3.35E-06 |
| met-d-GlycA | Glycoprotein acetyls | NMR_metabo_UKBB | 0.2472 | 5.34E-02 | 3.76E-06 |
| ukb-b-13184 | Types of physical activity in last 4 weeks: Heavy DIY (eg: weeding, lawn mowing, carpentry, digging) | activity_fitness_sleep | -0.207 | 4.49E-02 | 4.10E-06 |
| met-d-Omega_6_pct | Ratio of omega-6 fatty acids to total fatty acids | NMR_metabo_UKBB | -0.2642 | 5.75E-02 | 4.35E-06 |
| ebi-a-GCST005843 | Ischemic stroke | diseases | 0.2589 | 5.64E-02 | 4.38E-06 |
| ukb-b-8468 | Vascular/heart problems diagnosed by doctor: Angina | diseases | 0.2718 | 5.95E-02 | 4.91E-06 |
| met-d-XS_VLDL_PL_pct | Phospholipids to total lipids ratio in very small VLDL | NMR_metabo_UKBB | 0.2139 | 4.77E-02 | 7.31E-06 |
| met-d-M_HDL_PL_pct | Phospholipids to total lipids ratio in medium HDL | NMR_metabo_UKBB | 0.2167 | 4.86E-02 | 8.15E-06 |
| ukb-d-M25 | Diagnoses - main ICD10: M25 Other joint disorders, not elsewhere classified | diseases | 0.5857 | 0.1313 | 8.22E-06 |
| met-d-L_HDL_CE_pct | Cholesteryl esters to total lipids ratio in large HDL | NMR_metabo_UKBB | -0.2314 | 5.20E-02 | 8.76E-06 |
| met-d-M_LDL_C_pct | Cholesterol to total lipids ratio in medium LDL | NMR_metabo_UKBB | -0.2902 | 6.59E-02 | 1.07E-05 |
| ukb-b-703 | Duration walking for pleasure | activity_fitness_sleep | -0.2478 | 5.63E-02 | 1.08E-05 |
| ukb-d-20117_2 | Alcohol drinker status: Current | alcohol | -0.2533 | 5.80E-02 | 1.25E-05 |
| ukb-d-T84 | Diagnoses - main ICD10: T84 Complications of internal orthopaedic prosthetic devices, implants and grafts | diseases | 0.4815 | 0.1103 | 1.26E-05 |
| bbj-a-81 | Smoking behaviors : Smoking cessation | smoking | -0.3475 | 8.05E-02 | 1.58E-05 |
| met-d-PUFA_by_MUFA | Ratio of polyunsaturated fatty acids to monounsaturated fatty acids | NMR_metabo_UKBB | -0.268 | 6.22E-02 | 1.65E-05 |
| ukb-b-12849 | Mouth/teeth dental problems: Loose teeth | dental | 0.2903 | 6.76E-02 | 1.74E-05 |

| GWAS_ID | Trait | Group | $r_g$ | se | p |
|---|---|---|---|---|---|
| ieu-a-963 | Former vs current smoker | smoking | -0.451 | 0.1051 | 1.78E-05 |
| met-d-IDL_TG | Triglycerides in IDL | NMR_metabo_UKBB | 0.1813 | 4.23E-02 | 1.78E-05 |
| ieu-b-111 | triglycerides | biomarker | 0.1875 | 4.37E-02 | 1.82E-05 |
| ukb-b-18009 | Medication for cholesterol, blood pressure, diabetes, or take exogenous hormones: Blood pressure medication | drugs_supplements | 0.1984 | 4.63E-02 | 1.84E-05 |
| met-d-L_LDL_C_pct | Cholesterol to total lipids ratio in large LDL | NMR_metabo_UKBB | -0.2603 | 6.09E-02 | 1.94E-05 |
| ukb-a-528 | Diagnoses - main ICD10: G56 Mononeuropathies of upper limb | diseases | 0.3009 | 7.07E-02 | 2.09E-05 |
| ukb-b-13346 | Fractured/broken bones in last 5 years | pain_skeletal | 0.2681 | 6.31E-02 | 2.13E-05 |
| met-d-LDL_TG | Triglycerides in LDL | NMR_metabo_UKBB | 0.1887 | 4.44E-02 | 2.13E-05 |
| ukb-b-15272 | Ever manic/hyper for 2 days | mood_behaviour | 0.3831 | 9.01E-02 | 2.14E-05 |
| met-d-SFA_pct | Ratio of saturated fatty acids to total fatty acids | NMR_metabo_UKBB | 0.2592 | 6.18E-02 | 2.77E-05 |
| ukb-b-8196 | Ever highly irritable/argumentative for 2 days | mood_behaviour | 0.2627 | 6.31E-02 | 3.16E-05 |
| ukb-b-11025 | Transport type for commuting to job workplace: Car/motor vehicle | activity_fitness_sleep | 0.2751 | 6.61E-02 | 3.18E-05 |
| met-d-L_LDL_TG_pct | Triglycerides to total lipids ratio in large LDL | NMR_metabo_UKBB | 0.2566 | 6.18E-02 | 3.25E-05 |
| ukb-d-2664_1 | Reason for reducing amount of alcohol drunk: Illness or ill health | alcohol | 0.7359 | 0.178 | 3.55E-05 |
| ieu-b-107 | apolipoprotein A-I | biomarker | -0.1456 | 3.54E-02 | 3.88E-05 |
| ukb-b-469 | Number of cigarettes currently smoked daily (current cigarette smokers) | smoking | 0.3845 | 9.47E-02 | 4.90E-05 |
| ukb-b-5716 | Average weekly champagne plus white wine intake | alcohol | -0.2134 | 5.26E-02 | 4.92E-05 |
| met-d-XL_HDL_FC_pct | Free cholesterol to total lipids ratio in very large HDL | NMR_metabo_UKBB | 0.1883 | 4.64E-02 | 5.02E-05 |
| met-d-Unsaturation | Degree of unsaturation | NMR_metabo_UKBB | -0.2526 | 6.24E-02 | 5.17E-05 |
| ukb-b-17805 | Medication for cholesterol, blood pressure, diabetes, or take exogenous hormones: Cholesterol lowering medication | drugs_supplements | 0.2185 | 5.41E-02 | 5.33E-05 |
| ukb-b-8888 | Medication for pain relief, constipation, heartburn: Ibuprofen (e.g. Nurofen) | drugs_supplements | 0.2258 | 5.59E-02 | 5.34E-05 |

**Table S10. Results of Mendelian randomization analysis (exposure -> pneumonia).**

Column headers: exposure, name of the exposure trait as in IEU Open GWAS Project website (https://gwas.mrcieu.ac.uk/) and study ID for the site; method, MR method used; nsnp, number of valid SNPs used for analysis; p, p-value; or, odds ratio; or_lci95; odds ratio lower 95 % confidence interval; or_uci95, odds ratio upper 95 % confidence interval; Q_pval, Q-test p-value (for testing heterogeneity); egger_intercept, MR Egger intercept; and ple_pval, pleiotropy p-value.

| exposure | method | nsnp | pval | or | or_lci95 | or_uci95 | Q_pval | egger_intercept | ple_pval |
|---|---|---|---|---|---|---|---|---|---|
| Acetate \|\| id:met-d-Acetate | MR Egger | 7 | 7.84E-01 | 1.12 | 0.53 | 2.35 | 6.38E-04 | -9.36E-03 | 0.533 |
| Acetate \|\| id:met-d-Acetate | Inverse variance weighted | 7 | 2.92E-01 | 0.88 | 0.69 | 1.12 | 6.50E-04 | | |
| Age at first live birth \|\| id:ukb-b-12405 | MR Egger | 30 | 5.66E-01 | 0.84 | 0.46 | 1.52 | 8.24E-02 | 1.61E-03 | 0.818 |
| Age at first live birth \|\| id:ukb-b-12405 | Inverse variance weighted | 30 | 4.40E-02 | 0.90 | 0.81 | 1.00 | 1.02E-01 | | |
| Albumin \|\| id:met-d-Albumin | MR Egger | 23 | 7.04E-02 | 1.13 | 1.00 | 1.28 | 1.77E-03 | -7.29E-03 | 0.031 |
| Albumin \|\| id:met-d-Albumin | Inverse variance weighted | 23 | 8.31E-01 | 1.01 | 0.92 | 1.10 | 7.72E-05 | | |
| Alcohol drinker status: Current \|\| id:ukb-d-20117_2 | MR Egger | 3 | 4.26E-01 | 75.88 | 0.09 | 62256.49 | 6.91E-01 | -1.73E-02 | 0.447 |
| Alcohol drinker status: Current \|\| id:ukb-d-20117_2 | Inverse variance weighted | 3 | 6.46E-01 | 1.60 | 0.22 | 11.80 | 4.60E-01 | | |
| apolipoprotein A-I \|\| id:ieu-b-107 | MR Egger | 247 | 5.68E-03 | 1.07 | 1.02 | 1.13 | 9.15E-06 | -1.70E-03 | 0.030 |
| apolipoprotein A-I \|\| id:ieu-b-107 | Inverse variance weighted | 247 | 8.33E-02 | 1.03 | 1.00 | 1.06 | 3.68E-06 | | |
| Blood clot, DVT, bronchitis, emphysema, asthma, rhinitis, eczema, allergy diagnosed by doctor: Blood clot in the lung \|\| id:ukb-b-13704 | MR Egger | 6 | 1.42E-01 | 678.15 | 0.62 | 741834.75 | 9.87E-01 | -5.48E-03 | 0.516 |
| Blood clot, DVT, bronchitis, emphysema, asthma, rhinitis, eczema, allergy diagnosed by doctor: Blood clot in the lung \|\| id:ukb-b-13704 | Inverse variance weighted | 6 | 6.03E-04 | 62.01 | 5.87 | 655.71 | 9.74E-01 | | |
| Body mass index (BMI) \|\| id:ukb-b-19953 | MR Egger | 387 | 4.22E-01 | 1.04 | 0.94 | 1.15 | 2.73E-05 | 2.35E-03 | 0.013 |
| Body mass index (BMI) \|\| id:ukb-b-19953 | Inverse variance weighted | 387 | 1.39E-16 | 1.17 | 1.13 | 1.22 | 1.08E-05 | | |
| Chest pain or discomfort \|\| id:ukb-b-10591 | MR Egger | 7 | 6.89E-01 | 0.05 | 0.00 | 54520.83 | 1.78E-03 | 2.22E-02 | 0.586 |
| Chest pain or discomfort \|\| id:ukb-b-10591 | Inverse variance weighted | 7 | 2.03E-01 | 2.97 | 0.56 | 15.82 | 2.27E-03 | | |
| Childhood asthma (age<16) \|\| id:ukb-d-ASTHMA_CHILD | Inverse variance weighted | 2 | 4.16E-01 | 245.37 | 0.00 | 141791790.67 | 6.99E-02 | NA | NA |
| Diagnoses - main ICD10: J33 Nasal polyp \|\| id:ukb-a-541 | MR Egger | 6 | 8.17E-01 | 0.13 | 0.00 | 1786414.62 | 4.32E-01 | 1.56E-02 | 0.215 |
| Diagnoses - main ICD10: J33 Nasal polyp \|\| id:ukb-a-541 | Inverse variance weighted | 6 | 1.37E-05 | 20236.99 | 232.00 | 1765236.52 | 3.08E-01 | | |
| Diagnoses - main ICD10: J44 Other chronic obstructive pulmonary disease \|\| id:ukb-a-543 | Inverse variance weighted | 2 | 9.44E-01 | 0.71 | 0.00 | 8756.62 | 2.72E-01 | NA | NA |

| exposure | method | nsnp | pval | or | or_lci95 | or_uci95 | Q_pval | egger_intercept | ple_pval |
|---|---|---|---|---|---|---|---|---|---|
| Docosahexaenoic acid \|\| id:met-d-DHA | MR Egger | 38 | 4.80E-02 | 1.06 | 1.00 | 1.11 | 2.84E-03 | -3.69E-04 | 0.866 |
| Docosahexaenoic acid \|\| id:met-d-DHA | Inverse variance weighted | 38 | 5.73E-03 | 1.05 | 1.02 | 1.09 | 3.87E-03 | | |
| Forced expiratory volume in 1-second (FEV1) \|\| id:ukb-b-19657 | MR Egger | 232 | 3.88E-01 | 1.09 | 0.90 | 1.33 | 2.21E-07 | -1.15E-03 | 0.458 |
| Forced expiratory volume in 1-second (FEV1) \|\| id:ukb-b-19657 | Inverse variance weighted | 232 | 6.23E-01 | 1.02 | 0.95 | 1.08 | 2.37E-07 | | |
| Forced expiratory volume in 1-second (FEV1), Best measure \|\| id:ukb-b-11141 | MR Egger | 199 | 6.30E-01 | 1.05 | 0.86 | 1.29 | 9.70E-07 | -1.98E-04 | 0.909 |
| Forced expiratory volume in 1-second (FEV1), Best measure \|\| id:ukb-b-11141 | Inverse variance weighted | 199 | 2.36E-01 | 1.04 | 0.98 | 1.11 | 1.22E-06 | | |
| Forced expiratory volume in 1-second (FEV1), predicted percentage \|\| id:ukb-b-13405 | MR Egger | 81 | 5.32E-01 | 1.12 | 0.78 | 1.62 | 1.37E-12 | -4.01E-03 | 0.418 |
| Forced expiratory volume in 1-second (FEV1), predicted percentage \|\| id:ukb-b-13405 | Inverse variance weighted | 81 | 4.31E-01 | 0.97 | 0.90 | 1.05 | 1.31E-12 | | |
| Forced vital capacity (FVC) \|\| id:ukb-b-7953 | MR Egger | 276 | 8.18E-02 | 1.14 | 0.98 | 1.32 | 2.26E-03 | -2.09E-03 | 0.069 |
| Forced vital capacity (FVC) \|\| id:ukb-b-7953 | Inverse variance weighted | 276 | 8.94E-01 | 1.00 | 0.95 | 1.06 | 1.58E-03 | | |
| Forced vital capacity (FVC), Best measure \|\| id:ukb-b-14713 | MR Egger | 263 | 8.03E-01 | 1.02 | 0.88 | 1.18 | 3.59E-05 | 2.18E-04 | 0.859 |
| Forced vital capacity (FVC), Best measure \|\| id:ukb-b-14713 | Inverse variance weighted | 263 | 2.61E-01 | 1.03 | 0.98 | 1.09 | 4.24E-05 | | |
| Fractured/broken bones in last 5 years \|\| id:ukb-b-13346 | MR Egger | 14 | 4.37E-01 | 0.38 | 0.03 | 4.09 | 3.79E-02 | 6.47E-03 | 0.296 |
| Fractured/broken bones in last 5 years \|\| id:ukb-b-13346 | Inverse variance weighted | 14 | 5.41E-01 | 1.31 | 0.56 | 3.07 | 2.98E-02 | | |
| Frequency of tiredness / lethargy in last 2 weeks \|\| id:ukb-b-929 | MR Egger | 37 | 7.08E-01 | 1.18 | 0.50 | 2.78 | 6.06E-02 | 2.85E-03 | 0.583 |
| Frequency of tiredness / lethargy in last 2 weeks \|\| id:ukb-b-929 | Inverse variance weighted | 37 | 2.62E-05 | 1.49 | 1.24 | 1.80 | 6.98E-02 | | |
| Glycoprotein acetyls \|\| id:met-d-GlycA | MR Egger | 43 | 2.76E-01 | 0.96 | 0.90 | 1.03 | 2.16E-01 | 5.39E-03 | 0.012 |
| Glycoprotein acetyls \|\| id:met-d-GlycA | Inverse variance weighted | 43 | 1.00E-01 | 1.04 | 0.99 | 1.08 | 7.50E-02 | | |
| HbA1c \|\| id:ieu-b-4842 | MR Egger | 25 | 8.73E-01 | 1.01 | 0.93 | 1.09 | 2.40E-01 | -1.12E-03 | 0.697 |
| HbA1c \|\| id:ieu-b-4842 | Inverse variance weighted | 25 | 6.88E-01 | 0.99 | 0.96 | 1.03 | 2.79E-01 | | |
| HDL cholesterol \|\| id:ieu-b-109 | MR Egger | 295 | 2.07E-02 | 1.06 | 1.01 | 1.11 | 7.42E-06 | -1.69E-03 | 0.015 |
| HDL cholesterol \|\| id:ieu-b-109 | Inverse variance weighted | 295 | 4.79E-01 | 1.01 | 0.98 | 1.04 | 2.46E-06 | | |
| Illness, injury, bereavement, stress in last 2 years: Financial difficulties \|\| id:ukb-b-9667 | MR Egger | 8 | 6.33E-01 | 0.49 | 0.03 | 7.84 | 3.07E-01 | 8.08E-03 | 0.298 |
| Illness, injury, bereavement, stress in last 2 years: Financial difficulties \|\| id:ukb-b-9667 | Inverse variance weighted | 8 | 1.09E-01 | 2.22 | 0.84 | 5.91 | 2.75E-01 | | |
| Mouth/teeth dental problems: None of the above \|\| id:ukb-b-17601 | MR Egger | 12 | 4.92E-01 | 1.67 | 0.41 | 6.89 | 5.75E-02 | -9.25E-03 | 0.156 |
| Mouth/teeth dental problems: None of the above \|\| id:ukb-b-17601 | Inverse variance weighted | 12 | 9.03E-02 | 0.60 | 0.34 | 1.08 | 2.39E-02 | | |
| Overall health rating \|\| id:ukb-b-6306 | MR Egger | 96 | 2.29E-03 | 2.65 | 1.44 | 4.87 | 1.15E-01 | -5.38E-03 | 0.108 |

| exposure | method | nsnp | pval | or | or_lci95 | or_uci95 | Q_pval | egger_intercept | ple_pval |
|---|---|---|---|---|---|---|---|---|---|
| Overall health rating \|\| id:ukb-b-6306 | Inverse variance weighted | 96 | 4.99E-15 | 1.62 | 1.43 | 1.82 | 9.20E-02 | | |
| Pack years of smoking \|\| id:ukb-b-10831 | MR Egger | 10 | 2.12E-03 | 1.76 | 1.37 | 2.26 | 5.57E-01 | -9.69E-03 | 0.063 |
| Pack years of smoking \|\| id:ukb-b-10831 | Inverse variance weighted | 10 | 1.97E-07 | 1.37 | 1.22 | 1.55 | 2.46E-01 | | |
| Parental longevity (combined parental age at death) \|\| id:ebi-a-GCST006702 | MR Egger | 7 | 3.81E-01 | 2.70 | 0.36 | 20.48 | 4.22E-14 | -2.74E-02 | 0.361 |
| Parental longevity (combined parental age at death) \|\| id:ebi-a-GCST006702 | Inverse variance weighted | 7 | 9.64E-01 | 1.02 | 0.51 | 2.02 | 1.73E-16 | | |
| Peak expiratory flow (PEF) \|\| id:ukb-b-12019 | MR Egger | 124 | 9.70E-01 | 1.01 | 0.73 | 1.38 | 5.04E-07 | -1.35E-04 | 0.956 |
| Peak expiratory flow (PEF) \|\| id:ukb-b-12019 | Inverse variance weighted | 124 | 9.61E-01 | 1.00 | 0.91 | 1.10 | 6.76E-07 | | |
| Platelet crit \|\| id:ukb-d-30090_irnt | MR Egger | 296 | 1.20E-01 | 1.04 | 0.99 | 1.09 | 1.62E-06 | -2.56E-04 | 0.739 |
| Platelet crit \|\| id:ukb-d-30090_irnt | Inverse variance weighted | 296 | 2.38E-02 | 1.03 | 1.00 | 1.06 | 1.90E-06 | | |
| Saturated fatty acids \|\| id:met-d-SFA | MR Egger | 43 | 2.08E-01 | 1.04 | 0.98 | 1.11 | 3.71E-01 | 7.19E-05 | 0.968 |
| Saturated fatty acids \|\| id:met-d-SFA | Inverse variance weighted | 43 | 1.79E-02 | 1.04 | 1.01 | 1.08 | 4.13E-01 | | |
| Seen doctor (GP) for nerves, anxiety, tension or depression \|\| id:ukb-b-6991 | MR Egger | 37 | 4.33E-03 | 24.46 | 3.14 | 190.77 | 6.13E-02 | -1.64E-02 | 0.022 |
| Seen doctor (GP) for nerves, anxiety, tension or depression \|\| id:ukb-b-6991 | Inverse variance weighted | 37 | 1.04E-04 | 2.05 | 1.42 | 2.94 | 1.51E-02 | | |
| Smoking status: Current \|\| id:ukb-a-225 | MR Egger | 15 | 3.49E-01 | 19.29 | 0.05 | 7570.80 | 1.15E-05 | -1.10E-02 | 0.486 |
| Smoking status: Current \|\| id:ukb-a-225 | Inverse variance weighted | 15 | 1.41E-01 | 2.25 | 0.76 | 6.62 | 1.14E-05 | | |
| Sodium in urine \|\| id:ukb-a-335 | MR Egger | 28 | 9.45E-01 | 1.03 | 0.49 | 2.14 | 1.40E-02 | 2.76E-03 | 0.682 |
| Sodium in urine \|\| id:ukb-a-335 | Inverse variance weighted | 28 | 3.33E-02 | 1.19 | 1.01 | 1.41 | 1.78E-02 | | |
| Squamous cell lung cancer \|\| id:ieu-a-967 | MR Egger | 4 | 8.54E-01 | 1.03 | 0.78 | 1.36 | 1.66E-07 | -1.80E-04 | 0.997 |
| Squamous cell lung cancer \|\| id:ieu-a-967 | Inverse variance weighted | 4 | 5.00E-01 | 1.03 | 0.95 | 1.12 | 7.64E-07 | | |
| triglycerides \|\| id:ieu-b-111 | MR Egger | 262 | 4.65E-01 | 0.98 | 0.94 | 1.03 | 9.11E-06 | 8.71E-04 | 0.216 |
| triglycerides \|\| id:ieu-b-111 | Inverse variance weighted | 262 | 7.94E-01 | 1.00 | 0.97 | 1.03 | 7.79E-06 | | |
| Type 2 diabetes \|\| id:ebi-a-GCST006867 | MR Egger | 108 | 9.57E-01 | 1.00 | 0.96 | 1.04 | 4.44E-03 | 5.71E-05 | 0.972 |
| Type 2 diabetes \|\| id:ebi-a-GCST006867 | Inverse variance weighted | 108 | 9.58E-01 | 1.00 | 0.98 | 1.02 | 5.36E-03 | | |
| Usual walking pace \|\| id:ukb-b-4711 | MR Egger | 51 | 4.00E-01 | 0.73 | 0.35 | 1.52 | 4.69E-01 | -5.01E-04 | 0.886 |
| Usual walking pace \|\| id:ukb-b-4711 | Inverse variance weighted | 51 | 4.23E-05 | 0.69 | 0.58 | 0.82 | 5.08E-01 | | |
| Vascular/heart problems diagnosed by doctor: Angina \|\| id:ukb-b-8468 | MR Egger | 20 | 1.62E-01 | 0.09 | 0.00 | 2.33 | 7.11E-03 | 5.37E-03 | 0.327 |
| Vascular/heart problems diagnosed by doctor: Angina \|\| id:ukb-b-8468 | Inverse variance weighted | 20 | 1.62E-01 | 0.42 | 0.12 | 1.42 | 5.93E-03 | | |
| Vascular/heart problems diagnosed by doctor: Heart attack \|\| id:ukb-b-11590 | MR Egger | 13 | 3.20E-02 | 0.01 | 0.00 | 0.36 | 2.17E-01 | 1.44E-02 | 0.045 |

| exposure | method | nsnp | pval | or | or_lci95 | or_uci95 | Q_pval | egger_intercept | ple_pval |
|---|---|---|---|---|---|---|---|---|---|
| Vascular/heart problems diagnosed by doctor: Heart attack \|\| id:ukb-b-11590 | Inverse variance weighted | 13 | 4.13E-01 | 0.52 | 0.11 | 2.49 | 5.08E-02 | | |
| Vascular/heart problems diagnosed by doctor: High blood pressure \|\| id:ukb-b-14177 | MR Egger | 202 | 3.38E-01 | 1.17 | 0.85 | 1.61 | 3.83E-04 | -7.95E-04 | 0.564 |
| Vascular/heart problems diagnosed by doctor: High blood pressure \|\| id:ukb-b-14177 | Inverse variance weighted | 202 | 2.46E-01 | 1.07 | 0.95 | 1.20 | 4.25E-04 | | |
| Waist-to-hip ratio \|\| id:ieu-a-72 | MR Egger | 29 | 5.80E-01 | 0.89 | 0.58 | 1.35 | 1.45E-01 | 1.24E-03 | 0.819 |
| Waist-to-hip ratio \|\| id:ieu-a-72 | Inverse variance weighted | 29 | 1.31E-01 | 0.93 | 0.85 | 1.02 | 1.75E-01 | | |
| Wheeze or whistling in the chest in last year \|\| id:ukb-b-18335 | MR Egger | 42 | 9.35E-02 | 6.29 | 0.77 | 51.25 | 5.92E-12 | -6.52E-03 | 0.374 |
| Wheeze or whistling in the chest in last year \|\| id:ukb-b-18335 | Inverse variance weighted | 42 | 3.72E-04 | 2.47 | 1.50 | 4.06 | 4.18E-12 | | |
| Whole body fat mass \|\| id:ukb-b-19393 | MR Egger | 377 | 1.17E-02 | 1.15 | 1.03 | 1.27 | 1.81E-04 | 5.77E-04 | 0.549 |
| Whole body fat mass \|\| id:ukb-b-19393 | Inverse variance weighted | 377 | 1.41E-17 | 1.18 | 1.14 | 1.23 | 1.96E-04 | | |
| Whole body fat-free mass \|\| id:ukb-b-13354 | MR Egger | 481 | 3.69E-01 | 1.05 | 0.94 | 1.17 | 3.00E-06 | 1.33E-03 | 0.079 |
| Whole body fat-free mass \|\| id:ukb-b-13354 | Inverse variance weighted | 481 | 7.45E-09 | 1.15 | 1.10 | 1.20 | 2.06E-06 | | |
| Years of schooling \|\| id:ieu-a-1239 | MR Egger | 278 | 2.07E-02 | 0.75 | 0.59 | 0.96 | 4.70E-07 | 1.51E-03 | 0.374 |
| Years of schooling \|\| id:ieu-a-1239 | Inverse variance weighted | 278 | 1.37E-08 | 0.83 | 0.78 | 0.89 | 4.73E-07 | | |

**Table S11. Results of Mendelian randomization analysis (pneumonia -> outcome).**

Column headers: outcome, name of the outcome trait as in IEU Open GWAS Project website (https://gwas.mrcieu.ac.uk/) and study ID for the site; method, MR method used; nsnp, number of valid SNPs used for analysis; p, p-value; b, beta estimate; lo_ci, lower 95 % confidence interval; up_ci, upper 95 % confidence interval; Q_pval, Q-test p-value (for testing heterogeneity); egger_intercept, MR Egger intercept; and ple_pval, pleiotropy p-value.

| outcome | method | nsnp | b | pval | lo_ci | up_ci | Q | Q_pval | egger_intercept | ple_pval |
|---|---|---|---|---|---|---|---|---|---|---|
| Acetate \|\| id:met-d-Acetate | MR Egger | 11 | 0.413 | 5.87E-02 | 0.039 | 0.788 | 6.353 | 7.04E-01 | -1.71E-02 | 0.031 |
| Acetate \|\| id:met-d-Acetate | Inverse variance weighted | 11 | -0.063 | 1.55E-01 | -0.150 | 0.024 | 12.838 | 2.33E-01 | | |
| Albumin \|\| id:met-d-Albumin | MR Egger | 11 | 0.060 | 8.51E-01 | -0.547 | 0.667 | 23.963 | 4.36E-03 | -3.37E-03 | 0.764 |
| Albumin \|\| id:met-d-Albumin | Inverse variance weighted | 11 | -0.034 | 5.72E-01 | -0.152 | 0.084 | 24.218 | 7.04E-03 | | |
| apolipoprotein A-I \|\| id:ieu-b-107 | MR Egger | 11 | 0.919 | 3.86E-01 | -1.058 | 2.896 | 1133.133 | 3.29E-238 | -2.78E-02 | 0.454 |
| apolipoprotein A-I \|\| id:ieu-b-107 | Inverse variance weighted | 11 | 0.146 | 4.70E-01 | -0.250 | 0.543 | 1210.289 | 8.69E-254 | | |
| Blood clot, DVT, bronchitis, emphysema, asthma, rhinitis, eczema, allergy diagnosed by doctor: Blood clot in the lung \|\| id:ukb-b-13704 | MR Egger | 10 | 0.014 | 1.40E-01 | -0.003 | 0.031 | 5.221 | 7.34E-01 | -4.96E-04 | 0.149 |
| Blood clot, DVT, bronchitis, emphysema, asthma, rhinitis, eczema, allergy diagnosed by doctor: Blood clot in the lung \|\| id:ukb-b-13704 | Inverse variance weighted | 10 | 0.001 | 7.17E-01 | -0.003 | 0.004 | 7.777 | 5.57E-01 | | |
| Blood clot, DVT, bronchitis, emphysema, asthma, rhinitis, eczema, allergy diagnosed by doctor: Emphysema/chronic bronchitis \|\| id:ukb-b-16207 | MR Egger | 10 | 0.014 | 5.51E-01 | -0.031 | 0.059 | 26.917 | 7.31E-04 | -1.10E-04 | 0.895 |
| Blood clot, DVT, bronchitis, emphysema, asthma, rhinitis, eczema, allergy diagnosed by doctor: Emphysema/chronic bronchitis \|\| id:ukb-b-16207 | Inverse variance weighted | 10 | 0.011 | 1.45E-02 | 0.002 | 0.020 | 26.980 | 1.41E-03 | | |
| Body mass index (BMI) \|\| id:ukb-b-19953 | MR Egger | 10 | 0.042 | 8.82E-01 | -0.495 | 0.579 | 72.112 | 1.86E-12 | -5.10E-04 | 0.960 |
| Body mass index (BMI) \|\| id:ukb-b-19953 | Inverse variance weighted | 10 | 0.028 | 6.10E-01 | -0.080 | 0.136 | 72.137 | 5.79E-12 | | |
| Bring up phlegm/sputum/mucus on most days \|\| id:ukb-b-12841 | MR Egger | 10 | -0.098 | 4.79E-01 | -0.358 | 0.161 | 47.321 | 1.33E-07 | 3.44E-03 | 0.487 |
| Bring up phlegm/sputum/mucus on most days \|\| id:ukb-b-12841 | Inverse variance weighted | 10 | -0.004 | 8.84E-01 | -0.058 | 0.050 | 50.465 | 8.81E-08 | | |
| Childhood asthma (age<16) \|\| id:ukb-d-ASTHMA_CHILD | MR Egger | 11 | -0.005 | 6.86E-01 | -0.027 | 0.018 | 18.717 | 2.77E-02 | 2.37E-04 | 0.574 |
| Childhood asthma (age<16) \|\| id:ukb-d-ASTHMA_CHILD | Inverse variance weighted | 11 | 0.002 | 4.37E-01 | -0.003 | 0.006 | 19.426 | 3.52E-02 | | |
| circulating leptin levels \|\| id:ebi-a-GCST003367 | MR Egger | 7 | -0.075 | 8.53E-01 | -0.822 | 0.673 | 8.436 | 1.34E-01 | 2.33E-03 | 0.872 |

| | | | | | | | | | | |
|---|---|---|---|---|---|---|---|---|---|---|
| circulating leptin levels \|\| id:ebi-a-GCST003367 | Inverse variance weighted | 7 | -0.012 | 8.92E-01 | -0.183 | 0.159 | 8.484 | 2.05E-01 | | |
| Diagnoses - main ICD10: J33 Nasal polyp \|\| id:ukb-a-541 | MR Egger | 10 | -0.012 | 3.13E-01 | -0.034 | 0.010 | 16.800 | 3.23E-02 | 4.22E-04 | 0.327 |
| Diagnoses - main ICD10: J33 Nasal polyp \|\| id:ukb-a-541 | Inverse variance weighted | 10 | -0.001 | 7.92E-01 | -0.005 | 0.004 | 19.090 | 2.44E-02 | | |
| Diagnoses - main ICD10: J44 Other chronic obstructive pulmonary disease \|\| id:ukb-a-543 | MR Egger | 10 | 0.002 | 7.63E-01 | -0.011 | 0.015 | 9.976 | 2.67E-01 | -2.90E-05 | 0.902 |
| Diagnoses - main ICD10: J44 Other chronic obstructive pulmonary disease \|\| id:ukb-a-543 | Inverse variance weighted | 10 | 0.001 | 3.50E-01 | -0.001 | 0.004 | 9.996 | 3.51E-01 | | |
| Diagnoses - main ICD10: K21 Gastro-oesophageal reflux disease \|\| id:ukb-a-545 | MR Egger | 10 | 0.003 | 9.19E-01 | -0.044 | 0.049 | 17.107 | 2.90E-02 | -1.41E-04 | 0.873 |
| Diagnoses - main ICD10: K21 Gastro-oesophageal reflux disease \|\| id:ukb-a-545 | Inverse variance weighted | 10 | -0.001 | 7.78E-01 | -0.011 | 0.008 | 17.165 | 4.62E-02 | | |
| Diagnoses - main ICD10: R07 Pain in throat and chest \|\| id:ukb-a-581 | MR Egger | 10 | 0.006 | 8.08E-01 | -0.042 | 0.054 | 3.341 | 9.11E-01 | -4.26E-04 | 0.636 |
| Diagnoses - main ICD10: R07 Pain in throat and chest \|\| id:ukb-a-581 | Inverse variance weighted | 10 | -0.006 | 2.81E-01 | -0.016 | 0.005 | 3.582 | 9.37E-01 | | |
| Diagnoses - main ICD10: T84 Complications of internal orthopaedic prosthetic devices, implants and grafts \|\| id:ukb-d-T84 | MR Egger | 11 | -0.013 | 3.04E-01 | -0.037 | 0.011 | 10.871 | 2.85E-01 | 5.22E-04 | 0.256 |
| Diagnoses - main ICD10: T84 Complications of internal orthopaedic prosthetic devices, implants and grafts \|\| id:ukb-d-T84 | Inverse variance weighted | 11 | 0.001 | 6.35E-01 | -0.004 | 0.006 | 12.652 | 2.44E-01 | | |
| Forced expiratory volume in 1-second (FEV1) \|\| id:ukb-b-19657 | MR Egger | 10 | 0.081 | 7.10E-01 | -0.330 | 0.492 | 54.671 | 5.11E-09 | -6.12E-03 | 0.436 |
| Forced expiratory volume in 1-second (FEV1) \|\| id:ukb-b-19657 | Inverse variance weighted | 10 | -0.087 | 4.60E-02 | -0.173 | -0.002 | 59.268 | 1.85E-09 | | |
| Forced expiratory volume in 1-second (FEV1), Best measure \|\| id:ukb-b-11141 | MR Egger | 10 | 0.160 | 5.10E-01 | -0.295 | 0.616 | 55.812 | 3.07E-09 | -9.49E-03 | 0.284 |
| Forced expiratory volume in 1-second (FEV1), Best measure \|\| id:ukb-b-11141 | Inverse variance weighted | 10 | -0.100 | 4.54E-02 | -0.199 | -0.002 | 64.999 | 1.44E-10 | | |
| Forced expiratory volume in 1-second (FEV1), predicted percentage \|\| id:ukb-b-13405 | MR Egger | 10 | 0.259 | 6.14E-01 | -0.708 | 1.225 | 73.317 | 1.07E-12 | -1.56E-02 | 0.401 |
| Forced expiratory volume in 1-second (FEV1), predicted percentage \|\| id:ukb-b-13405 | Inverse variance weighted | 10 | -0.169 | 1.02E-01 | -0.372 | 0.034 | 80.522 | 1.27E-13 | | |
| Forced vital capacity (FVC) \|\| id:ukb-b-7953 | MR Egger | 10 | 0.072 | 6.92E-01 | -0.271 | 0.414 | 42.461 | 1.11E-06 | -5.92E-03 | 0.370 |
| Forced vital capacity (FVC) \|\| id:ukb-b-7953 | Inverse variance weighted | 10 | -0.090 | 1.42E-02 | -0.163 | -0.018 | 47.260 | 3.51E-07 | | |
| Fractured/broken bones in last 5 years \|\| id:ukb-b-13346 | MR Egger | 10 | -0.010 | 7.25E-01 | -0.066 | 0.045 | 3.554 | 8.95E-01 | -1.96E-04 | 0.851 |
| Fractured/broken bones in last 5 years \|\| id:ukb-b-13346 | Inverse variance weighted | 10 | -0.016 | 9.07E-03 | -0.028 | -0.004 | 3.591 | 9.36E-01 | | |
| Frequency of tiredness / lethargy in last 2 weeks \|\| id:ukb-b-929 | MR Egger | 10 | 0.058 | 6.80E-01 | -0.208 | 0.325 | 22.737 | 3.72E-03 | -2.32E-03 | 0.645 |

| | | | | | | | | | | |
|---|---|---|---|---|---|---|---|---|---|---|
| Frequency of tiredness / lethargy in last 2 weeks \|\| id:ukb-b-929 | Inverse variance weighted | 10 | -0.006 | 8.41E-01 | -0.060 | 0.049 | 23.389 | 5.38E-03 | | |
| Major depressive disorder (ICD-10 coded) \|\| id:ebi-a-GCST005903 | MR Egger | 9 | 0.030 | 3.08E-01 | -0.023 | 0.083 | 2.845 | 8.99E-01 | -1.11E-03 | 0.285 |
| Major depressive disorder (ICD-10 coded) \|\| id:ebi-a-GCST005903 | Inverse variance weighted | 9 | -0.001 | 9.00E-01 | -0.013 | 0.011 | 4.184 | 8.40E-01 | | |
| Number of older siblings \|\| id:ukb-b-1997 | MR Egger | 10 | 0.136 | 3.72E-01 | -0.146 | 0.417 | 4.619 | 7.97E-01 | -3.51E-03 | 0.512 |
| Number of older siblings \|\| id:ukb-b-1997 | Inverse variance weighted | 10 | 0.039 | 1.96E-01 | -0.020 | 0.099 | 5.090 | 8.26E-01 | | |
| Parental longevity (combined parental age at death) \|\| id:ebi-a-GCST006702 | MR Egger | 9 | 0.119 | 8.66E-01 | -1.220 | 1.459 | 141.911 | 2.02E-27 | -5.20E-03 | 0.836 |
| Parental longevity (combined parental age at death) \|\| id:ebi-a-GCST006702 | Inverse variance weighted | 9 | -0.024 | 8.70E-01 | -0.305 | 0.258 | 142.843 | 6.08E-27 | | |
| Peak expiratory flow (PEF) \|\| id:ukb-b-12019 | MR Egger | 10 | 0.048 | 7.56E-01 | -0.244 | 0.339 | 24.543 | 1.86E-03 | -1.90E-03 | 0.729 |
| Peak expiratory flow (PEF) \|\| id:ukb-b-12019 | Inverse variance weighted | 10 | -0.004 | 8.85E-01 | -0.063 | 0.054 | 24.938 | 3.04E-03 | | |
| Platelet crit \|\| id:ukb-d-30090_irnt | MR Egger | 11 | -0.264 | 5.69E-01 | -1.139 | 0.612 | 165.371 | 5.68E-31 | 1.87E-02 | 0.265 |
| Platelet crit \|\| id:ukb-d-30090_irnt | Inverse variance weighted | 11 | 0.256 | 5.96E-03 | 0.074 | 0.439 | 191.360 | 1.02E-35 | | |
| Smoking status: Current \|\| id:ukb-a-225 | MR Egger | 10 | -0.018 | 7.31E-01 | -0.118 | 0.082 | 18.016 | 2.11E-02 | 4.09E-04 | 0.827 |
| Smoking status: Current \|\| id:ukb-a-225 | Inverse variance weighted | 10 | -0.007 | 4.98E-01 | -0.027 | 0.013 | 18.130 | 3.37E-02 | | |
| Sodium in urine \|\| id:ukb-a-335 | MR Egger | 10 | -0.036 | 8.26E-01 | -0.346 | 0.274 | 16.722 | 3.31E-02 | 1.42E-03 | 0.807 |
| Sodium in urine \|\| id:ukb-a-335 | Inverse variance weighted | 10 | 0.003 | 9.25E-01 | -0.059 | 0.065 | 16.855 | 5.10E-02 | | |
| Squamous cell lung cancer \|\| id:ieu-a-967 | MR Egger | 9 | 0.604 | 8.77E-01 | -6.795 | 8.003 | 62.873 | 4.02E-11 | -2.20E-02 | 0.876 |
| Squamous cell lung cancer \|\| id:ieu-a-967 | Inverse variance weighted | 9 | 0.007 | 9.93E-01 | -1.601 | 1.615 | 63.110 | 1.14E-10 | | |
| triglycerides \|\| id:ieu-b-111 | MR Egger | 11 | -0.431 | 4.41E-01 | -1.480 | 0.617 | 310.748 | 1.37E-61 | 1.24E-02 | 0.527 |
| triglycerides \|\| id:ieu-b-111 | Inverse variance weighted | 11 | -0.087 | 4.14E-01 | -0.295 | 0.122 | 325.681 | 5.71E-64 | | |
| Type 2 diabetes \|\| id:ebi-a-GCST006867 | MR Egger | 7 | 0.097 | 9.17E-01 | -1.623 | 1.816 | 27.312 | 4.96E-05 | -5.34E-03 | 0.870 |
| Type 2 diabetes \|\| id:ebi-a-GCST006867 | Inverse variance weighted | 7 | -0.050 | 7.92E-01 | -0.426 | 0.325 | 27.475 | 1.18E-04 | | |
| Usual walking pace \|\| id:ukb-b-4711 | MR Egger | 10 | 0.087 | 4.24E-01 | -0.115 | 0.289 | 24.804 | 1.68E-03 | -4.01E-03 | 0.306 |
| Usual walking pace \|\| id:ukb-b-4711 | Inverse variance weighted | 10 | -0.023 | 2.92E-01 | -0.067 | 0.020 | 28.511 | 7.83E-04 | | |
| Vascular/heart problems diagnosed by doctor: Heart attack \|\| id:ukb-b-11590 | MR Egger | 10 | -0.033 | 3.65E-01 | -0.101 | 0.035 | 46.525 | 1.89E-07 | 9.41E-04 | 0.468 |
| Vascular/heart problems diagnosed by doctor: Heart attack \|\| id:ukb-b-11590 | Inverse variance weighted | 10 | -0.008 | 2.96E-01 | -0.022 | 0.007 | 49.896 | 1.13E-07 | | |
| Vascular/heart problems diagnosed by doctor: High blood pressure \|\| id:ukb-b-14177 | MR Egger | 10 | 0.028 | 6.41E-01 | -0.086 | 0.143 | 15.455 | 5.09E-02 | -1.65E-03 | 0.450 |

| | | | | | | | | | | |
|---|---|---|---|---|---|---|---|---|---|---|
| Vascular/heart problems diagnosed by doctor: High blood pressure \|\| id:ukb-b-14177 | Inverse variance weighted | 10 | -0.017 | 1.63E-01 | -0.041 | 0.007 | 16.671 | 5.41E-02 | | |
| Vascular/heart problems diagnosed by doctor: Stroke \|\| id:ukb-b-8714 | MR Egger | 10 | 0.000 | 9.83E-01 | -0.028 | 0.028 | 11.651 | 1.67E-01 | -7.37E-05 | 0.888 |
| Vascular/heart problems diagnosed by doctor: Stroke \|\| id:ukb-b-8714 | Inverse variance weighted | 10 | -0.002 | 5.49E-01 | -0.007 | 0.004 | 11.682 | 2.32E-01 | | |
| Wheeze or whistling in the chest in last year \|\| id:ukb-b-18335 | MR Egger | 10 | -0.088 | 4.92E-01 | -0.326 | 0.151 | 76.965 | 1.99E-13 | 3.75E-03 | 0.412 |
| Wheeze or whistling in the chest in last year \|\| id:ukb-b-18335 | Inverse variance weighted | 10 | 0.015 | 5.48E-01 | -0.035 | 0.065 | 84.173 | 2.39E-14 | | |
| Whole body fat-free mass \|\| id:ukb-b-13354 | MR Egger | 10 | 0.016 | 9.15E-01 | -0.259 | 0.291 | 48.001 | 9.87E-08 | -2.36E-03 | 0.649 |
| Whole body fat-free mass \|\| id:ukb-b-13354 | Inverse variance weighted | 10 | -0.049 | 8.36E-02 | -0.105 | 0.007 | 49.344 | 1.43E-07 | | |
| Years of schooling \|\| id:ieu-a-1239 | MR Egger | 10 | -0.185 | 8.03E-02 | -0.366 | -0.004 | 11.073 | 1.98E-01 | 6.27E-03 | 0.093 |
| Years of schooling \|\| id:ieu-a-1239 | Inverse variance weighted | 10 | -0.013 | 5.70E-01 | -0.057 | 0.031 | 16.111 | 6.46E-02 | | |

**Table S10. APOE allele determination.**

*APOE* allele determination based on genotype of alleles rs429358 and rs7412.

*Ambiguous alleles were coded as ε2ε4 due to the rarity of ε1 allele.

** Extremely rare, omitted from analyses

| APOE alleles | rs429358 genotype | rs7412 genotype |
|---|---|---|
| ε4ε4 | C/C | C/C |
| ε3ε4 | T/C | C/C |
| ε2ε4* | T/C | T/C |
| ε3ε3 | T/T | C/C |
| ε2ε3 | T/T | T/C |
| ε2ε2 | T/T | T/T |
| ε1ε3* | T/C | T/C |
| ε1ε4** | C/C | T/C |
| ε1ε2** | C/T | T/T |
